# Psychiatric polygenic profiles characterize difficult-to-treat depression versus antidepressant responders in the AGDS:Cell-o cohort

**DOI:** 10.1101/2025.09.16.25335873

**Authors:** BL Mitchell, D Gilroy, LA Wallace, J Kiewa, R Parker, L Nunn, A Walker, T Lin, L Ziser, G English, M O’Neil, RAN Pertile, S McKenna, SJ Hockey, A Hill, A Shim, T To, A Treneman, AM McIntosh, JJ Crouse, M Ziller, CA Olsen, DC Whiteman, EM Byrne, PA Lind, SE Medland, NG Martin, IB Hickie, AK Henders, NR Wray

## Abstract

**Background:** Major depressive disorder is highly heterogeneous, yet clinically useful subtypes remain difficult to define. Multiple studies align to suggest one-third of people respond to first line antidepressants, one-third respond after some trial-and-error, while one-third remain treatment resistant (difficult-to-treat depression, DTD). Early identification of individuals on a trajectory toward DTD is an important research goal.

**Methods:** Using 4.5 years of prescription records together with three self-report measures we defined seven stringently defined treatment-based groups to stratify participants of the Australian Genetics of Depression Study (AGDS). Four groups comprised antidepressant responders (based on different specific antidepressants), two were DTD groups, and one a bipolar disorder group. We contrasted the groups by polygenic scores (PGS) for psychiatric traits and self-report phenotypes. Based on group allocations we recruited the AGDS:Cell-o sub-cohort; participants provided a blood sample processed to allow future characterisation through cell-based assays.

**Results:** Of 9,964 AGDS eligible individuals, 5,381 (54%) could be classified into one of the seven groups. All groups had significantly higher PGS compared to controls across multiple psychiatric traits and the DTD groups had significantly higher PGS across multiple psychiatric traits compared to the treatment responding groups. 721 AGDS participants were recruited into AGDS:Cell-o.

**Conclusions:** PGS differences between treatment responder and DTD groups imply biological differences between the groups. The collection of the AGDS:Cell-o cohort paves the way for generating cell-based (e.g., induced pluripotent stem cell) assays that could stratify patients at an early stage in the treatment course. This could underpin clinical trials and ultimately revised treatment regimes.

## Introduction

Major depressive disorder (MDD) is a heterogeneous condition(1). While subtyping based on clinical features has been an active area of debate(2, 3) and research(4-6) for many decades, translation to clinically useful subtypes has not been achieved. Here, we consider an approach to subtyping based on antidepressant medication use.

Consistent across international treatment guidelines is the recommendation that selective serotonin reuptake inhibitors (SSRIs) or serotonin-norepinephrine reuptake inhibitors (SNRIs) should be prescribed as a first-line medication owing to their favourable tolerability profiles(7-9). Meta-analyses of clinical trials and head-to-head comparisons(10, 11) provide an evidence-base to support clinicians in guiding patients through a sequential trial-and-error approach to find an antidepressant that works for them. While the majority find an antidepressant adopted for sustained-use across multiple episodes of depression(12), one-third of people with MDD are reported to be treatment resistant(13, 14). A major clinical challenge in the care of people presenting with depression symptoms is the early identification of those likely to follow a treatment-resistant course and differentiating them from people presenting in a depressive episode of bipolar disorder (BIP). Early identification would enable research to improve clinical outcomes.

Our study is embedded within the phenotypically rich Australian Genetics of Depression Study (AGDS)(15). The majority of AGDS participants have genome-wide genotype data(16) and have self-reported their use, response and tolerability to antidepressants(17). A subset of participants consented to data linkage with prescription record data. Here, we report analyses on stringently defined treatment response and difficult-to-treat depression (DTD). A targeted set of AGDS participants were invited to participate in the sub-study, AGDS:Cell-o, based on their group allocation. Group assignments remained robust despite 7 years between prescription records used to define the groups and recruitment into the study. AGDS:Cell-o participants provided blood samples which were processed to harvest peripheral blood mononuclear cells (PBMCs) to allow future biomarker research, including for lymphoblastoid cell line (LCL), induced pluripotent stem cell (iPSC)(18), organoid(19) or assembloid (20) research.

## METHODS

A full description of the AGDS cohort was provided by Byrne *et al*(15) and with incorporation of genome-wide genotypes by Mitchell *et al*.(16, 21) Briefly, N = 20,689 participants aged 18 years or older and from across Australia were recruited during 2016–2019. After the initial data freeze(15), recruitment remained open so that the cohort totals N = 22,263. These participants self-reported having had major depression at some point in their lifetime and completed a comprehensive online questionnaire specifically designed to evaluate the genetic and psychosocial factors that contribute to the risk of depression and individual responses to antidepressant medications. The survey included the Composite International Diagnostic Interview Short Form (CIDI–SF) (which can be used to assign a lifetime diagnosis of MDD), and a questionnaire about experiences with regularly prescribed antidepressants. Like other volunteer studies(22, 23), participants in the AGDS study report a significantly higher level of education compared to census data(15). The majority of AGDS participants consented to provide a saliva sample for genetic analysis. Genome-wide genetic data were generated on the Illumina global screening array, and data from N=15,937 passed standard quality control (see Mitchell *et al* (16, 21) for full details).

Approval for consented record linkage to records of prescriptions dispensed under the Pharmaceutical Benefits Scheme (PBS) was obtained from the Australian Government Department Services Australia External Request Evaluation Committee prior to the commencement of the AGDS study. The majority of AGDS participants (N=16,657) consented to linkage to PBS data. Consented record linkage was available for 4.5-year period covering July 1, 2013, to December 31, 2017. Of these N =13,861 participants had genome-wide genetic data.

### Antidepressant usage groups

As expected for an Australian Study, the majority (>75%) of AGDS participants are of European ancestry (inferred from genetic data), with many global ancestries contributing to the remainder.^15^ Given the novel goals of our study, we focussed on European ancestry to reduce heterogeneity associated with genetic differences across ancestral groups. From AGDS, N=9,964 participants with CIDI-SF defined MDD, inferred European ancestry, genome-wide genotypes, and at least one antidepressant prescription record were prospective participants for allocation into one of seven mutually exclusive groups based on antidepressant use derived from prescription data. Four of the seven groups comprised treatment response groups: two for SSRIs (sertraline or escitalopram/citalopram) and for two for Serotonin-Norepinephrine Reuptake Inhibitors (SNRIs) (desvenlafaxine/venlafaxine or duloxetine). Treatment response was defined as sustained-use (> 20 prescriptions of the specific antidepressant in the 4.5 year window) and a response of ‘very well’ or ‘moderately well’ to the self-report efficacy question “How well does/did (name of antidepressant) work for you?’, with response options of ‘very well’, ‘moderately well’, ‘not at all well’ and ‘don’t know’. Two groups were defined to represent DTD, one required self-reported treatment or recommended treatment with electro-convulsive therapy (ECT), no more than 10 scripts of the same antidepressant, and no self-report of any antidepressant working “moderately well” or “very well”. The other DTD group was defined by prescription records showing augmentation of antidepressants with either lithium or antipsychotics for at least 10 scripts. The final group had self-reported diagnosis of BIP and prescription history of lithium (BIP+L group). The inclusion and exclusion criteria are described in **Table 1**. Treatment-resistant depression (TRD) is typically defined as MDD that does not significantly improve after adequate trials of at least two different antidepressants(24). Given that our groups were defined without direct knowledge of improvement, we use DTD over TRD but our expectation is that DTD would be strongly concordant with TRD.

**Table 1.**
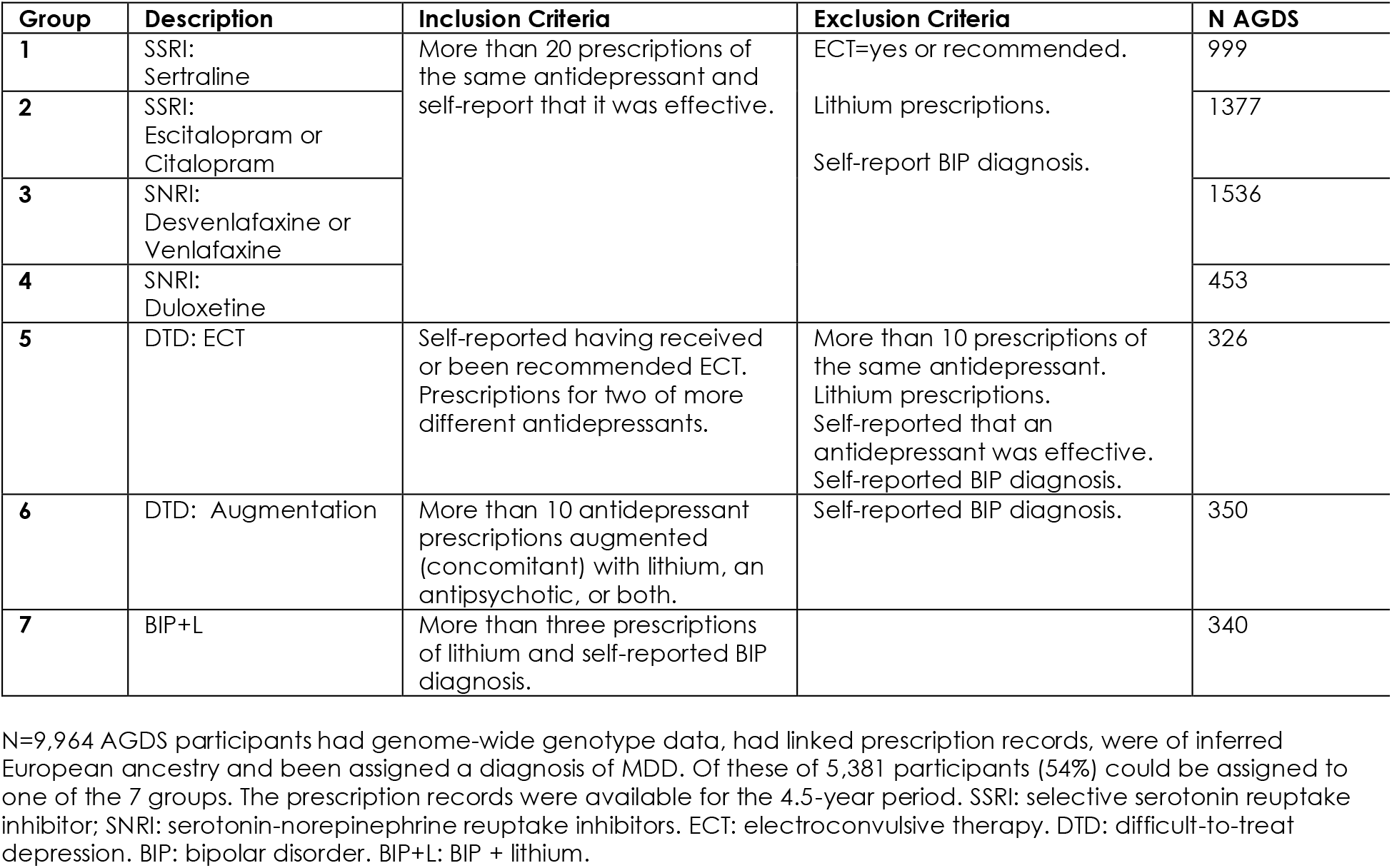
Definition of prescription-based groups in AGDS.

### Analysis

SBayesRC(25) polygenic scores (PGS) for 5 major psychiatric disorders that had well powered genome-wide association study (GWAS) data were calculated for all AGDS participants with genotype data (N=15,937): major depression (MD)(26), BIP(27), schizophrenia(SCZ)(28), ADHD(29) and major anxiety disorders (ANX)(30). The GWAS data used were independent of the AGDS cohort (i.e., Australian cohorts had been excluded from the MD and BIP GWAS). The PGS were each standardised to an Australian control cohort (21) (N = 13,696). Mean differences in PGS between groups were tested with the two sided t-test. We also fitted linear models that included within cohort genetic principal components (PCs; Group ∼ PCs + PGS); any differences in association p-values did not impact conclusions, so we present the more parsimonious t-test with direct visualisation of group means. There was a natural hierarchy built into our study design. We first tested differences in mean PGS between pairs of groups, i.e, between the two SSRI responding groups, between the two SNRI responding groups, and between the two DTD groups. Groups were combined if there was no evidence for differences between them. The primary analysis contrasted the combined SSRI and SNRI groups with the combined DTD groups; differences with the BIP+L group were also tested. Significance was declared after correction for 3 contrasts × 5 = 10 tests p < 0.05/15 = 3.3 ×10^-3^. Secondary analyses of these combined groups considered PGS derived from the genomic structural equation modelling applied to GWAS summary statistics of 14 childhood- and adult-onset disorders (31) (the 5 disorders already considered were counted amongst this 14, although sometimes less powerful GWAS had been included; AGDS participants would have been included in the GWAS of MD and BIP that contributed to the modelling comprising 1.8% and 0.8% of the case sample size, respectively). SBayesRC PGS were calculated for each AGDS individual for the P-factor (factor shared across disorders) and 5 latent factors (F1-F5). We calculate the probability of a participant in the DTD group being ranked higher than a participant in the SSRI/SNRI from the pROC R package (area under the curve).

The baseline questionnaire completed by AGDS participants was extensive (445 questions), analyses of which have been previously reported. Two high level phenotypes that have been derived are atypical depression(32) and circadian depression(33). Atypical depression was assigned if participants reported both weight gain and hypersomnia during their worst depression episode (0.21 of AGDS). Circadian depression was assigned based on endorsements to seasonal changes in sleep, mood, appetite, activity and to sleep and chronotype questions (0.23 of AGDS). Circadian and atypical assignment were correlated (0.20, p < 10^-16^); 0.09 were assigned both atypical and circadian types. We used contingency table tests to test for differences in frequency of atypical depression, circadian depression and self-reported anxiety disorders, contrasting the SSRI/SNRI responding group with the DTD group. Statistical significance after accounting for 3 tests was set at p< 0.05/3 = 1.67 ×10^-2^.

Anyone in AGDS who self-reported BIP had subsequently been invited into the Australian Genetics of BIP Study(34) where online questionnaires probed diagnoses of BIP type 1 (characterised by a severe episode of mania and recurrent episodes of major depression) or type 2 (characterised by having a hypomania episode and recurrent episodes of major depression). We cross-checked BIP diagnoses for those allocated to our BIP+L group. All analyses were conducted in R version 2024.04.24.

### AGDS:Cell-o Recruitment

Having allocated AGDS participants to our prescription-informed groups, we set out to collect blood samples from participants from each group and process the blood to allow harvesting of PBMCs; the AGDS Cell-omics (AGDS:Cell-o) study. Full details of the recruitment, questionnaire, and protocols for collection and processing of biological samples are provided in the **Supplementary Methods**. Briefly, AGDS:Cell-o participants completed a detailed online questionnaire (116 questions) which was designed to focus on experiences with treatments for depression. Participants were blinded to their group allocation. We used questionnaire responses to determine if participants remained adherent to their group allocation (see **Table 2** for criteria).

**Table 2.** AGDS:Cell-o sub-cohort recruitment.

| Group | Description | Flagged as adherent-to-group criterion using responses from AGDS:Cell-o questionnaire | N recruited (N Flagged; %) |
| --- | --- | --- | --- |
| 1 | SSRI: Sertraline | Included the medication matching their group assignment amongst their 'most effective' medications, either as an individual drug or in combination.<br>Excluded if self-reported diagnosis of BIP and were currently using lithium.<br>Excluded if ECT was amongst their 'most effective' physical therapies. | 113 (90; 80%) |
| 2 | SSRI: Escitalopram or Citalopram |  | 147 (97; 66%) |
| 3 | SNRI: Desvenlafaxine or Venlafaxine |  | 192 (103; 54%) |
| 4 | SNRI: Duloxetine |  | 76 (31; 41%) |
| 5 | DTD: ECT | Excluded if self-reported diagnosis of BIP and were currently using lithium as their only psychiatric medication.<br>Excluded if self-reported to have tried fewer than three medications. | 71 (62; 87%) |
| 6 | DTD: Augmentation | Excluded if self-reported diagnosis of BIP and were currently using lithium as their only psychiatric medication<br>Excluded if ECT was amongst their 'most effective' physical therapies.<br>Excluded if self-reported to have tried fewer than three medications. | 64 (48; 75%) |
| 7 | BIP+L | Excluded if did not currently endorse previous diagnosis of bipolar disorder, unless were endorsed in the Australian Genetics of Bipolar Study(34) | 58 (51; 88%) |
|  | Total |  | 721 (482; 67%) |

See **Table 1** for abbreviations.

### Governance

Ethics approval for this study was obtained from the Human Research Ethics Committee (HREC) of QIMR Berghofer (HREC code EC00278, protocol number P2118), and The University of Queensland (HREC code EC00456; protocol number: 2023/HE000050). PBS data access was approved by Services Australia external research ethics committee (MI3967 to QIMR Berghofer).

## RESULTS

### PGS analyses

As an initial benchmark, we tested for differences in PGS between all AGDS participants (N=15,937) and controls (N = 13,696) which updates previous reports with the current PGS. PGS were significantly higher for all 5 psychiatric disorders tested in AGDS participants compared to controls: MD: 0.57; BIP: 0.41 SCZ: 0.18; ADHD: 0.18; ANX: 0.35 (control sample standard deviation units, p < 10^-52^ across all tests) (**Figure 1**). Of the N=9,964 AGDS participants considered for this study 5,381 (54%) could be allocated to one of seven mutually exclusive stringently defined groups. Given the natural hierarchy in our study design, we first contrasted PGS between the two SSRI responding groups, the two SNRI groups and the two DTD groups, finding only small differences in mean PGS which were not significantly different. Consistent with our analyses of sustained-use groups in AGDS(35), we found the mean PGS to be similar across SSRI and SNRI treatment response groups, and formal testing with the sertraline group as the reference showed no significant differences. Similarly, between the two DTD groups the mean PGS were similar and not significantly different. Based on these results, we tested the difference in PGS between the combined SSRI/SNRI group with the combined DTD group. The mean PGS were always higher for the DTD group reaching Bonferroni statistical significance for all tests except ANX: SSRI/SNRI vs DTD MD: 0.54 vs 0.67, difference (Δ) 0.13, p = 3.0 ×10^-3^; BIP 0.38 vs 0.54 Δ = 0.16, p = 7.9×10^-5^; SCZ: 0.14 vs 0.28, Δ = 0.14 p = 1.5×10^-3^; ADHD 0.14 vs 0.26, Δ = 0.12 p = 3.6×10^-3^; ANX 0.34 vs 0.43 Δ = 0.09 p = 2.5×10^-2^ (**Figure 1a; Supplementary Tables 1**).

**Figure 1.**
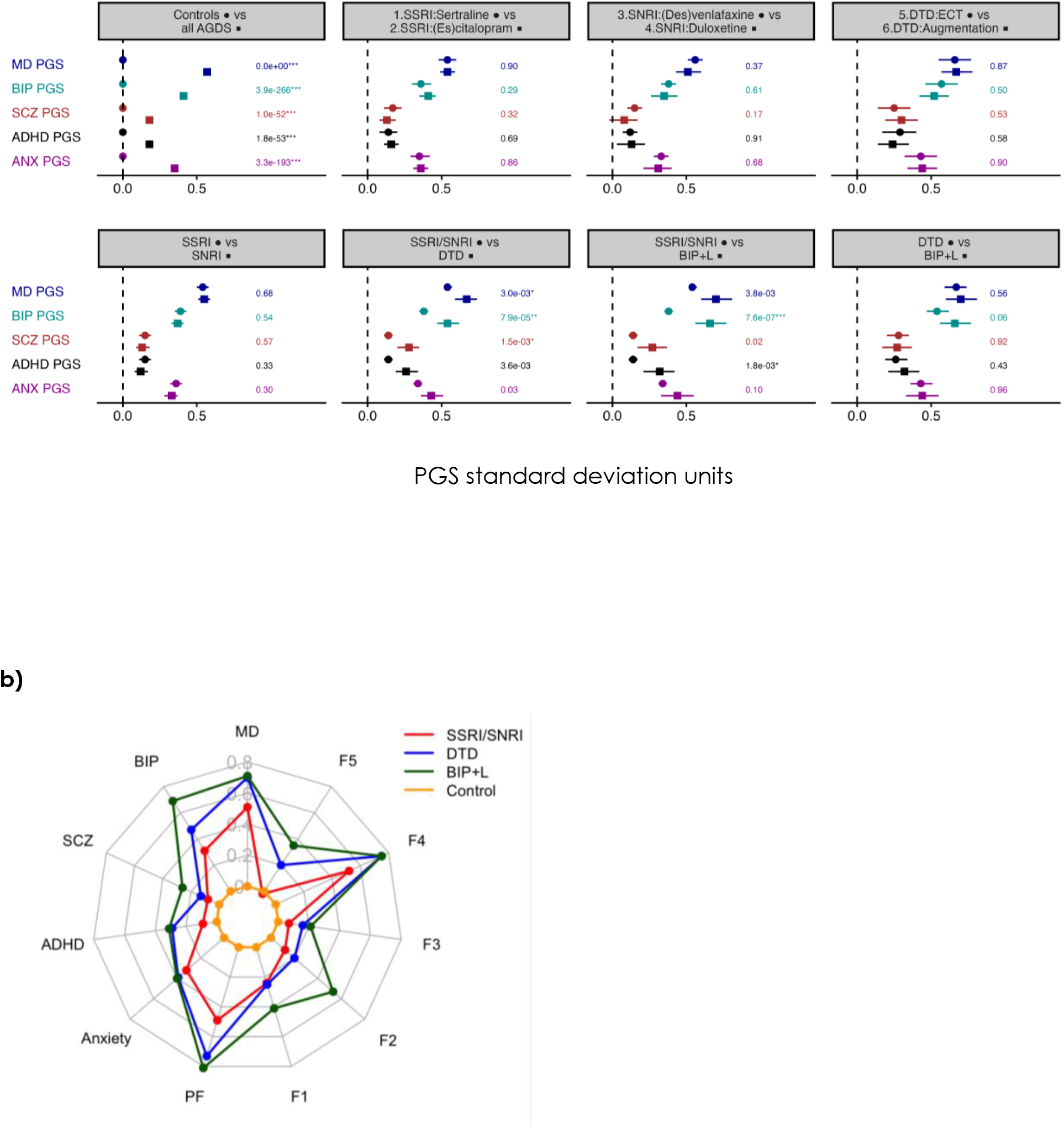
Contrasting groups based on polygenic score analyses. a) PGS for 5 psychiatric disorders scaled to a control sample (N=13,689) so that each PGS in the control sample has mean 0 and standard deviation (SD) of 1. Plotted are mean and 95% confidence interval with numbers being the P-value from a two-sided t-test. b) Radar plot of PGS, contrasting the 4 treatment responding groups combined (SSRI/SNRI red); the two DTD groups combined (blue), the BIP+L group (green), controls (orange). See text for list of significant differences. All results are provided in **Supplementary Tables 1 and 2**. SSRI: selective serotonin reuptake inhibitor; SNRI: serotonin-norepinephrine reuptake inhibitors. ECT: electroconvulsive therapy. DTD: difficult-to-treat depression. BIP: bipolar disorder. BIP+L: BIP + lithium. MD: major depression; SCZ: schizophrenia; ADHD: attention hyperactivity disorder; PF and F1-F5: P-factor and five latent factors from genomic structural equation modelling applied to 14 psychiatric disorders: Labels applied to the 5 factors. F1: Compulsive disorders factor; F2: SCZ-BIP factor; F3: Neurodevelopmental; factor F4: Internalizing disorders factor and F5: Substance-use disorders factor.

We also tested PGS differences using the factor analysis results applied to GWAS of 14 psychiatric disorders(31). The full cohort had significantly higher (p-value < 1 ×10^-13^) mean PGS than controls for all factors tested: PFactor: 0.59; F1 Compulsive disorders factor: 0.26; F2 SCZ-BIP factor: 0.23; F3 Neurodevelopmental factor: 0.13; F4 Internalizing disorders factor: 0.57; F5 Substance-use disorders factor: 0.08 (control standard deviation units). Compared to the SSRI/SNRI group the combined DTD group values were higher for all factors bar F3, and significantly so for some: PFactor: 0.57 vs 0.73, Δ = 0.16, p = 1.3×10^-4^ ; F1: 0.25 vs 0.31, Δ = 0.06, p = 0.16; F2: 0.20 vs 0.32, Δ = 0.12, p = 2.2×10^-3^; F3: 0.15 vs 0.14, Δ = -0.01, p = 0.81;F4: 0.58 vs 0.70, Δ = 0.12, p = 3.8×10^-3^; F5: 0.04 vs 0.12, Δ = 0.07, p = 0.079 (**Figure 1b; Supplementary Table 2**). Despite the significant mean differences between the groups PGS cannot usefully stratify them; for example, the probability that a DTD person is ranked higher than a person in the SSRI/SNRI group using MD PGS is only 0.53 (0.55 using BIP PGS).

The PGS mean values for the BIP+L were not significantly different to those from the DTD group, but point estimates were higher for BIP+L for 9 of the 11 PGS tested, with the biggest differences being for PGS of BIP (0.66 vs 0.54, Δ = 0.12, p = 0.063) and the F2 factor (0.47 vs 0.32, Δ = 0.15, p = 0.039). Taken together these results suggest anti-depressant responders, DTD and BIP+L are more extreme in genetic liability across multiple psychiatric disorders and most importantly that significant genetic differences separate antidepressant responders and DTD (**Figure 1b**).

### Phenotypic differences between groups

Atypical depression was significantly more common in the DTD group compared to the SSRI/SNRI group (0.275 vs 0.205, p = 1.4 ×10^-4)^, but there were no differences in representation of circadian depression between the groups (**Supplementary Table 3**). Given the significantly higher PGS for ANX in the DTD group compared to the SSRI/SNRI group we tested for differences in the rate of self-reported anxiety disorders, recorded in the baseline AGDS questionnaire, endorsed by 0.55 of respondents(15). Although this endorsement represented 0.575 in DTD and 0.536 in SSRI/SNRI groups respectively, this difference was not significant (p = 0.055).

There is an ongoing debate about the validity of a self-report diagnosis of BIP. Hence, in order to make a stringent definition given the data available to us, we had defined our BIP+L group as self-report of BIP diagnosis plus more than 3 records of lithium prescription that included prescriptions not concurrent with antidepressant medication. Of the N = 340 allocated to our BIP+L group N = 199 (0.59) completed the BIP questionnaire in the Australian Genetics of BIP Study(34) with 0.5 and 0.3 endorsing responses which allowed assignment of diagnoses BIP type 1 and type 2, respectively. The remaining 0.2 were considered sub-threshold BIP.

### AGDS:Cell-o Recruitment

A summary of the AGDS:Cell-o study design is presented in **Figure 2**. 3,591 AGDS participants were invited, and 742 (21%) self-recruited into the AGDS:Cell-o portal, fulfilled all protocols and provided a blood sample. Participants landing on AGDS:Cell-o webpage were highly likely to participate in the study. Blood collections were taken across Australia and shipped to our laboratory for processing (**Supplementary Figure 1**). Despite this, of the 742 participants providing blood samples, extracted PBMCs were judged as of acceptable quality for N = 721 (**Supplementary Table 4; Supplementary 2**). These 721 participants comprise the AGDS:Cell-o version 1 resource described here. They did not differ significantly from those invited but not recruited into the resource, across a range of measures (**Supplementary Table 5**). The numbers recruited into each group are presented in **Table 2**.

**Figure 2.**
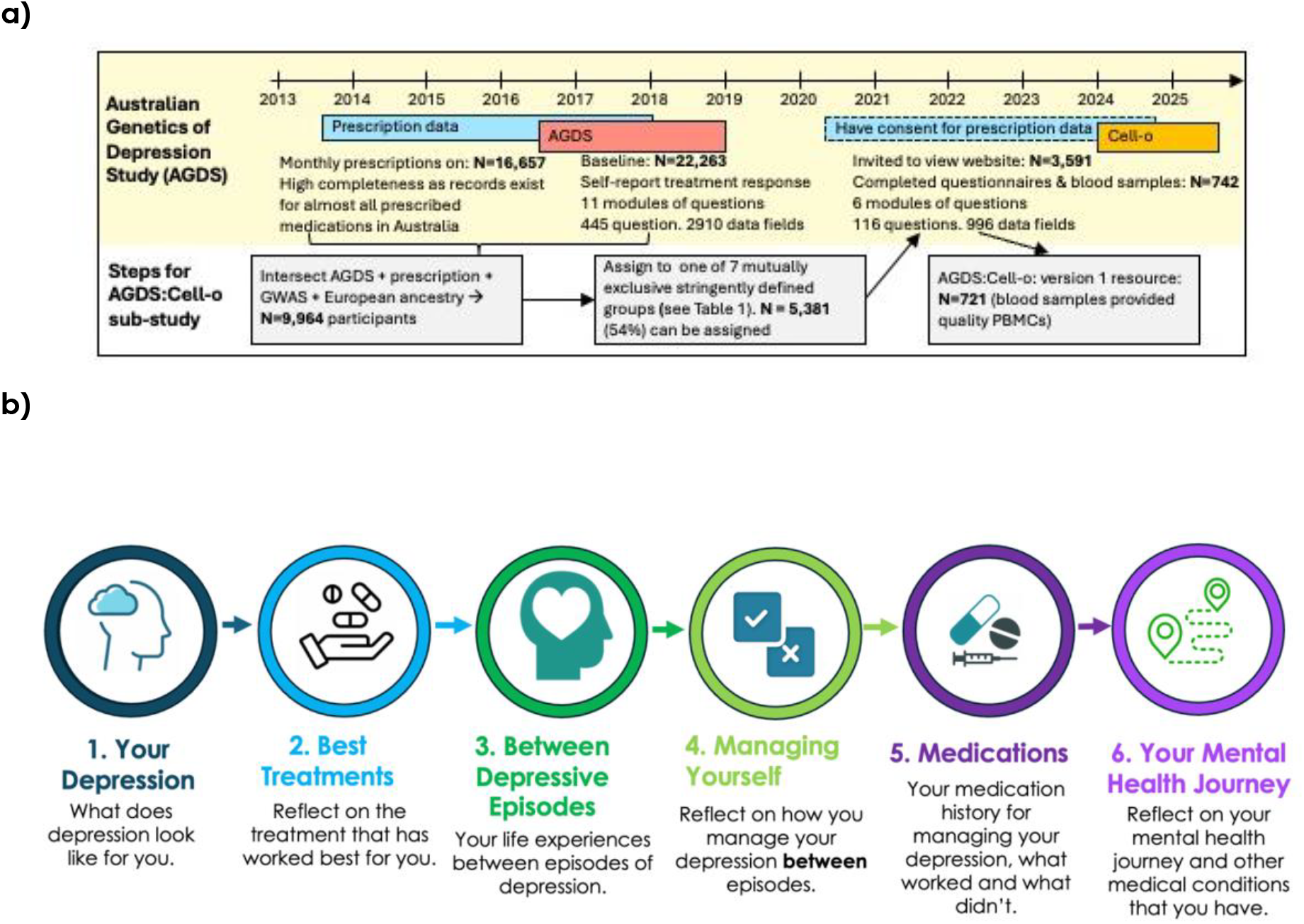
AGDS:Cell-o study design. a) Time and steps for recruitment of the AGDS:Cell-o sub-study b) Modules of the AGDS:Cell-o questionnaire (Full questionnaires Supplementary File 1)

In the AGDS:Cell-o questionnaire, participants were asked to report the single – or combination of – medications that worked best for their depression. 94% of all AGDS:Cell-o participants self-reported currently taking medication associated with psychiatric conditions. Given that our group allocation was based on a 4.5-year snapshot of prescription records, we recognised that some participants may have been allocated to one of the SSRI or SNRI treatment response groups but could later have transitioned to a more DTD classification after sustaining long-term side-effects or intolerability. We flagged participants who indicated that they still tightly adhered to their group allocations, N= 482 (67%) were flagged (**Table 2**). The proportion of AGDS:Cell-o participants self-reporting to be currently taking the medications used to make our groups are provided in **Figure 3** for flagged-as-adherent-to-group participants (and for all recruited participants in **Supplementary Figure 3**; data in **Supplementary Table 6)**. The degree of stringent adherence to group allocation despite the 7-year time period between AGDS baseline recruitment and recruitment into AGDS:Cell-o supports the validity of our approach and allows prioritisation of the flagged adherent-to-group participants for cell-based studies.

**Figure 3.**
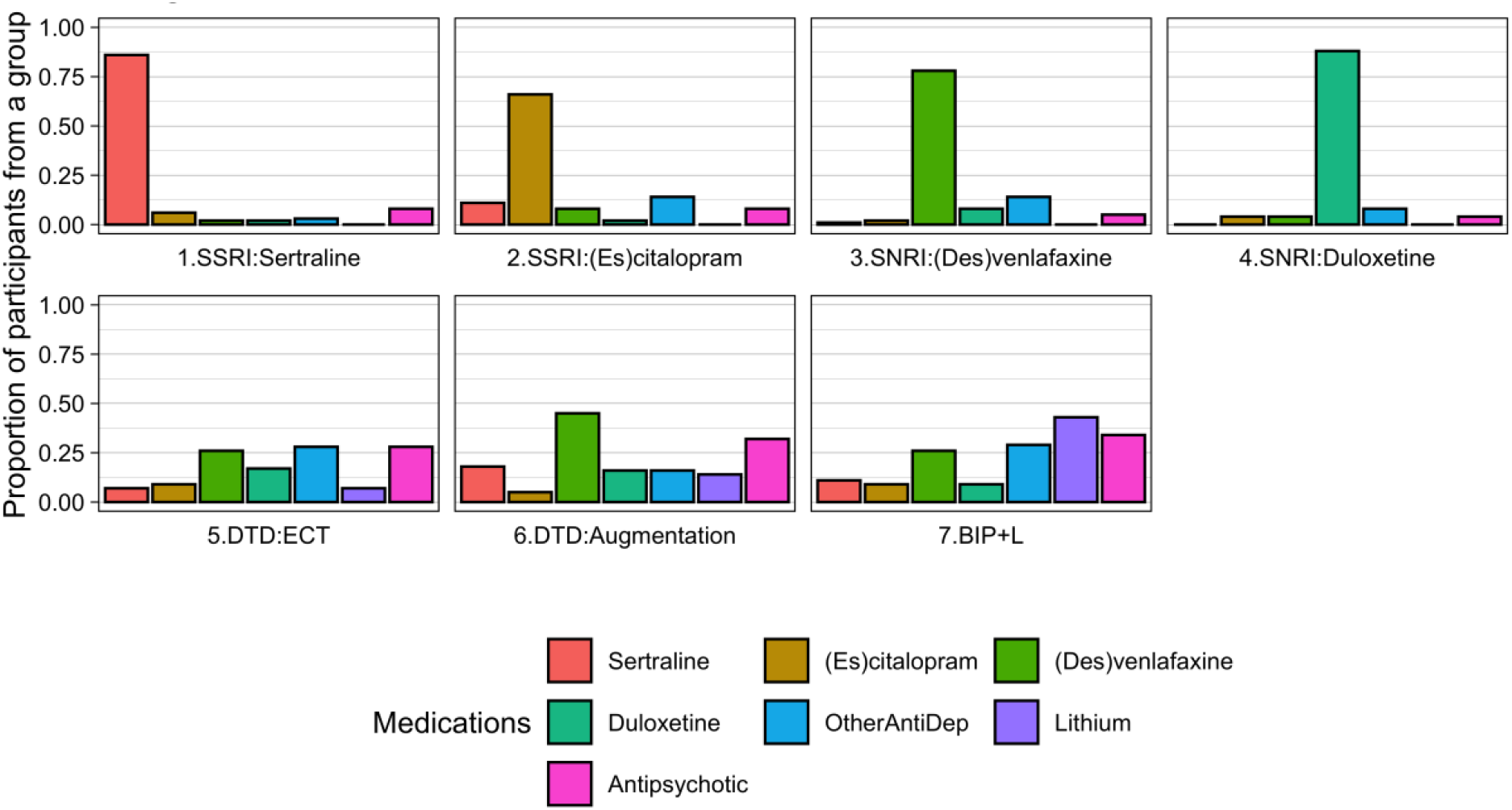
Current self-report medication per group, as recorded in the AGDS-Cello questionnaire for participants flagged as adherent-to-group (N=482) See **Supplementary Table 6** for the values used to generate this graph and **Supplementary Figure 3** for this Figure for all recruited participants.

PGS analyses conducted for the full AGDS cohort were repeated for the 721 participants of the AGDS:Cell-o resource and flagged-as-adherent to group (**Supplementary Table 2**). The point estimates of mean PGS for each group aligned with those from the full AGDS cohort, but given the smaller sample sizes in the AGDS:Cell-o cohort, few differences are recorded as significantly different. The radar plot for the participants flagged into the stringently defined adherent-to-group are supportive of genetic differences between the groups (**Supplementary Figure 4**).

## DISCUSSION

Using only prescription records and 3 self-report items (1. Impression of efficacy of the antidepressant, 2. Recommendation of ECT, 3. BIP diagnosis) we constructed groups of treatment response and DTD amongst AGDS study participants. Genetic (PGS) profiling analyses of psychiatric disorder traits led to three important observations. First, we report no significant differences in PGS profiles between responders to SSRI or SNRI antidepressants. In Australia, prescribing guidelines are similar to those of other countries. While seven antidepressants are currently recommended as first-line treatments (with choices between antidepressants dependant on symptom profiles)(8) the majority of patients in primary care are prescribed sertraline as the first-line treatment, with a transition to other SSRIs and then SNRIs if treatment response is inadequate or if there are undesirable side-effects (36). Our results imply an absence of biological differences in the domains tested between those that were grouped as responders to specific SSRIs or SNRIs. This is consistent with the shared mechanism of SSRIs and SNRIs in their inhibition of the reuptake of monoamine neurotransmitters in the brain, increasing their availability in the synaptic cleft. Relevant results from other AGDS reports are that participants were found to report high co-occurrence in reporting of side-effects for across antidepressants (17) and that sustained-users of SNRIs were found to have significantly higher body mass index (BMI) than sustained-users of SSRIs which was partly explained by BMI PGS (35). The AGDS data did not allow us to distinguish whether the observed difference in BMI association between SSRIs and SNRIs reflects a true causal difference or ascertainment biases, such as clinician/patient preference and switching due to weight gain from first-line SSRIs to SNRIs (the latter consistent with the PGS differences). Including patient preference in first line antidepressant choice has been shown to be significantly associated with adherence to antidepressants over an 8-week trial period (37), which will likely contribute to ultimate sustained use of a specific antidepressant.

Our second observation was that the DTD group had significantly higher PGS profiles compared to SSRI/SNRI responders across the spectrum of psychiatric disorders tested. These results are consistent with previous reports from AGDS (e.g., higher PGS for psychiatric disorders were associated with markers of severity such as age of onset, and number of episodes (16, 21), and with treatment complexity compared to sustained use(35)) as well as from published studies contrasting major depression from clinic recruited cohorts with community recruited cohorts (26) (where the former is expected to be enriched with DTD). However, the simple but stringent group definitions make the differences reported more profound (**Figure 1**). These results imply that biological differences exist between those that respond to established SSRI/SNRI antidepressants and those that do not. However, despite the significant differences at the mean level between groups, PGS are not sufficiently powered for patient stratification at the individual level. Evidence of biological difference motivates the search for biomarkers to stratify groups. Attempts to identify blood biomarkers associated with treatment response is hampered by the lack of prospective studies (38), and studies that take biological samples after treatment response status has been identified are likely to suffer confounding by disease state-associated factors. Recognising these limitations, the AGDS sub-study, AGDS-Cello was established specifically to collect PBMCs for future cell-based studies. Specifically, iPSCs allow study of disease-relevant cell types, and importantly, are largely insulated from confounding factors such as current medication, stress, or disease state, and mostly reflect the genome blueprint of the individual. iPSC perturbation studies (e.g., perturbation with serotonin or lithium) will allow comparison of groups based on treatment-specific responses as well as static differences in unperturbed cells. Pilot studies in treatment resistant depression iPSCs have been positive(39-41), but larger samples sizes and replication studies are needed to validate these encouraging results. Larger scale studies have been pioneered for schizophrenia (42-44) and iPSC studies need to be fast-tracked to address the important clinical need of identifying those who will not be respondent to antidepressants at the time of first prescription. If those identified as treatment resistant could be identified early, much needed research could be advanced to determine effective treatments for these individuals.

Our third observation was that the point estimates were mostly higher for the BIP+L group compared to the DTD group. However, the differences were not significant, which likely reflects the smaller group size for the DTD and BIP+L groups compared to the SSRI/SNRI group. The point-estimate observation implies that DTD is on a cross-disorder genetic liability continuum between those with SSRI/SNRI responding MDD and BIP. We cannot exclude that the DTD group includes people with undiagnosed BIP, but to minimise this possibility, we used self-report of BIP as an exclusion criterion for initial allocation to the DTD groups. Subsequently, in characterisation of AGDS:Cell-o groups, a self-reported BIP diagnosis was an exclusion criterion for our flagged-as-adherent-to-group status. Our BIP+L group was allocated based on self-report diagnosis of BIP within the AGDS cohort. Since depression is a debilitating characteristic of BIP, it is not surprising that those with BIP self-recruited into a study focussed on depression. The Psychiatric Genomics Consortium Working Group for BIP showed a low genetic correlation between cohorts that were clinically recruited compared to self-report diagnosis cohorts (∼0.2) (27). To enrich our BIP group for clinical diagnoses we required evidence of lithium prescriptions and the absence of concurrent antidepressant prescription records. Furthermore, a second self-report of BIP diagnosis was required in order to be flagged-as-adherent-to-group.

Our study provides evidence for biological differences between those that respond to first-line SSRI/SNRI antidepressants and those that do not. A key strength of our study is the large sample size for the baseline cohorts and the stringent assignment of groups. Our group design (two SSRI and two SNRI groups and two DTD groups) enables multiple comparison combinations for exploratory analyses and allows within-cohort replication across groups that *a priori* were expected to be similar to each other. However, our study has some limitations. First, in Australia prescription records available for research comprise only a 4.5-year window prior to the consent date provided by research participants (**Figure 2a**). Hence, the 4.5-year window represents a different snapshot of the treatment trajectory for each individual relative to their date of first diagnosis. A key concern for our recruitment into AGDS:Cell-o was whether the group classifications would be stable over the 7-year period. Participants were blinded to their group allocation, but importantly we found good stability of groups (**Supplementary Figure 3**). We further identified a flagged-as-adherent-to-group (minimum of 48 participants per group **Figure 3**) to be prioritised for future cell-based studies.

A second limitation of our study is that our analyses and AGDS:Cell-o recruitment were restricted to those of inferred European ancestry, a choice that reflects ancestry demographics of Australia and desire to make homogeneous groups given recruitment costs. Another limitation of the study is that our design is case-only. This was a deliberate choice given the interest in comparison between the seven groups and since control cell lines can be sourced from other studies. However, we now have ethics approval to collect blood samples from age- and sex-matched European ancestry Australians who report no lifetime diagnosis of psychiatric conditions. In this way we can include control cell lines in our downstream analyses that have been generated under the same laboratory protocols as our MDD cell lines.

Participants have consented to allow broad sharing of biological samples for research groups operating under approved ethical protocols. We plan a centralised database to enable collaborating researchers to integrate and analyse the multi-layered data, excluding the extensive questionnaire response. An online visualisation tool will further support researchers in designing experiments based on their own inclusion/exclusion criteria. We invite collaborators to access data and cell-lines.

## Data Availability

We invite collaborators to access data and cell-lines. We are open to a range of collaboration models. The participants have consented to sharing of data and biological material international and with both academic and commercial partners. Collaboration agreements require sharing of any data they generate into a central resource.

## ACKNOWLEDGEMENTS

We are indebted to the AGDS participants for giving their time to contribute to this study. We would like to the University of Sydney’s Brain and Mind Centre LEWG for the time taken in constructively evaluating the AGDS:Cell-o questionnaire. We thank Chloe Yap for making Figure 2a, and Preethi Lakshmi for protocol development in building the online questionnaire.

## AUTHOR CONTRIBUTIONS

Concept: NRW; AGDS cohort: NGM, IBH, SEM, EMB, NRW, RP, JJC, PAL; Control cohort: CAO, DCM; Analysis: BLM,JK, NRW,AW, TL, PAL; AGDS:Cell-o Project team: DG, AKH, NRW, IHB, LN, LZ, MO’N, AMMcI; AGDS:Cell-lab team: LAW, AKH, GE, RANP, MZ; University of Sydney Brain and Mind Centre Lived Experience Working Group: Facilitators: SJH, SMcK, IHB, JJC, Members: AH, AS, TT, AT. Manuscript first draft: NRW, JK, BLM. Manuscript review: All authors.

