## SupplementMethodsFigures for "Psychiatric polygenic profiles characterize difficult-to-treat depression versus antidepressant responders in the AGDS:Cell-o cohort"

### In this document

Supplementary Methods
Supplementary Figure 1. Map of location of AGDS:Cell-o participants (N=721)
Supplementary Figure 2. PBMCs sample processing and quality.
Supplementary Figure 3. Current self-report medication per group, as recorded in the AGDS-Cello questionnaire for all recruited participants (N=721)
Supplementary Figure 4. PGS Radar plots

### Excel File

Supplementary Table 1. Polygenic score analysis for the 5 psychiatric disorders
Supplementary Table 2. Polygenic score analysis for full AGDS, AGDS:Cell-o Recruited Participants and AGDS:Cell-o Flagged as adherent-to-group Participants
Supplementary Table 3. Phenotypic subtype analysis
Supplementary Table 4. Numbers at each stage of AGDS:Cell-o cohort collection
Supplementary Table 5. A comparison of invited (but not in resource) and recruited (in resource) participants to AGDS:Cell-o
Supplementary Table 6. Proportion of participants in each group self-reporting to currently taking medications related to the group definitions

### Supplementary File:

AGDS:Cell-o questionnaire

**Supplementary Methods**

### AGDS:Cell-o Recruitment

Selected AGDS participants who had consented to recontact were sent personalised initial invitations from the Principal Investigator of the AGDS study at QIMR Berghofer Medical Research Institute (QIMR Berghofer). A link to the AGDS:Cell-o study website was included (https://auscello.org/). The study website provided information about the study. Each invited participant had a unique code to access the consent page, hosted by the University of Queensland's Human Studies Unit research portal. The QIMR Berghofer research team followed up potential participants who had not consented or responded to the initial email invitation. Follow-up efforts included email and text message reminders, as well as telephone calls, to gauge continued interest in participation. Invitations were issued in small batches between October 2023 and August 2025 to ensure the receiving laboratory could process blood samples within the required timeframe and to monitor for approximately equal recruitment within each of the seven groups. We aimed to recruit ~100 participants from each group. Among prospective participants allocated to each antidepressant group, those aged under 45 years at baseline were invited first; followed by those aged under 65 years. Our online study page included a video(1) 'What to expect if you participate in iPSC research'. Participants who then self-recruited into the study provided informed e-consent. They were offered the opportunity to contact the research team to ask questions and seek clarification. The online consenting process enabled participants to review study information at their own pace; this included interactive options for accessing study information, and a list of frequently asked questions. Participants were invited to consent to data linkage with the original AGDS study and to allow the sharing of data across national and international sites, including collaboration with for-profit organisations. Participants were given the option to consent to a new linkage with Pharmaceutical Benefits Scheme data, which would provide a more recent 4.5 years of prescription dispensing data. A summary of the study design is presented in **Figure 2a**.

### *Governance*

Ethics approval was obtained from the Human Research Ethics Committee (HREC) of QIMR Berghofer (HREC code EC00278, protocol number P2118), and The University of Queensland (HREC code EC00456; protocol number: 2023/HE000050). PBS data access was approved by Services Australia external research ethics committee (MI3967 to QIMR Berghofer). Participants may withdraw from the study at any time. Our report excludes participant withdrawals reported before May 5 2026.

### AGDS:Cell-o Questionnaire

Participants entered the AGDS:Cell-o questionnaire (coded in the LimeSurvey interface) using a unique research participant code. The questionnaire was developed by the research team in consultation with mental health clinicians. Given the extensive questionnaire data collected in the baseline AGDS protocol(2) the AGDS:Cell-o questionnaire was designed to focus on detailed experiences with treatments for depression. A pilot phase recruited 50 participants. At this stage participants' responses were reviewed, and feedback was received from the Lived Experience Working Group (LEWG) at the University of Sydney's Brain and Mind Centre(3). The questionnaire was revised before the recruitment of the remaining of participants. Feedback from LEWG was particularly helpful in improving the user experience of undertaking a long questionnaire. For example, improvements to the functionality of the pause/resume button and allowing participants to elect not to answer questions found distressing.

The questionnaire comprised six sections focussing on symptom profiles, medications, therapies, and other treatments that were self-reported to be used during and between depressive episodes to manage symptoms. In addition, participants reported on other self-reported mental health diagnoses, their general health, and their mental health journey. One section included questions relating to menstruation, pregnancy and menopause history. In total, there were 116 questions and 996 data fields, with provisions for free text boxes at the end of each section (**Figure 2b; Supplementary File 1**).

### Biological samples

All AGDS:Cell-o participants had provided a saliva sample during the AGDS baseline study for the purpose of DNA extraction and genome-wide genotype data had been generated. Each AGDS:Cell-o participant was sent a barcoded biological sample collection kit to take to a local accredited pathology service for blood collection. The kit included instructions for the pathologist, one 9 mL EDTA Vacuette® tube, two 9 mL ACD-A Vacuette® tubes, and a biobottle. All items were enclosed in a laboratory mailer for return via the regular postal system (Australia Post) to the Human Studies Unit laboratory at the University of Queensland for processing. Participants are from across Australia (**Supplementary Figure 1**). To ensure samples were received in a timely manner, participants were instructed to attend pathology clinics for phlebotomy on Monday, Tuesday, or Wednesday so that samples would arrive at the laboratory within the same working week. Owing to the nature of the proposed future research with LCLs and iPSCs (*e.g.*, the possibly endless longevity of derived biological samples and the variety of uses for research), participants were given the option to choose their level of consent i.e., specific: for this project only, extended:  for  use in mental health research,  and unspecified: ongoing future use of their samples.

Whole blood collected in the 9 mL EDTA tube was fractionated to isolate plasma, buffy coat (for DNA), and stored at –80$^{O}$C. Whole blood for PBMC harvesting was collected in ACD-A tubes to ensure high viability of cells for up to 72 hours from time of collection. PBMCs were isolated using the EasySep™ Direct Human PBMC Isolation Kit (StemCell Technologies) and cryopreserved in CryoStor® CS10 for long-term storage in liquid nitrogen, following the manufacturer's instructions. Despite blood samples being collected across Australia, 91% of PBMC isolations were performed within two days of phlebotomy. We isolated good quality PBMCs (average number of isolated cells per person is 22 $x10^{6}$) from 721 participants. An overview of sample processing and quality is provided in **Supplementary Figure 2**.

We also checked the viability of the frozen PBMCs using a subset of samples (N=18). To assess viability, cells were rapidly thawed in a 37 °C water bath and centrifuged at 500 ×g for 8 minutes in RPMI 1640 medium supplemented with 10 % Fetal Bovine Serum (FBS, Bovogen) and 1 % GlutaMAX™. After centrifugation, cells were resuspended in fresh complete medium and counted using a haemocytometer before being plated in 96-well plates. The results showed that approximately 91% of PBMCs remain viable immediately after thawing, and 81% after 24 h of cell culture.

### BIP+L group

Of N = 58 participants recruited into the BIP+L group, N= 41(71%) had completed the Australian Genetics of BIP Study questionnaire. Of these N = 21 (51%) were recorded as BIP type1, N = 11 (27%) as BIP type 2 and N = 9 (22%) as sub-threshold BIP. The flagged-as-adherent group retained N = 51, N=37 (73%) had completed the BIP questionnaire; of these N = 21 (57%) were recorded as BIP type 1, N = 11 (30%) as BIP type 2 and N = 5 (14%) as sub-threshold BIP. The latter were retained because the participants again self-reported a BIP diagnosis in the AGDS:Cell-o, whereas those dropped from the flagged group no longer self-reported a BIP diagnosis (even though it was self-reported in the baseline questionnaire).

**URL**

What to expect if you participate in iPSC research

**Video: https://www.youtube.com/watch?v=NecrZNf4Gbc**

**Supplementary Figure 1. Map of location of AGDS:Cell-o participants (N=721)**

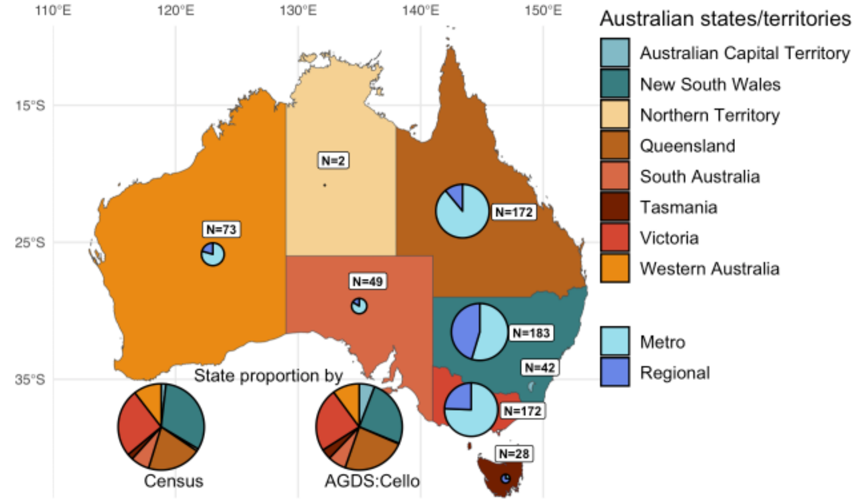

**Supplementary Figure 2. PBMCs sample processing and quality.** a) Schematic of PBMC sample processing (Biorender.com)( b) Cell counts per person and number of days between collection and processing. Data not available for 40 participants. Data from 8 individuals were winsorized to 90. c) Quality of PBMC compared to days to process. Data not available for 40 participants.

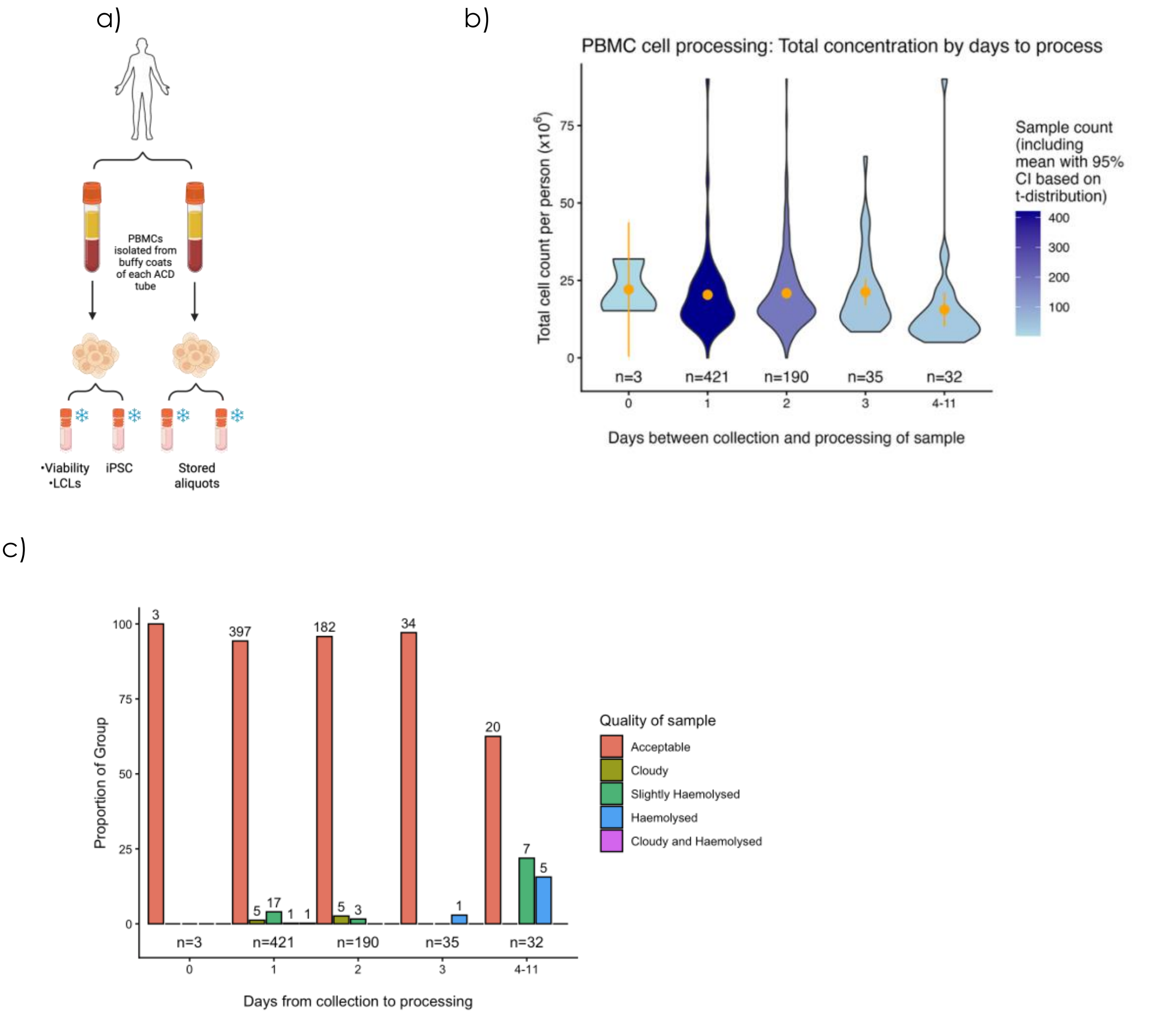

**Supplementary Figure 3. Current self-report medication per group, as recorded in the AGDS-Cello questionnaire for all recruited participants (N=721)**

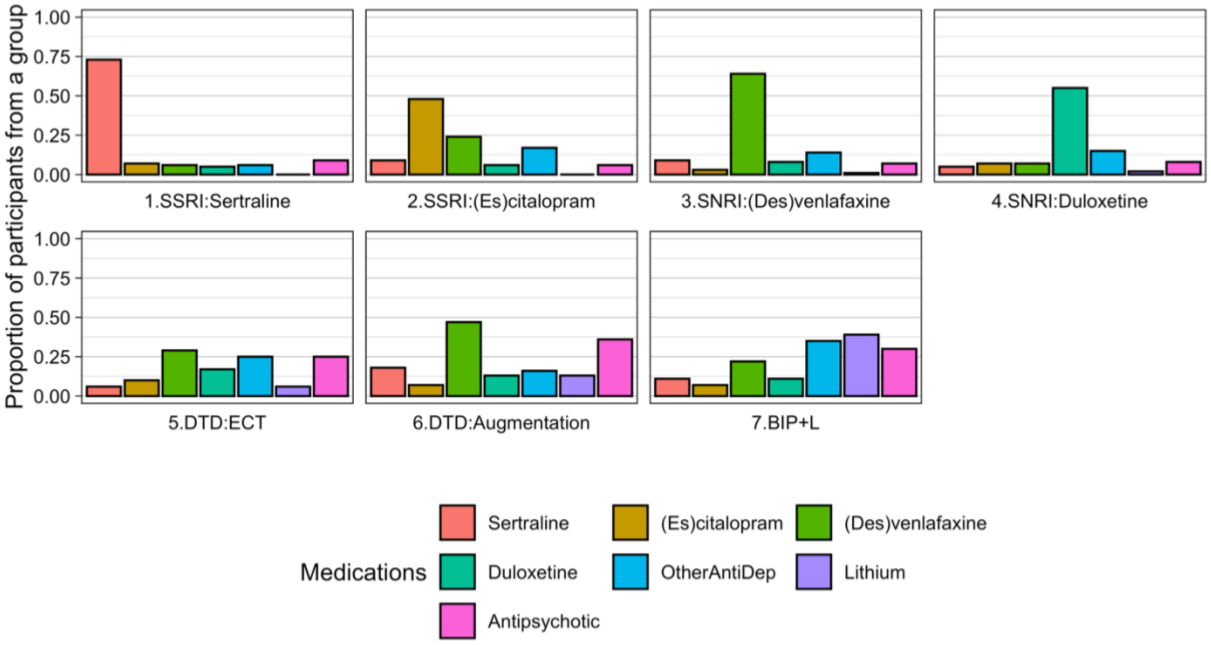


See **Supplementary Table 6** for the values used to generate this graph.

**Supplementary Figure 4. PGS Radar plots**

a) Flagged-as-adherent to group (N=482) b) All recruited participants (N=721)

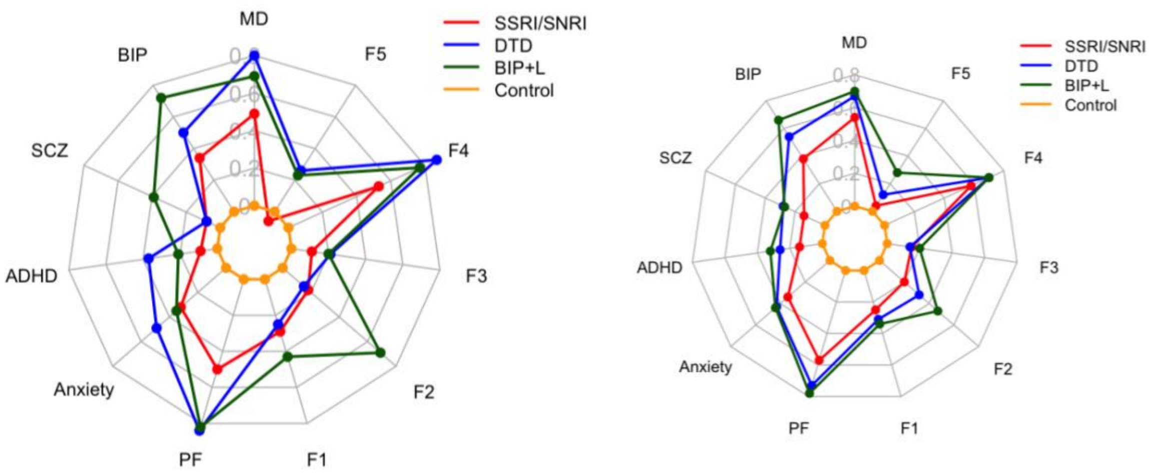
