## SupplementaryFileQuestionnaire for "Psychiatric polygenic profiles characterize difficult-to-treat depression versus antidepressant responders in the AGDS:Cell-o cohort"

### Australian Genetics of Depression Study: Anti-depressant Cell-omics

**People differ in the symptoms they experience as part of their depression and the way in which they respond to treatments. Our long-term goal is that treatments should be tailored to individuals rather than a "one size fits all" approach. This concept of personalised medicine is already applied in other health settings, for example cancer care. Advancing this approach in depression requires more research to understand differences between people.**

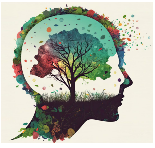

There are 116 questions in this survey.

#### Introduction

We know that depression is different for different people – understanding the reasons for these differences and how and why people respond differently to treatments will help us understand this complex disorder. The following questions relate to your lifetime story/journey and the medications, therapies and other treatments that you have tried to manage your depression as well as your lifetime story/journey with depression. Some people are prescribed medication that works first time around, while others have tried several medications or other treatments before a suitable intervention is found. We are interested in your experiences.

The questionnaire should take approximately 20 - 30 minutes to complete. Please endeavour to answer all questions because this will make our research more robust, but you don't have to answer every question if you don't want to. Before clicking on the "Next" or "Submit" button at the bottom of each page, carefully check your responses as you will not be able to return to prior pages.

Please note that the questionnaire is not mobile-friendly due to individualized logics. For a smoother experience, **we recommend using a desktop, laptop, or tablet**. If you encounter any issues with question progression, please attempt another browser. Should you still experience difficulties, feel free to contact us at.

**Below is a summary of each section of the questionnaire.**

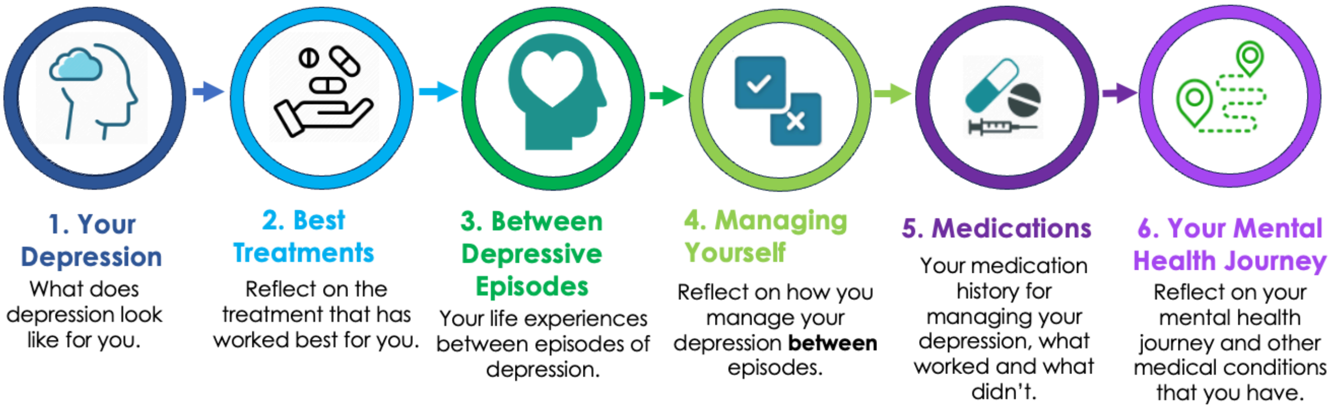

**At the beginning of each section, the above image will be displayed to show your progress through the questionnaire. Completed sections and the section that you are up to will be highlighted in yellow.**

### Informed Consent

**I agree to consent to complete the Personal Journey questionnaire *AGDS-CellO*.**

Please choose **only one** of the following:

- ◯ I agree to participate in this research questionnaire
- ◯ I disagree and do not wish to participate in this research questionnaire

**Thank you for agreeing to complete the questionnaire for our research.**

**If you are unable to complete the questionnaire by yourself you can ask a carer or family member to assist you, or one of our research personnel can help you.**

**Please let us know who will be filling in this questionnaire.**

Only answer this question if the following conditions are met:
((consentInfo.NAOK == "A1"))

Please choose **only one** of the following:

- ◯ I am filling this in myself
- ◯ I will have help from a carer or family member
- ◯ Research personnel are assisting me

**If you wish to assist us in future research, please provide your reason on declination.**

Only answer this question if the following conditions are met:
((consentInfo.NAOK == "A2"))

Please choose **all** that apply:

- ☐ I have changed my mind
- ☐ I do not have time
- ☐ Other: ____________

### Personal Information

#### What is your name?

Please write your answer(s) here:

- First Name
- Last Name

#### What is your Date of Birth?

❗ Please complete all parts of the date.
❗ Answer must be greater or equal to 01/01/1930
Please enter a date:

#### What is your biological sex?

Please choose **only one** of the following:

- ◯ Female
- ◯ Male

#### What is your gender identity?

❗ Choose one of the following answers
Please choose **only one** of the following:

- ◯ Male
- ◯ Female
- ◯ Indeterminate
- ◯ Prefer not to say

#### What is the Australian postcode of your current address?

Please write your answer here:

- 

#### Please provide your contact information

Please write your answer(s) here:

- Email
- Mobile

**What is the best contact should we need to speak with you?**

Please choose **only one** of the following:

- ◯ Email
- ◯ Mobile

### SECTION 1: Your Depression and How You Feel

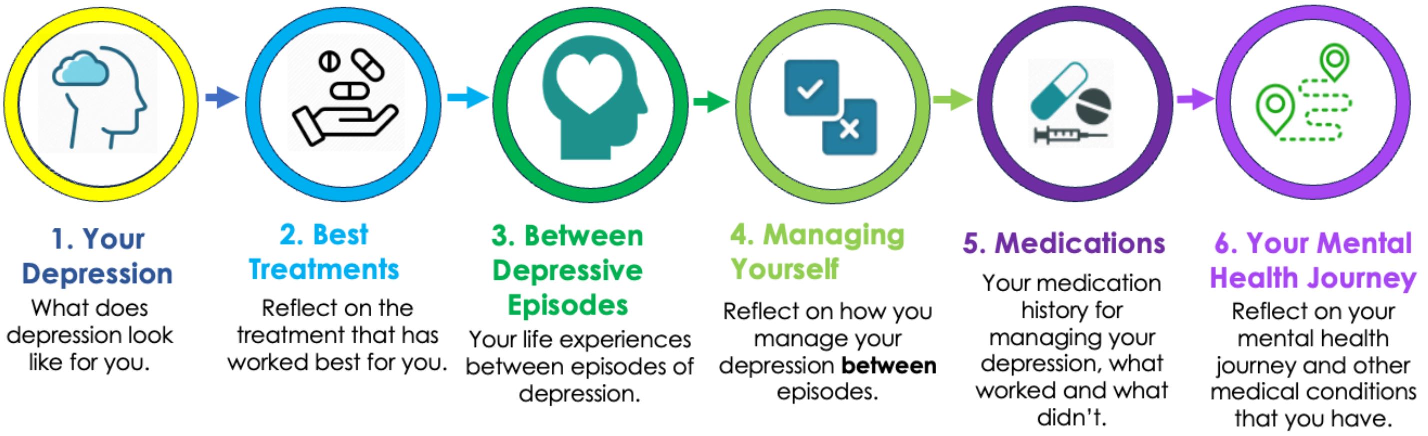

In this section, we want to try and understand how you feel **when you are depressed and struggling with your depression (an episode of depression)**.

Each person has a unique experience with depression and how it makes them feel. Symptoms associated with depression can be grouped into 6 categories; mood, anxiety, cognitive functioning, body clock and sleep, behavioural, and physical symptoms.

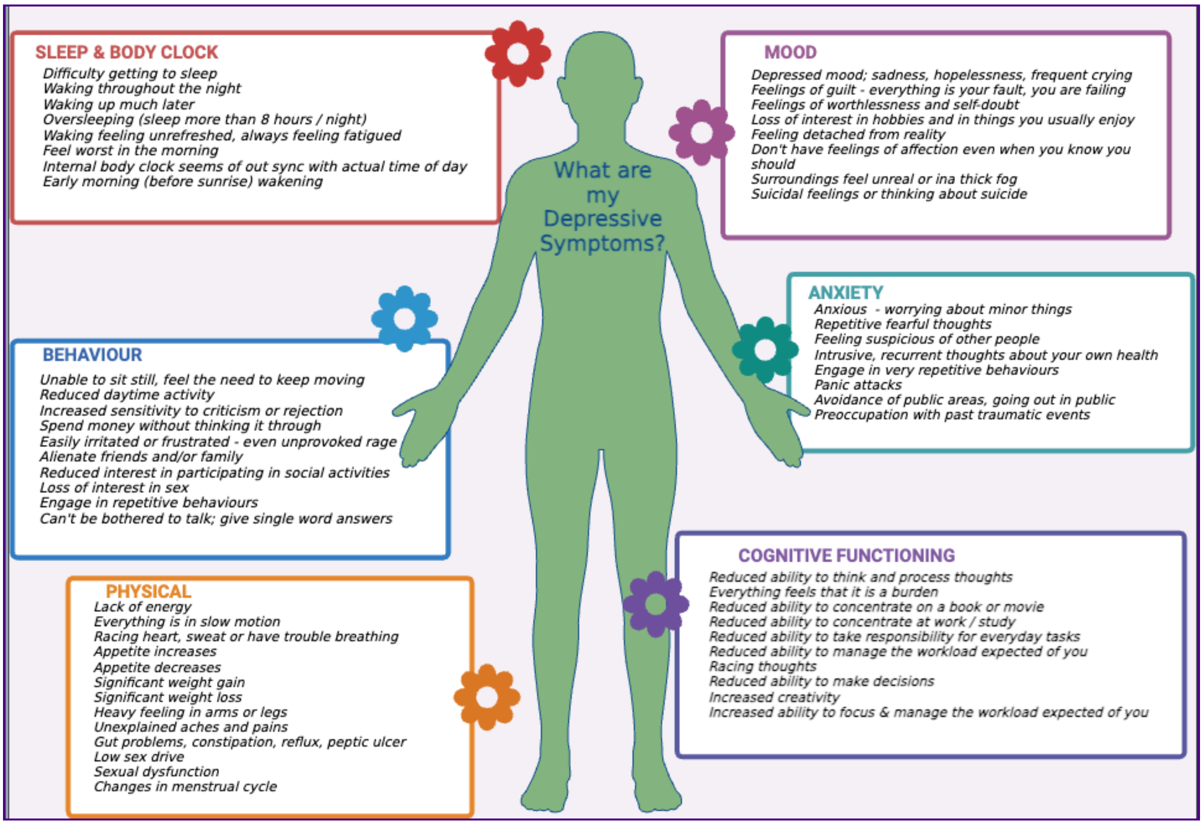

*In this section, there are 10 questions, and depending on your responses, this can increase. We anticipate that this section should take no longer than 10 minutes to complete.*

#### How well do you think you are able to answer questions about symptoms you experience when you are in a depressive episode?

❗ Choose one of the following answers
Please choose **only one** of the following:

- ◯ Not good
- ◯ I am OK at this
- ◯ I know myself well

#### When you are feeling depressed, do you experience any of the following symptoms related to your mood? Please answer YES or NO to each of the symptoms listed.

Please choose the appropriate response for each item:

| | Yes | No |
|---|---|---|
| **Depressed Mood: sadness, hopelessness, frequent crying** | ◯ | ◯ |
| **Feelings of Guilt: that everything is your fault and you are failing** | ◯ | ◯ |
| **Feelings of worthlessness and self-doubt** | ◯ | ◯ |
| **Loss of interest in hobbies and in things you usually enjoyed** | ◯ | ◯ |
| **Feeling detached from reality** | ◯ | ◯ |
| **Don’t have feelings of affection even when you know you should** | ◯ | ◯ |
| **Surroundings feel unreal or in a thick fog** | ◯ | ◯ |
| **Suicidal feelings or thinking about suicide** | ◯ | ◯ |

#### Please rate the impact of these symptoms on your day-to-day life when you are in a depressive episode

Please choose the appropriate response for each item:

| | 1 No Impact | 2 Some Impact | 3 Moderate Impact | 4 Major Impact | 5 Severe Impact |
|---|---|---|---|---|---|
| **Depressed Mood: sadness, hopelessness, frequent crying** | ○ | ○ | ○ | ○ | ○ |
| **Feelings of Guilt: that everything is your fault and you are failing** | ○ | ○ | ○ | ○ | ○ |
| **Feelings of worthlessness and self-doubt** | ○ | ○ | ○ | ○ | ○ |
| **Loss of interest in hobbies and in things you usually enjoyed** | ○ | ○ | ○ | ○ | ○ |
| **Feeling detached from reality** | ○ | ○ | ○ | ○ | ○ |
| **Don’t have feelings of affection even when you know you should** | ○ | ○ | ○ | ○ | ○ |
| **Surroundings feel unreal or in a thick fog** | ○ | ○ | ○ | ○ | ○ |
| **Suicidal feelings or thinking about suicide** | ○ | ○ | ○ | ○ | ○ |

**When you are feeling depressed, do you experience any of the following symptoms related to anxiety? Please answer YES or NO to each of the symptoms listed.**

Please choose the appropriate response for each item:

| | Yes | No |
|---|---|---|
| **Anxious - worrying about minor things** | ◯ | ◯ |
| **Repetitive fearful thoughts - like being afraid of catching an infection, or that a disaster might happen** | ◯ | ◯ |
| **Feeling suspicious of other people** | ◯ | ◯ |
| **Intrusive, recurrent thoughts about your own health** | ◯ | ◯ |
| **Engagement in very repetitive behaviours – like unnecessary washing, cleaning, or checking things over** | ◯ | ◯ |
| **Panic attacks** | ◯ | ◯ |
| **Avoidance of public areas going out in public or travelling outside your home** | ◯ | ◯ |
| **Preoccupation with past traumatic events** | ◯ | ◯ |

#### Please rate the impact of these symptoms on your day-to-day life when you are in a depressive episode

Please choose the appropriate response for each item:

| | 1 No Impact | 2 Some Impact | 3 Moderate Impact | 4 Major Impact | 5 Severe Impact |
|---|---|---|---|---|---|
| {if(anxietySymptoms_SQ001=="AO01", anxietySymptoms_SQ001.question)} | ○ | ○ | ○ | ○ | ○ |
| {if(anxietySymptoms_SQ002=="AO01", anxietySymptoms_SQ002.question)} | ○ | ○ | ○ | ○ | ○ |
| {if(anxietySymptoms_SQ003=="AO01", anxietySymptoms_SQ003.question)} | ○ | ○ | ○ | ○ | ○ |
| {if(anxietySymptoms_SQ004=="AO01", anxietySymptoms_SQ004.question)} | ○ | ○ | ○ | ○ | ○ |
| {if(anxietySymptoms_SQ005=="AO01", anxietySymptoms_SQ005.question)} | ○ | ○ | ○ | ○ | ○ |
| {if(anxietySymptoms_SQ006=="AO01", anxietySymptoms_SQ006.question)} | ○ | ○ | ○ | ○ | ○ |
| {if(anxietySymptoms_SQ007=="AO01", anxietySymptoms_SQ007.question)} | ○ | ○ | ○ | ○ | ○ |
| {if(anxietySymptoms_SQ008=="AO01", anxietySymptoms_SQ008.question)} | ○ | ○ | ○ | ○ | ○ |

**When you are feeling depressed, do you experience any of the following symptoms related to your memory, ability to focus and think (general cognitive funtion)? Please answer YES or NO to each of the symptoms listed.**

Please choose the appropriate response for each item:

| | Yes | No |
|---|---|---|
| Reduced ability to think and process thoughts | ◯ | ◯ |
| Everything feels like it is a burden: can't be bothered with anything | ◯ | ◯ |
| Reduced ability to concentrate on a book or movie | ◯ | ◯ |
| Reduced ability to concentrate at work/study | ◯ | ◯ |
| Reduced ability to take responsibility for every day tasks | ◯ | ◯ |
| Reduced ability to manage the workload expected of you | ◯ | ◯ |
| Racing thoughts - Your mind isn't able to "shut off" and you can't fully relax | ◯ | ◯ |
| Reduced ability to make decisions | ◯ | ◯ |
| Increased creativity | ◯ | ◯ |
| Increased ability to focus and manage the workload expected of you | ◯ | ◯ |

#### Please rate the impact of these symptoms on your day-to-day life when you are in a depressive episode

Please choose the appropriate response for each item:

| | 1 No Impact | 2 Some Impact | 3 Moderate Impact | 4 Major Impact | 5 Severe Impact |
|---|---|---|---|---|---|
| {if(cognitiveSymptoms_SQ001=="AO01", cognitiveSymptoms_SQ001.question)} | ○ | ○ | ○ | ○ | ○ |
| {if(cognitiveSymptoms_SQ002=="AO01", cognitiveSymptoms_SQ002.question)} | ○ | ○ | ○ | ○ | ○ |
| {if(cognitiveSymptoms_SQ003=="AO01", cognitiveSymptoms_SQ003.question)} | ○ | ○ | ○ | ○ | ○ |
| {if(cognitiveSymptoms_SQ004=="AO01", cognitiveSymptoms_SQ004.question)} | ○ | ○ | ○ | ○ | ○ |
| {if(cognitiveSymptoms_SQ005=="AO01", cognitiveSymptoms_SQ005.question)} | ○ | ○ | ○ | ○ | ○ |
| {if(cognitiveSymptoms_SQ006=="AO01", cognitiveSymptoms_SQ006.question)} | ○ | ○ | ○ | ○ | ○ |
| {if(cognitiveSymptoms_SQ007=="AO01", cognitiveSymptoms_SQ007.question)} | ○ | ○ | ○ | ○ | ○ |
| {if(cognitiveSymptoms_SQ008=="AO01", cognitiveSymptoms_SQ008.question)} | ○ | ○ | ○ | ○ | ○ |
| {if(cognitiveSymptoms_SQ009=="AO01", cognitiveSymptoms_SQ009.question)} | ○ | ○ | ○ | ○ | ○ |
| {if(cognitiveSymptoms_SQ010=="AO01", cognitiveSymptoms_SQ010.question)} | ○ | ○ | ○ | ○ | ○ |

**When you are feeling depressed, do you experience any of the following changes in your sleep pattern? Please answer YES or NO to each of the symptoms listed.**

Please choose the appropriate response for each item:

| | Yes | No |
|---|---|---|
| **Difficulty getting to sleep** | ○ | ○ |
| **Waking throughout the night** | ○ | ○ |
| **Waking up much later** | ○ | ○ |
| **Oversleeping (sleep more than 8-10 hours a night)** | ○ | ○ |
| **Waking up feeling unrefreshed, always feeling fatigued** | ○ | ○ |
| **Feel worst in the morning** | ○ | ○ |
| **Internal body clock seems out of sync with actual time of day – like jetlag** | ○ | ○ |
| **Early morning (before sunrise) wakening** | ○ | ○ |

#### Please rate the impact of these symptoms on your day-to-day life when you are in a depressive episode

Please choose the appropriate response for each item:

| | 1 No Impact | 2 Some Impact | 3 Moderate Impact | 4 Major Impact | 5 Severe Impact |
|---|---|---|---|---|---|
| **{if(sleepSymptoms_SQ001=="AO01", sleepSymptoms_SQ001.question)}** | ○ | ○ | ○ | ○ | ○ |
| **{if(sleepSymptoms_SQ007=="AO01", sleepSymptoms_SQ007.question)}** | ○ | ○ | ○ | ○ | ○ |
| **{if(sleepSymptoms_SQ008=="AO01", sleepSymptoms_SQ008.question)}** | ○ | ○ | ○ | ○ | ○ |
| **{if(sleepSymptoms_SQ003=="AO01", sleepSymptoms_SQ003.question)}** | ○ | ○ | ○ | ○ | ○ |
| **{if(sleepSymptoms_SQ002=="AO01", sleepSymptoms_SQ002.question)}** | ○ | ○ | ○ | ○ | ○ |
| **{if(sleepSymptoms_SQ004=="AO01", sleepSymptoms_SQ004.question)}** | ○ | ○ | ○ | ○ | ○ |
| **{if(sleepSymptoms_SQ005=="AO01", sleepSymptoms_SQ005.question)}** | ○ | ○ | ○ | ○ | ○ |
| **{if(sleepSymptoms_SQ006=="AO01", sleepSymptoms_SQ006.question)}** | ○ | ○ | ○ | ○ | ○ |

**When you are feeling depressed, do you experience any of the following changes in your behaviour? Please answer YES or NO to each of the symptoms listed.**

Please choose the appropriate response for each item:

| | Yes | No |
|---|---|---|
| **Unable to sit still, feeling the need to keep moving** | ○ | ○ |
| **Reduced daytime activity** | ○ | ○ |
| **Increased sensitivity to criticism or rejection** | ○ | ○ |
| **Spend money without thinking it through** | ○ | ○ |
| **Easily irritated or frustrated - even unprovoked rage** | ○ | ○ |
| **Alienate friends and/or family** | ○ | ○ |
| **Reduced interest in participating in social activities** | ○ | ○ |
| **Loss of interest in sex** | ○ | ○ |
| **Engage very repetitive behaviours - like unnecessary washing, cleaning or checking over and over** | ○ | ○ |
| **Cant be bothered to talk: give single word answers when asked a question** | ○ | ○ |

#### Please rate the impact of these symptoms on your day-to-day life when you are in a depressive episode

Please choose the appropriate response for each item:

| | 1 No Impact | 2 Some Impact | 3 Moderate Impact | 4 Major Impact | 5 Severe Impact |
|---|---|---|---|---|---|
| {if(behaviouralSymptoms_SQ001=="AO01", behaviouralSymptoms_SQ001.question)} | ○ | ○ | ○ | ○ | ○ |
| {if(behaviouralSymptoms_SQ009=="AO01", behaviouralSymptoms_SQ009.question)} | ○ | ○ | ○ | ○ | ○ |
| {if(behaviouralSymptoms_SQ002=="AO01", behaviouralSymptoms_SQ002.question)} | ○ | ○ | ○ | ○ | ○ |
| {if(behaviouralSymptoms_SQ003=="AO01", behaviouralSymptoms_SQ003.question)} | ○ | ○ | ○ | ○ | ○ |
| {if(behaviouralSymptoms_SQ004=="AO01", behaviouralSymptoms_SQ004.question)} | ○ | ○ | ○ | ○ | ○ |
| {if(behaviouralSymptoms_SQ005=="AO01", behaviouralSymptoms_SQ005.question)} | ○ | ○ | ○ | ○ | ○ |
| {if(behaviouralSymptoms_SQ006=="AO01", behaviouralSymptoms_SQ006.question)} | ○ | ○ | ○ | ○ | ○ |
| {if(behaviouralSymptoms_SQ007=="AO01", behaviouralSymptoms_SQ007.question)} | ○ | ○ | ○ | ○ | ○ |
| {if(behaviouralSymptoms_SQ010=="AO01", behaviouralSymptoms_SQ010.question)} | ○ | ○ | ○ | ○ | ○ |
| {if(behaviouralSymptoms_SQ008=="AO01", behaviouralSymptoms_SQ008.question)} | ○ | ○ | ○ | ○ | ○ |

**When you are feeling depressed, do you experience any of the following physical symptoms? Please answer YES or NO to each of the symptoms listed. Please answer YES or NO to each of the symptoms listed.**

Please choose the appropriate response for each item:

| | Yes | No |
|---|---|---|
| Lack of energy | ◯ | ◯ |
| Everything is in slow motion | ◯ | ◯ |
| Racing heart, sweat or have trouble breathing | ◯ | ◯ |
| Appetite increases | ◯ | ◯ |
| Appetite decreases | ◯ | ◯ |
| Significant weight gain | ◯ | ◯ |
| Significant weight loss | ◯ | ◯ |
| Heavy feeling in arms or legs | ◯ | ◯ |
| Unexplained aches and pains | ◯ | ◯ |
| Headaches and/or migraines | ◯ | ◯ |
| Gut problems e.g., constipation, peptic ulcers, reflux, irritable bowel | ◯ | ◯ |
| Low sex drive | ◯ | ◯ |
| Sexual dysfunction e.g., inability to get an erection or to reach orgasm or painful | ◯ | ◯ |
| Changes in menstrual cycle (female only, if applicable) | ◯ | ◯ |

#### Please rate the impact of these symptoms on your day-to-day life when you are in a depressive episode

Please choose the appropriate response for each item:

| | 1 No Impact | 2 Some Impact | 3 Moderate Impact | 4 Major Impact | 5 Severe Impact |
| --- | --- | --- | --- | --- | --- |
| {if(physicalSymptoms_SQ014=="AO01", physicalSymptoms_SQ014.question)} | ○ | ○ | ○ | ○ | ○ |
| {if(physicalSymptoms_SQ001=="AO01", physicalSymptoms_SQ001.question)} | ○ | ○ | ○ | ○ | ○ |
| {if(physicalSymptoms_SQ002=="AO01", physicalSymptoms_SQ002.question)} | ○ | ○ | ○ | ○ | ○ |
| {if(physicalSymptoms_SQ003=="AO01", physicalSymptoms_SQ003.question)} | ○ | ○ | ○ | ○ | ○ |
| {if(physicalSymptoms_SQ004=="AO01", physicalSymptoms_SQ004.question)} | ○ | ○ | ○ | ○ | ○ |
| {if(physicalSymptoms_SQ005=="AO01", physicalSymptoms_SQ005.question)} | ○ | ○ | ○ | ○ | ○ |
| {if(physicalSymptoms_SQ006=="AO01", physicalSymptoms_SQ006.question)} | ○ | ○ | ○ | ○ | ○ |
| {if(physicalSymptoms_SQ007=="AO01", physicalSymptoms_SQ007.question)} | ○ | ○ | ○ | ○ | ○ |
| {if(physicalSymptoms_SQ008=="AO01", physicalSymptoms_SQ008.question)} | ○ | ○ | ○ | ○ | ○ |
| {if(physicalSymptoms_SQ009=="AO01", physicalSymptoms_SQ009.question)} | ○ | ○ | ○ | ○ | ○ |
| {if(physicalSymptoms_SQ010=="AO01", physicalSymptoms_SQ010.question)} | ○ | ○ | ○ | ○ | ○ |
| {if(physicalSymptoms_SQ012=="AO01", physicalSymptoms_SQ012.question)} | ○ | ○ | ○ | ○ | ○ |
| {if(physicalSymptoms_SQ013=="AO01", physicalSymptoms_SQ013.question)} | ○ | ○ | ○ | ○ | ○ |
| {if(physicalSymptoms_SQ011=="AO01", physicalSymptoms_SQ011.question)} | ○ | ○ | ○ | ○ | ○ |

#### Do you experience challenges in maintaining your health and physical wellbeing due to symptoms of depression? Please choose any relevant options where depression impacts on your ability to manage that particular aspect of your wellbeing.

❗ Check all that apply
Please choose **all** that apply:

- ☐ Dental or oral hygiene (e.g., brushing, flossing, regular dental appointments)
- ☐ Food quality, adequate nutrition, and diet management
- ☐ Bodily hygiene (e.g., washing, hair care, regular showering or bathing, physical activity)
- ☐ Keeping a clean home or living space (e.g., daily chores)

#### Do you find that the time of year (season) impacts your depressive symptoms?

Please choose **only one** of the following:

- ◯ Yes
- ◯ No

##### If yes, at what time of year does the depression or low energy state come on?

Only answer this question if the following conditions are met:
((seasonalEffects.NAOK == "Y"))

❗ Check all that apply
Please choose **all** that apply:

- ☐ Summer
- ☐ Winter
- ☐ Autumn
- ☐ Spring

##### In later sections we will ask about how you feel between episodes of depression and about your medications, but if you wish, in your own words describe your experience when feeling depressed.

Please write your answer here:

### SECTION 2: Best Treatments for Your Depression

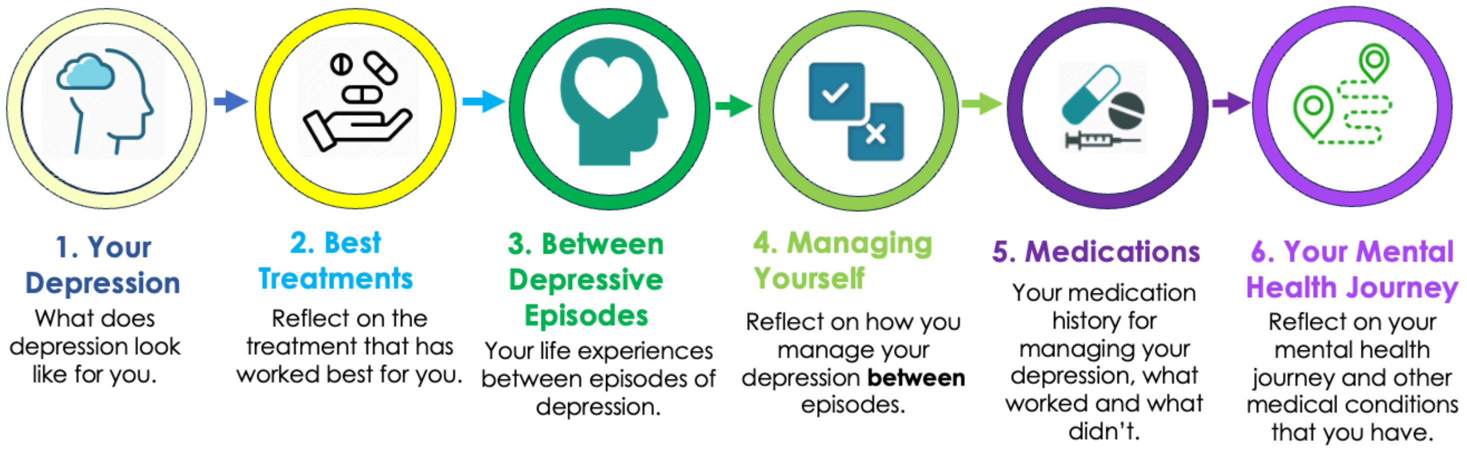

We know that the effectiveness of treatment(s) for depression is different for everyone. We are interested, from your experience, what has been **the most effective** treatment or combination of treatments in managing your depression **when you are feeling depressed**.

In later sections we will ask you about all treatments or medications you have tried. Here, we focus on the treatment that has been **most effective in relieving your symptoms** of depression. For some people, and for a variety of reasons, the best treatment for relieving symptoms may mean this treatment is not their current treatment of choice. Still, we ask you to focus on the treatment you think has been most effective with respect to symptoms of depression.

*In this section, there are 20 questions, and depending on your responses, this can increase. We anticipate that this section should take no longer than 10-12 minutes to complete.*

#### <u>In your experience, has any treatment you have tried for depression</u> provided some 'positive' change in the depression symptoms you experienced?

Please choose **only one** of the following:

- ◯ Yes
- ◯ No

#### In that case we will move on to the next section, unless you would like to tell us about your experiences with the best treatment you have tried so far

Only answer this question if the following conditions are met:
Answer was 'No' at question ' [positiveChange]' (In your experience, has any treatment you have tried for depression provided some ‘positive’ change in the depression symptoms you experienced?)

❶ Choose one of the following answers
Please choose **only one** of the following:

- ◯ Yes, I would like to answer the questions posted in this section
- ◯ No, I would like to continue to the next section

#### How would you rate the change in symptoms you experienced for the treatment that worked the best for you?

Only answer this question if the following conditions are met:
((positiveChange.NAOK == "Y") or (bestTreatmentExp.NAOK == "AO01"))

Please choose the appropriate response for each item:

| 1 None | 2 Minor | 3 Some | 4 Moderate | 5 Major | 6 Significant | 7 Extreme |
|---|---|---|---|---|---|---|
| ◯ | ◯ | ◯ | ◯ | ◯ | ◯ | ◯ |

#### In your experience, what has been the <u>most effective psychological therapy and physical treatment</u> OR <u>combination of these</u> that you have ever tried for getting out of an episode of depression?

Only answer this question if the following conditions are met:
((positiveChange.NAOK == "Y") or (bestTreatmentExp.NAOK == "AO01"))

❗ Check all that apply
Please choose **all** that apply:

- ☐ Cognitive behavioural therapy (CBT)
- ☐ Acceptance and commitment therapy (ACT)
- ☐ Interpersonal therapy (IPT)
- ☐ Dialectical behaviour therapy (DBT)
- ☐ Behavioural therapy
- ☐ Counselling for specific issues/challenges
- ☐ Online or self-management interventions (e.g. mindfulness apps like headspace)

- ☐ Physical exercise (e.g. gym, yoga)
- ☐ Meditation
- ☐ Repetitive Transcranial Magnetic Stimulation (rTMS/TMS)
- ☐ Electroconvulsive therapy (ECT)
- ☐ Light therapy
- ☐ None
- ☐ Other: ____________

#### Please rate the impact of each psychological therapies and physical treatments in getting you out of an episode of depression.

Only answer this question if the following conditions are met:
((positiveChange.NAOK == "Y") or (bestTreatmentExp.NAOK == "AO01")) and ((is_empty(mostEffectiveTherapy_SQ013.NAOK)) and ( ! is_empty(mostEffectiveTherapy)))

Please choose the appropriate response for each item:

| | 1 None | 2 Minor Impact | 3 Some Impact | 4 Moderate Impact | 5 Major Impact | 6 Significant Impact | 7 Extreme Impact |
|---|---|---|---|---|---|---|---|
| **{if(mostEffectiveTherapy_SQ001=="Y", mostEffectiveTherapy_SQ001.question)}** | ○ | ○ | ○ | ○ | ○ | ○ | ○ |
| **{if(mostEffectiveTherapy_SQ006=="Y", mostEffectiveTherapy_SQ006.question)}** | ○ | ○ | ○ | ○ | ○ | ○ | ○ |
| **{if(mostEffectiveTherapy_SQ005=="Y", mostEffectiveTherapy_SQ005.question)}** | ○ | ○ | ○ | ○ | ○ | ○ | ○ |
| **{if(mostEffectiveTherapy_SQ014=="Y", mostEffectiveTherapy_SQ014.question)}** | ○ | ○ | ○ | ○ | ○ | ○ | ○ |
| **{if(mostEffectiveTherapy_SQ003=="Y", mostEffectiveTherapy_SQ003.question)}** | ○ | ○ | ○ | ○ | ○ | ○ | ○ |
| **{if(mostEffectiveTherapy_SQ004=="Y", mostEffectiveTherapy_SQ004.question)}** | ○ | ○ | ○ | ○ | ○ | ○ | ○ |
| **{if(mostEffectiveTherapy_SQ010=="Y", mostEffectiveTherapy_SQ010.question)}** | ○ | ○ | ○ | ○ | ○ | ○ | ○ |
| **{if(mostEffectiveTherapy_SQ011=="Y", mostEffectiveTherapy_SQ011.question)}** | ○ | ○ | ○ | ○ | ○ | ○ | ○ |
| **{if(mostEffectiveTherapy_SQ012=="Y", mostEffectiveTherapy_SQ012.question)}** | ○ | ○ | ○ | ○ | ○ | ○ | ○ |
| **{if(mostEffectiveTherapy_SQ007=="Y", mostEffectiveTherapy_SQ007.question)}** | ○ | ○ | ○ | ○ | ○ | ○ | ○ |
| **{if(mostEffectiveTherapy_SQ008=="Y", mostEffectiveTherapy_SQ008.question)}** | ○ | ○ | ○ | ○ | ○ | ○ | ○ |
| **{if(mostEffectiveTherapy_SQ009=="Y", mostEffectiveTherapy_SQ009.question)}** | ○ | ○ | ○ | ○ | ○ | ○ | ○ |
| **{if(mostEffectiveTherapy_other, mostEffectiveTherapy_other)}** | ○ | ○ | ○ | ○ | ○ | ○ | ○ |

#### This image presents an overview of symptoms people may experience during depression. Think about your set of personal symptoms in each area when answering the questions below.

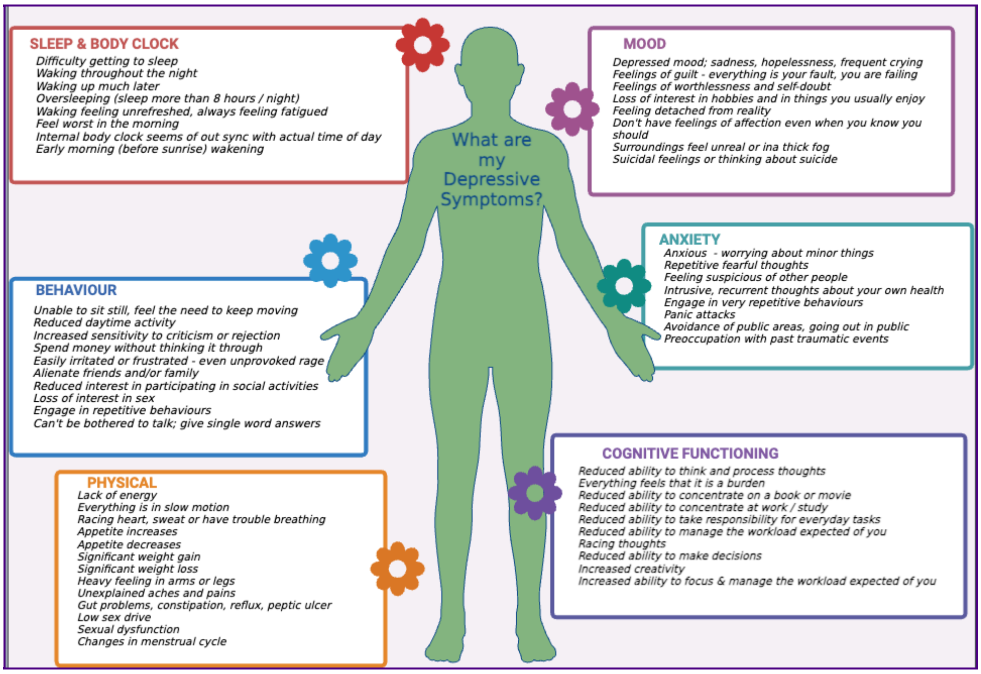

#### Please rate the impact <u>Electroconvulsive Therapy (ECT)</u> had on improving these areas of symptoms that you have reported you experience when feeling depressed.

Only answer this question if the following conditions are met:
Answer was at question ' [mostEffectiveTherapy]' (In your experience, what has been the most effective psychological therapy and physical treatment OR combination of these that you have ever tried for getting out of an episode of depression?)

Please choose the appropriate response for each item:

| | 1 None | 2 Minor Impact | 3 Some Impact | 4 Moderate Impact | 5 Major Impact | 6 Significant Impact | 7 Extreme Impact |
|---|---|---|---|---|---|---|---|
| **Mood** | ○ | ○ | ○ | ○ | ○ | ○ | ○ |
| **Anxiety** | ○ | ○ | ○ | ○ | ○ | ○ | ○ |
| **Memory, ability to focus and think** | ○ | ○ | ○ | ○ | ○ | ○ | ○ |
| **Sleep** | ○ | ○ | ○ | ○ | ○ | ○ | ○ |

| | | | | | | | |
|---|---|---|---|---|---|---|---|
| **Behaviour** | ◯ | ◯ | ◯ | ◯ | ◯ | ◯ | ◯ |
| **Physical** | ◯ | ◯ | ◯ | ◯ | ◯ | ◯ | ◯ |

#### Was Electroconvulsive Therapy (ECT) prescribed to you because you experienced any of the following symptoms,

#### 1. Psychotic Symptoms
#### 2. Suicidal Thoughts
#### 3. Refusal to eat/drink

Only answer this question if the following conditions are met:
Answer was at question ' [mostEffectiveTherapy]' (In your experience, what has been the most effective psychological therapy and physical treatment OR combination of these that you have ever tried for getting out of an episode of depression?)

Please choose **only one** of the following:

- ◯ Yes
- ◯ No

#### If you benefitted from the psychological therapies and physical treatments for getting out of an episode of depression, how quickly did that benefit come on?

Only answer this question if the following conditions are met:
((positiveChange.NAOK == "Y") or (bestTreatmentExp.NAOK == "AO01")) and (is_empty(mostEffectiveTherapy_SQ013.NAOK))

❗ Choose one of the following answers
Please choose **only one** of the following:

- ◯ Can't really specify
- ◯ Different for different episodes
- ◯ Within two weeks
- ◯ Within four weeks
- ◯ Within four to six weeks
- ◯ Greater than six weeks

#### We are now going to ask you about how successful medications have been in getting you out of an episode of depression. Here, we are interested in the most successful medication or combination of medications that have worked for you. In a later section we will ask about experiences with other medications.

Only answer this question if the following conditions are met:
((positiveChange.NAOK == "Y") or (bestTreatmentExp.NAOK == "AO01"))

#### In your experience, have you found a single medication OR combination of medications successful for getting you out of an episode of depression?

Only answer this question if the following conditions are met:
((positiveChange.NAOK == "Y") or (bestTreatmentExp.NAOK == "AO01"))

❗ Choose one of the following answers
Please choose **only one** of the following:

- ◯ Single medication
- ◯ Combination of medications
- ◯ None

**In your experience, what has been the <u>most effective 'single medication'</u> that you have ever tried for getting out of an episode of depression?**

***Hover on the ⓘ icon to see some examples for each of the options.***

Only answer this question if the following conditions are met:
Answer was 'Single medication' at question ' [Medication]' (In your experience, have you found a single medication OR combination of medications successful for getting you out of an episode of depression?)

❗ Choose one of the following answers
Please choose **only one** of the following:

- ◯ SSRIs (Selective serotonin reuptake inhibitors) ⓘ
- ◯ SNRIs (Serotonin norepinephrine reuptake inhibitors) ⓘ
- ◯ Vortioxetine
- ◯ TCAs (Tricyclic antidepressants) ⓘ
- ◯ TeCAs (Tetracyclic antidepressants) ⓘ
- ◯ MAOIs (Monoamine oxidase inhibitors) ⓘ
- ◯ Melatonin
- ◯ Agomelatine ⓘ
- ◯ Lamotrigine
- ◯ Lithium
- ◯ Antipsychotics ⓘ
- ◯ Stimulants ⓘ
- ◯ Ketamine, Esketamine
- ◯ Psychedelics
- ◯ Medicinal cannabis
- ◯ Brexanolone, Allopregnanolone
- ◯ Other ______

#### In your experience, what has been the <u>most effective “combination of medications”</u> that you have ever tried for getting out of an episode of depression?

##### *Hover on the ⓘ icon to see some examples for each of the options.*

Only answer this question if the following conditions are met:
Answer was 'Combination of medications' at question ' [Medication]' (In your experience, have you found a single medication OR combination of medications successful for getting you out of an episode of depression?)

❗ Check all that apply
❗ Please select from 2 to 4 answers.
Please choose **all** that apply:

- ☐ SSRIs (Selective serotonin reuptake inhibitors) ⓘ
- ☐ SNRIs (Serotonin norepinephrine reuptake inhibitors) ⓘ
- ☐ Vortioxetine
- ☐ TCAs (Tricyclic antidepressants) ⓘ
- ☐ TeCAs (Tetracyclic antidepressants) ⓘ
- ☐ MAOIs (Monoamine oxidase inhibitors) ⓘ
- ☐ Melatonin
- ☐ Agomelatine ⓘ
- ☐ Lamotrigine
- ☐ Lithium
- ☐ Antipsychotics ⓘ
- ☐ Stimulants ⓘ
- ☐ Ketamine, Esketamine
- ☐ Psychedelics
- ☐ Medicinal cannabis
- ☐ Brexanolone, Allopregnanolone
- ☐ Other: ____________

### SECTION 2 (cont.): Best Treatments for Your Depression

#### Please rate the impact of the medication in getting you out of an episode of depression

Only answer this question if the following conditions are met:
((positiveChange.NAOK == "Y") or (bestTreatmentExp.NAOK == "AO01")) and ((Medication.NAOK == "AO01"))

Please choose the appropriate response for each item:

| | 1 None | 2 Minor Impact | 3 Some Impact | 4 Moderate Impact | 5 Major Impact | 6 Significant Impact | 7 Extreme Impact |
|---|---|---|---|---|---|---|---|
| SSRIs (Selective serotonin reuptake inhibitors) ⓘ | ○ | ○ | ○ | ○ | ○ | ○ | ○ |
| SNRIs (Serotonin norepinephrine reuptake inhibitors) ⓘ | ○ | ○ | ○ | ○ | ○ | ○ | ○ |
| Vortioxetine | ○ | ○ | ○ | ○ | ○ | ○ | ○ |
| TCAs (Tricyclic antidepressants) ⓘ | ○ | ○ | ○ | ○ | ○ | ○ | ○ |
| TeCAs (Tetracyclic antidepressants) ⓘ | ○ | ○ | ○ | ○ | ○ | ○ | ○ |
| MAOIs (Monoamine oxidase inhibitors) ⓘ | ○ | ○ | ○ | ○ | ○ | ○ | ○ |
| Melatonin | ○ | ○ | ○ | ○ | ○ | ○ | ○ |
| Agomelatine ⓘ | ○ | ○ | ○ | ○ | ○ | ○ | ○ |
| Lamotrigine | ○ | ○ | ○ | ○ | ○ | ○ | ○ |
| Lithium | ○ | ○ | ○ | ○ | ○ | ○ | ○ |
| Antipsychotics ⓘ | ○ | ○ | ○ | ○ | ○ | ○ | ○ |
| Stimulants ⓘ | ○ | ○ | ○ | ○ | ○ | ○ | ○ |
| Ketamine, Esketamine | ○ | ○ | ○ | ○ | ○ | ○ | ○ |
| Psychedelics | ○ | ○ | ○ | ○ | ○ | ○ | ○ |

| | | | | | | | |
|---|---|---|---|---|---|---|---|
| **Medicinal cannabis** | ○ | ○ | ○ | ○ | ○ | ○ | ○ |
| **Brexanolone, Allopregnanolone** | ○ | ○ | ○ | ○ | ○ | ○ | ○ |
| **{singleMedication_other}** | ○ | ○ | ○ | ○ | ○ | ○ | ○ |

#### This image presents an overview of symptoms people may experience during depression. Think about your set of personal symptoms in each area when answering the questions below.

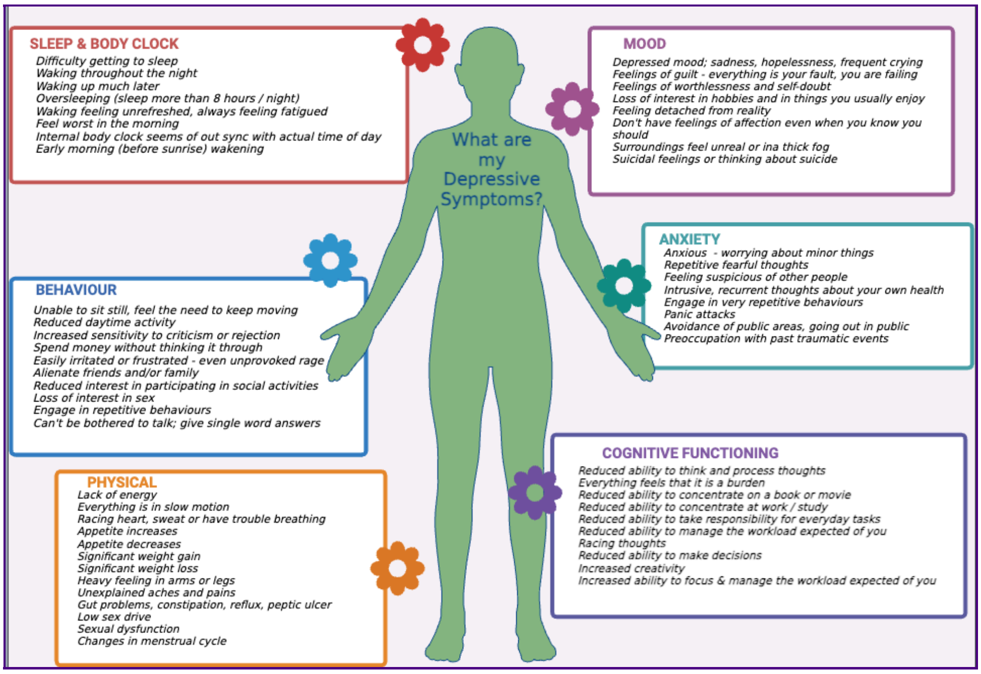

#### Please rate the impact of the medication that helped in improving the following areas of symptoms that you have reported you experience when feeling depressed?

Only answer this question if the following conditions are met:
((positiveChange.NAOK == "Y") or (bestTreatmentExp.NAOK == "AO01")) and ( ! is_empty(singleMedication))

Please choose the appropriate response for each item:

| | 1 None | 2 Minor Impact | 3 Some Impact | 4 Moderate Impact | 5 Major Impact | 6 Significant Impact | 7 Extreme Impact |
|---|---|---|---|---|---|---|---|

| |
|---|
| **Mood** |
| **Anxiety** |
| **Memory, ability to focus and think** |
| **Sleep** |
| **Behaviour** |
| **Physical** |

#### Please rate the impact of the medications in getting you out of an episode of depression

Only answer this question if the following conditions are met:
((positiveChange.NAOK == "Y") or (bestTreatmentExp.NAOK == "AO01")) and ((Medication.NAOK == "AO02"))

Please choose the appropriate response for each item:

| | 1 None | 2 Minor Impact | 3 Some Impact | 4 Moderate Impact | 5 Major Impact | 6 Significant Impact | 7 Extreme Impact |
|---|---|---|---|---|---|---|---|
| **{if(CombinationMeds_SQ001=="Y", CombinationMeds_SQ001.question)}** | | | | | | | |
| **{if(CombinationMeds_SQ002=="Y", CombinationMeds_SQ002.question)}** | | | | | | | |
| **{if(CombinationMeds_SQ016=="Y", CombinationMeds_SQ016.question)}** | | | | | | | |
| **{if(CombinationMeds_SQ003=="Y", CombinationMeds_SQ003.question)}** | | | | | | | |
| **{if(CombinationMeds_SQ004=="Y", CombinationMeds_SQ004.question)}** | | | | | | | |
| **{if(CombinationMeds_SQ005=="Y", CombinationMeds_SQ005.question)}** | | | | | | | |
| **{if(CombinationMeds_SQ006=="Y", CombinationMeds_SQ006.question)}** | | | | | | | |
| **{if(CombinationMeds_SQ007=="Y", CombinationMeds_SQ007.question)}** | | | | | | | |
| **{if(CombinationMeds_SQ008=="Y", CombinationMeds_SQ008.question)}** | | | | | | | |
| **{if(CombinationMeds_SQ009=="Y", CombinationMeds_SQ009.question)}** | | | | | | | |

| | | | | | | | |
|---|---|---|---|---|---|---|---|
| {if(CombinationMeds_SQ010=="Y", CombinationMeds_SQ010.question)} | ○ | ○ | ○ | ○ | ○ | ○ | ○ |
| {if(CombinationMeds_SQ011=="Y", CombinationMeds_SQ011.question)} | ○ | ○ | ○ | ○ | ○ | ○ | ○ |
| {if(CombinationMeds_SQ012=="Y", CombinationMeds_SQ012.question)} | ○ | ○ | ○ | ○ | ○ | ○ | ○ |
| {if(CombinationMeds_SQ013=="Y", CombinationMeds_SQ013.question)} | ○ | ○ | ○ | ○ | ○ | ○ | ○ |
| {if(CombinationMeds_SQ014=="Y", CombinationMeds_SQ014.question)} | ○ | ○ | ○ | ○ | ○ | ○ | ○ |
| {if(CombinationMeds_other, CombinationMeds_other)} | ○ | ○ | ○ | ○ | ○ | ○ | ○ |
| {if(CombinationMeds_SQ017=="Y", CombinationMeds_SQ017.question)} | ○ | ○ | ○ | ○ | ○ | ○ | ○ |

#### This image presents an overview of symptoms people may experience during depression. Think about your set of personal symptoms in each area when answering the questions below.

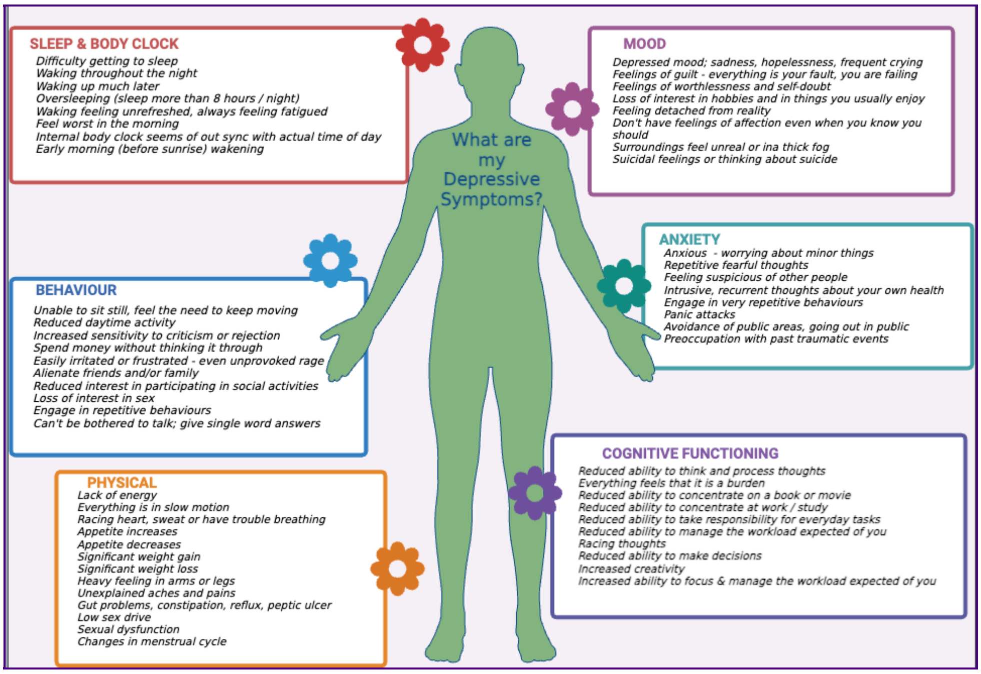

#### Please rate the impact of the following <u>medications</u> that helped in improving the following <u>areas of symptoms</u> that you have reported you experience when feeling depressed?

Only answer this question if the following conditions are met:

((positiveChange.NAOK == "Y") or (bestTreatmentExp.NAOK == "AO01")) and ( ! is_empty(CombinationMeds))

Please choose the appropriate response for each item:

| | 1 None | 2 Minor Impact | 3 Some Impact | 4 Moderate Impact | 5 Major Impact | 6 Significant Impact | 7 Extreme Impact |
|---|---|---|---|---|---|---|---|
| {if(CombinationMeds_SQ001=="Y", CombinationMeds_SQ001.question)} Mood | | | | | | | |
| {if(CombinationMeds_SQ001=="Y", CombinationMeds_SQ001.question)} Anxiety | | | | | | | |
| {if(CombinationMeds_SQ001=="Y", CombinationMeds_SQ001.question)} Memory, ability to focus and think | | | | | | | |
| {if(CombinationMeds_SQ001=="Y", CombinationMeds_SQ001.question)} Sleep | | | | | | | |
| {if(CombinationMeds_SQ001=="Y", CombinationMeds_SQ001.question)} Behaviour | | | | | | | |
| {if(CombinationMeds_SQ001=="Y", CombinationMeds_SQ001.question)} Physical | | | | | | | |
| {if(CombinationMeds_SQ002=="Y", CombinationMeds_SQ002.question)} Mood | | | | | | | |
| {if(CombinationMeds_SQ002=="Y", CombinationMeds_SQ002.question)} Anxiety | | | | | | | |
| {if(CombinationMeds_SQ002=="Y", CombinationMeds_SQ002.question)} Memory, ability to focus and think | | | | | | | |
| {if(CombinationMeds_SQ002=="Y", CombinationMeds_SQ002.question)} Sleep | | | | | | | |
| {if(CombinationMeds_SQ002=="Y", CombinationMeds_SQ002.question)} Behaviour | | | | | | | |
| {if(CombinationMeds_SQ002=="Y", CombinationMeds_SQ002.question)} Physical | | | | | | | |
| {if(CombinationMeds_SQ016=="Y", CombinationMeds_SQ016.question)} Mood | | | | | | | |

{if(CombinationMeds_SQ016=="Y", CombinationMeds_SQ016.question)} Anxiety

{if(CombinationMeds_SQ016=="Y", CombinationMeds_SQ016.question)} Memory, ability to focus and think

{if(CombinationMeds_SQ016=="Y", CombinationMeds_SQ016.question)} Sleep

{if(CombinationMeds_SQ016=="Y", CombinationMeds_SQ016.question)} Behaviour

{if(CombinationMeds_SQ016=="Y", CombinationMeds_SQ016.question)} Physical

{if(CombinationMeds_SQ003=="Y", CombinationMeds_SQ003.question)} Mood

{if(CombinationMeds_SQ003=="Y", CombinationMeds_SQ003.question)} Anxiety

{if(CombinationMeds_SQ003=="Y", CombinationMeds_SQ003.question)} Memory, ability to focus and think

{if(CombinationMeds_SQ003=="Y", CombinationMeds_SQ003.question)} Sleep

{if(CombinationMeds_SQ003=="Y", CombinationMeds_SQ003.question)} Behaviour

{if(CombinationMeds_SQ003=="Y", CombinationMeds_SQ003.question)} Physical

{if(CombinationMeds_SQ004=="Y", CombinationMeds_SQ004.question)} Mood

{if(CombinationMeds_SQ004=="Y", CombinationMeds_SQ004.question)} Anxiety

{if(CombinationMeds_SQ004=="Y", CombinationMeds_SQ004.question)} Memory, ability to focus and think

{if(CombinationMeds_SQ004=="Y", CombinationMeds_SQ004.question)} Sleep

{if(CombinationMeds_SQ004=="Y",

| | | | | | | | |
|---|---|---|---|---|---|---|---|
| CombinationMeds_SQ004.question)} Behaviour | ○ | ○ | ○ | ○ | ○ | ○ | ○ |
| {if(CombinationMeds_SQ004=="Y", CombinationMeds_SQ004.question)} Physical | ○ | ○ | ○ | ○ | ○ | ○ | ○ |
| {if(CombinationMeds_SQ005=="Y", CombinationMeds_SQ005.question)} Mood | ○ | ○ | ○ | ○ | ○ | ○ | ○ |
| {if(CombinationMeds_SQ005=="Y", CombinationMeds_SQ005.question)} Anxiety | ○ | ○ | ○ | ○ | ○ | ○ | ○ |
| {if(CombinationMeds_SQ005=="Y", CombinationMeds_SQ005.question)} Memory, ability to focus and think | ○ | ○ | ○ | ○ | ○ | ○ | ○ |
| {if(CombinationMeds_SQ005=="Y", CombinationMeds_SQ005.question)} Sleep | ○ | ○ | ○ | ○ | ○ | ○ | ○ |
| {if(CombinationMeds_SQ005=="Y", CombinationMeds_SQ005.question)} Behaviour | ○ | ○ | ○ | ○ | ○ | ○ | ○ |
| {if(CombinationMeds_SQ005=="Y", CombinationMeds_SQ005.question)} Physical | ○ | ○ | ○ | ○ | ○ | ○ | ○ |
| {if(CombinationMeds_SQ006=="Y", CombinationMeds_SQ006.question)} Mood | ○ | ○ | ○ | ○ | ○ | ○ | ○ |
| {if(CombinationMeds_SQ006=="Y", CombinationMeds_SQ006.question)} Anxiety | ○ | ○ | ○ | ○ | ○ | ○ | ○ |
| {if(CombinationMeds_SQ006=="Y", CombinationMeds_SQ006.question)} Memory, ability to focus and think | ○ | ○ | ○ | ○ | ○ | ○ | ○ |
| {if(CombinationMeds_SQ006=="Y", CombinationMeds_SQ006.question)} Sleep | ○ | ○ | ○ | ○ | ○ | ○ | ○ |
| {if(CombinationMeds_SQ006=="Y", CombinationMeds_SQ006.question)} Behaviour | ○ | ○ | ○ | ○ | ○ | ○ | ○ |
| {if(CombinationMeds_SQ006=="Y", CombinationMeds_SQ006.question)} Physical | ○ | ○ | ○ | ○ | ○ | ○ | ○ |
| {if(CombinationMeds_SQ007=="Y", CombinationMeds_SQ007.question)} Mood | ○ | ○ | ○ | ○ | ○ | ○ | ○ |
| {if(CombinationMeds_SQ007=="Y", CombinationMeds_SQ007.question)} Anxiety | ○ | ○ | ○ | ○ | ○ | ○ | ○ |

| | | | | | | |
|---|---|---|---|---|---|---|
| {if(CombinationMeds_SQ007=="Y", CombinationMeds_SQ007.question)} Memory, ability to focus and think | ○ | ○ | ○ | ○ | ○ | ○ | ○ |
| {if(CombinationMeds_SQ007=="Y", CombinationMeds_SQ007.question)} Sleep | ○ | ○ | ○ | ○ | ○ | ○ | ○ |
| {if(CombinationMeds_SQ007=="Y", CombinationMeds_SQ007.question)} Behaviour | ○ | ○ | ○ | ○ | ○ | ○ | ○ |
| {if(CombinationMeds_SQ007=="Y", CombinationMeds_SQ007.question)} Physical | ○ | ○ | ○ | ○ | ○ | ○ | ○ |
| {if(CombinationMeds_SQ008=="Y", CombinationMeds_SQ008.question)} Mood | ○ | ○ | ○ | ○ | ○ | ○ | ○ |
| {if(CombinationMeds_SQ008=="Y", CombinationMeds_SQ008.question)} Anxiety | ○ | ○ | ○ | ○ | ○ | ○ | ○ |
| {if(CombinationMeds_SQ008=="Y", CombinationMeds_SQ008.question)} Memory, ability to focus and think | ○ | ○ | ○ | ○ | ○ | ○ | ○ |
| {if(CombinationMeds_SQ008=="Y", CombinationMeds_SQ008.question)} Sleep | ○ | ○ | ○ | ○ | ○ | ○ | ○ |
| {if(CombinationMeds_SQ008=="Y", CombinationMeds_SQ008.question)} Behaviour | ○ | ○ | ○ | ○ | ○ | ○ | ○ |
| {if(CombinationMeds_SQ008=="Y", CombinationMeds_SQ008.question)} Physical | ○ | ○ | ○ | ○ | ○ | ○ | ○ |
| {if(CombinationMeds_SQ009=="Y", CombinationMeds_SQ009.question)} Mood | ○ | ○ | ○ | ○ | ○ | ○ | ○ |
| {if(CombinationMeds_SQ009=="Y", CombinationMeds_SQ009.question)} Anxiety | ○ | ○ | ○ | ○ | ○ | ○ | ○ |
| {if(CombinationMeds_SQ009=="Y", CombinationMeds_SQ009.question)} Memory, ability to focus and think | ○ | ○ | ○ | ○ | ○ | ○ | ○ |
| {if(CombinationMeds_SQ009=="Y", CombinationMeds_SQ009.question)} Sleep | ○ | ○ | ○ | ○ | ○ | ○ | ○ |
| {if(CombinationMeds_SQ009=="Y", CombinationMeds_SQ009.question)} Behaviour | ○ | ○ | ○ | ○ | ○ | ○ | ○ |
| {if(CombinationMeds_SQ009=="Y", CombinationMeds_SQ009.question)} | | | | | | | |

| | | | | | | | |
|---|---|---|---|---|---|---|---|
| Physical | ○ | ○ | ○ | ○ | ○ | ○ | ○ |
| {if(CombinationMeds_SQ010=="Y", CombinationMeds_SQ010.question)} Mood | ○ | ○ | ○ | ○ | ○ | ○ | ○ |
| {if(CombinationMeds_SQ010=="Y", CombinationMeds_SQ010.question)} Anxiety | ○ | ○ | ○ | ○ | ○ | ○ | ○ |
| {if(CombinationMeds_SQ010=="Y", CombinationMeds_SQ010.question)} Memory, ability to focus and think | ○ | ○ | ○ | ○ | ○ | ○ | ○ |
| {if(CombinationMeds_SQ010=="Y", CombinationMeds_SQ010.question)} Sleep | ○ | ○ | ○ | ○ | ○ | ○ | ○ |
| {if(CombinationMeds_SQ010=="Y", CombinationMeds_SQ010.question)} Behaviour | ○ | ○ | ○ | ○ | ○ | ○ | ○ |
| {if(CombinationMeds_SQ010=="Y", CombinationMeds_SQ010.question)} Physical | ○ | ○ | ○ | ○ | ○ | ○ | ○ |
| {if(CombinationMeds_SQ011=="Y", CombinationMeds_SQ011.question)} Mood | ○ | ○ | ○ | ○ | ○ | ○ | ○ |
| {if(CombinationMeds_SQ011=="Y", CombinationMeds_SQ011.question)} Anxiety | ○ | ○ | ○ | ○ | ○ | ○ | ○ |
| {if(CombinationMeds_SQ011=="Y", CombinationMeds_SQ011.question)} Memory, ability to focus and think | ○ | ○ | ○ | ○ | ○ | ○ | ○ |
| {if(CombinationMeds_SQ011=="Y", CombinationMeds_SQ011.question)} Sleep | ○ | ○ | ○ | ○ | ○ | ○ | ○ |
| {if(CombinationMeds_SQ011=="Y", CombinationMeds_SQ011.question)} Behaviour | ○ | ○ | ○ | ○ | ○ | ○ | ○ |
| {if(CombinationMeds_SQ011=="Y", CombinationMeds_SQ011.question)} Physical | ○ | ○ | ○ | ○ | ○ | ○ | ○ |
| {if(CombinationMeds_SQ012=="Y", CombinationMeds_SQ012.question)} Mood | ○ | ○ | ○ | ○ | ○ | ○ | ○ |
| {if(CombinationMeds_SQ012=="Y", CombinationMeds_SQ012.question)} Anxiety | ○ | ○ | ○ | ○ | ○ | ○ | ○ |
| {if(CombinationMeds_SQ012=="Y", CombinationMeds_SQ012.question)} Memory, ability to focus | ○ | ○ | ○ | ○ | ○ | ○ | ○ |

and think

{if(CombinationMeds_SQ012=="Y", CombinationMeds_SQ012.question)}
Sleep

{if(CombinationMeds_SQ012=="Y", CombinationMeds_SQ012.question)}
Behaviour

{if(CombinationMeds_SQ012=="Y", CombinationMeds_SQ012.question)}
Physical

{if(CombinationMeds_SQ013=="Y", CombinationMeds_SQ013.question)}
Mood

{if(CombinationMeds_SQ013=="Y", CombinationMeds_SQ013.question)}
Anxiety

{if(CombinationMeds_SQ013=="Y", CombinationMeds_SQ013.question)}
Memory, ability to focus and think

{if(CombinationMeds_SQ013=="Y", CombinationMeds_SQ013.question)}
Sleep

{if(CombinationMeds_SQ013=="Y", CombinationMeds_SQ013.question)}
Behaviour

{if(CombinationMeds_SQ013=="Y", CombinationMeds_SQ013.question)}
Physical

{if(CombinationMeds_SQ014=="Y", CombinationMeds_SQ014.question)}
Mood

{if(CombinationMeds_SQ014=="Y", CombinationMeds_SQ014.question)}
Anxiety

{if(CombinationMeds_SQ014=="Y", CombinationMeds_SQ014.question)}
Memory, ability to focus and think

{if(CombinationMeds_SQ014=="Y", CombinationMeds_SQ014.question)}
Sleep

{if(CombinationMeds_SQ014=="Y", CombinationMeds_SQ014.question)}
Behaviour

{if(CombinationMeds_SQ014=="Y", CombinationMeds_SQ014.question)}
Physical

{if(CombinationMeds_other,

| | | | | | | | |
|---|---|---|---|---|---|---|---|
| CombinationMeds_other)} Mood | ◯ | ◯ | ◯ | ◯ | ◯ | ◯ | ◯ |
| {if(CombinationMeds_other, CombinationMeds_other)} Anxiety | ◯ | ◯ | ◯ | ◯ | ◯ | ◯ | ◯ |
| {if(CombinationMeds_other, CombinationMeds_other)} Memory, ability to focus and think | ◯ | ◯ | ◯ | ◯ | ◯ | ◯ | ◯ |
| {if(CombinationMeds_other, CombinationMeds_other)} Sleep | ◯ | ◯ | ◯ | ◯ | ◯ | ◯ | ◯ |
| {if(CombinationMeds_other, CombinationMeds_other)} Behaviour | ◯ | ◯ | ◯ | ◯ | ◯ | ◯ | ◯ |
| {if(CombinationMeds_other, CombinationMeds_other)} Physical | ◯ | ◯ | ◯ | ◯ | ◯ | ◯ | ◯ |
| {if(CombinationMeds_SQ017=="Y", CombinationMeds_SQ017.question)} Mood | ◯ | ◯ | ◯ | ◯ | ◯ | ◯ | ◯ |
| {if(CombinationMeds_SQ017=="Y", CombinationMeds_SQ017.question)} Anxiety | ◯ | ◯ | ◯ | ◯ | ◯ | ◯ | ◯ |
| {if(CombinationMeds_SQ017=="Y", CombinationMeds_SQ017.question)} Memory, ability to focus and think | ◯ | ◯ | ◯ | ◯ | ◯ | ◯ | ◯ |
| {if(CombinationMeds_SQ017=="Y", CombinationMeds_SQ017.question)} Sleep | ◯ | ◯ | ◯ | ◯ | ◯ | ◯ | ◯ |
| {if(CombinationMeds_SQ017=="Y", CombinationMeds_SQ017.question)} Behaviour | ◯ | ◯ | ◯ | ◯ | ◯ | ◯ | ◯ |
| {if(CombinationMeds_SQ017=="Y", CombinationMeds_SQ017.question)} Physical | ◯ | ◯ | ◯ | ◯ | ◯ | ◯ | ◯ |

#### If you benefitted from the most effective medication or medication combination for getting out of an episode of depression, how quickly did that benefit come on?

Only answer this question if the following conditions are met:
((positiveChange.NAOK == "Y") or (bestTreatmentExp.NAOK == "AO01")) and (is_empty(Medication.NAOK == "AO03"))

❗ Choose one of the following answers
Please choose **only one** of the following:

- ◯ Can't really specify
- ◯ Different at different times
- ◯ Within two weeks
- ◯ Within four weeks
- ◯ Within four to six weeks
- ◯ Greater than six weeks

#### Do you experience any side effects from the most effective medications for getting out of an episode of depression?

Only answer this question if the following conditions are met:
((positiveChange.NAOK == "Y") or (bestTreatmentExp.NAOK == "AO01")) and (is_empty(Medication.NAOK == "AO03"))

Please choose **only one** of the following:

- ◯ Yes
- ◯ No

#### Can you please describe the side-effects you experienced?

Only answer this question if the following conditions are met:
((medSideEffects.NAOK == "Y"))

❗ Comment only when you choose an answer.
Please choose all that apply and provide a comment:

- ☐ Weight gain
- ☐ Nausea
- ☐ Dizziness
- ☐ Headaches
- ☐ Sleepy, zombie-like
- ☐ Loss of interest in sex
- ☐ Diarrhoea
- ☐ Other gut problems
- ☐ Other side effects

Please use the free text book to give more details about any side effects

#### When feeling depressed have you used any of the following substances to try and manage your anxiety, depression or related mental health problems?

Only answer this question if the following conditions are met:
((positiveChange.NAOK == "Y") or (bestTreatmentExp.NAOK == "AO01"))

Please choose the appropriate response for each item:

| | Yes | No |
|---|---|---|
| **Alcohol** | ◯ | ◯ |
| **Tobacco** | ◯ | ◯ |
| **Cannabis** | ◯ | ◯ |
| **Stimulants** | ◯ | ◯ |
| **Other Substances** | ◯ | ◯ |

#### Please describe what other substances you have tried. Do not include substances that have been prescribed by a medical professional.

Only answer this question if the following conditions are met:
((substancesUsage_SQ002.NAOK == "AO01"))

Please write your answer here:

❶ Check all that apply
Please choose **all** that apply:

- ☐ Psychological therapy
- ☐ Physical Treatment
- ☐ Medications

#### If you are able to rank the order of the successful therapy, or physical treatment, or medication that helped in getting you out of the depression, then add the ranking.

Only answer this question if the following conditions are met:
((positiveChange.NAOK == "Y") or (bestTreatmentExp.NAOK == "AO01")) and ((is_empty(mostEffectiveTherapy_SQ013.NAOK)) or (Medication.NAOK == "AO01" or Medication.NAOK == "AO02"))

❶ Please select at most 3 answers
Please number each box in order of preference from 1 to 3

- ☐ Psychological therapy
- ☐ Physical treatment
- ☐ Medications

Double-click or drag-and-drop items in the left list to move them to the right - your highest ranking item should be on the top right, moving through to your lowest ranking item.

**If you wish, in your own words describe your experience with what treatment/therapies/medications were effective for you.**

Only answer this question if the following conditions are met:
((positiveChange.NAOK == "Y") or (bestTreatmentExp.NAOK == "AO01"))

Please write your answer here:

### SECTION 3: Your Mental Health Between Depressive Episodes: periods when you feel your best

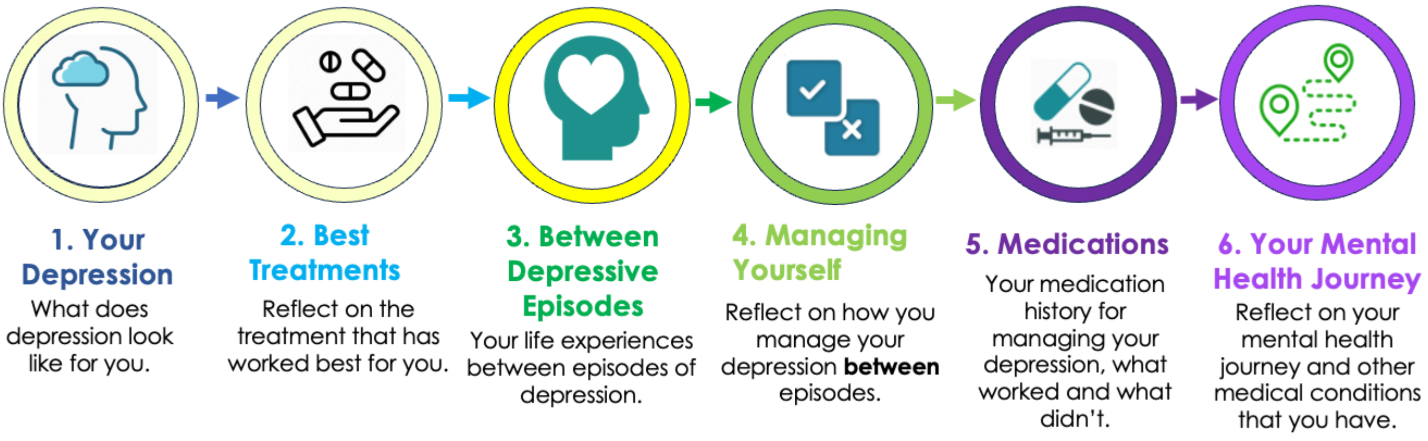

We would now like you to think about how you manage your mental health **between episodes of depression**, <u>or periods when you are feeling your best</u>. Again we know this will be different for different people and by collecting this information it will help us understand how you keep yourself well and prevent relapse.

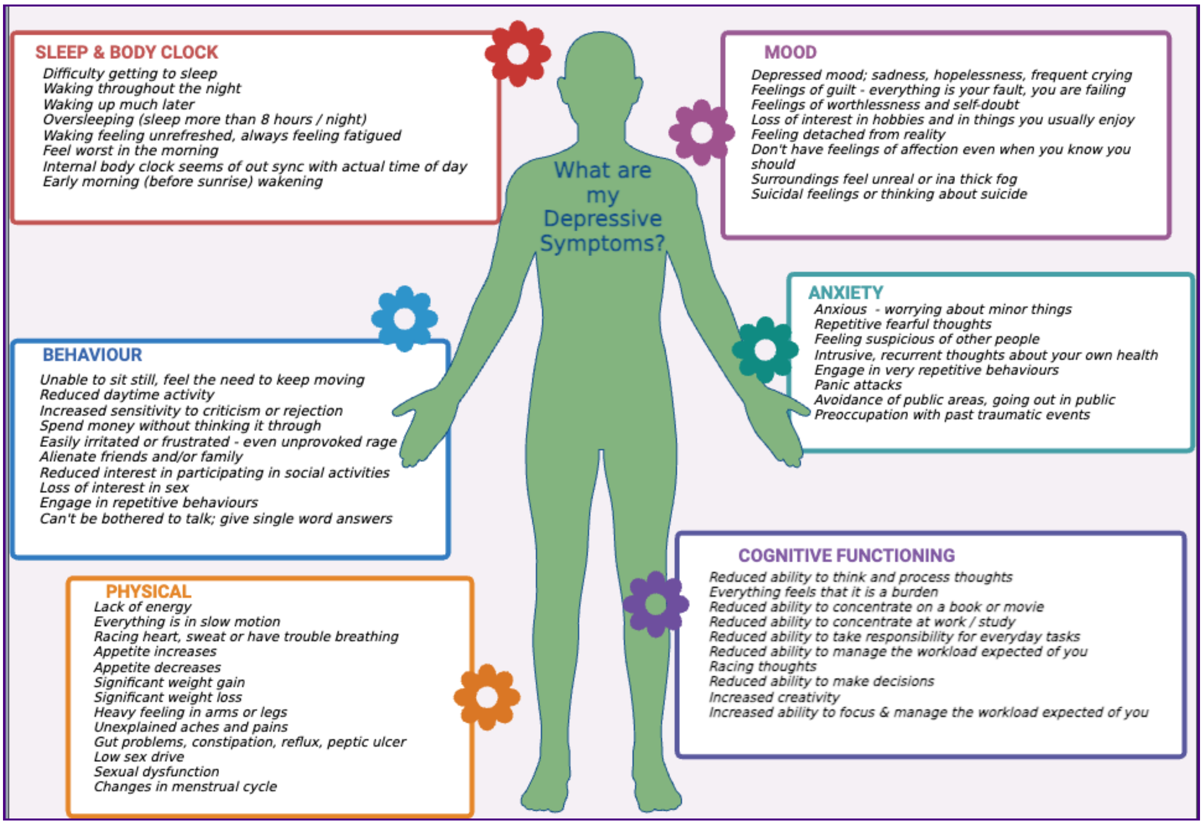

We will be asking about the same six categories of symptoms associated with depression. This section will ask you questions about any depressive symptoms you may experience **between episodes of depression** and how they may interfere with your everyday life.

*In this section, there are 10 questions, and depending on your responses, this can increase. We anticipate that this section should take no longer than 10 minutes to complete.*

#### How well do you think you are able to answer questions about depression symptoms you experience when you are well or in a period of being well-managed?

❗ Choose one of the following answers
Please choose **only one** of the following:

- ◯ Not good
- ◯ I am OK at this
- ◯ I know myself well

#### When you are between episodes of depression, do you experience any of the following symptoms related to your mood? Please answer YES or NO to each of the symptoms listed.

Please choose the appropriate response for each item:

| | Yes | No |
|---|---|---|
| **Depressed Mood: sadness, hopelessness, frequent crying** | ○ | ○ |
| **Feelings of Guilt: that everything is your fault and you are failing** | ○ | ○ |
| **Feelings of worthlessness and self-doubt** | ○ | ○ |
| **Loss of interest in hobbies and in things you usually enjoyed** | ○ | ○ |
| **Feeling detached from reality** | ○ | ○ |
| **Don’t have feelings of affection even when you know you should** | ○ | ○ |
| **Surroundings feel unreal or in a thick fog** | ○ | ○ |
| **Suicidal feelings or thinking about suicide** | ○ | ○ |

#### Please rate the impact of these symptoms on your day-to-day life when you are between episodes of depression

Please choose the appropriate response for each item:

| | 1<br>No Impact | 2<br>Some Impact | 3<br>Moderate Impact | 4<br>Major Impact | 5<br>Severe Impact |
|---|---|---|---|---|---|
| **{if(moodSymptoms1_SQ001=="AO01", moodSymptoms1_SQ001.question)}** | ◯ | ◯ | ◯ | ◯ | ◯ |
| **{if(moodSymptoms1_SQ002=="AO01", moodSymptoms1_SQ002.question)}** | ◯ | ◯ | ◯ | ◯ | ◯ |
| **{if(moodSymptoms1_SQ003=="AO01", moodSymptoms1_SQ003.question)}** | ◯ | ◯ | ◯ | ◯ | ◯ |
| **{if(moodSymptoms1_SQ004=="AO01", moodSymptoms1_SQ004.question)}** | ◯ | ◯ | ◯ | ◯ | ◯ |
| **{if(moodSymptoms1_SQ006=="AO01", moodSymptoms1_SQ006.question)}** | ◯ | ◯ | ◯ | ◯ | ◯ |
| **{if(moodSymptoms1_SQ007=="AO01", moodSymptoms1_SQ007.question)}** | ◯ | ◯ | ◯ | ◯ | ◯ |
| **{if(moodSymptoms1_SQ008=="AO01", moodSymptoms1_SQ008.question)}** | ◯ | ◯ | ◯ | ◯ | ◯ |
| **{if(moodSymptoms1_SQ005=="AO01", moodSymptoms1_SQ005.question)}** | ◯ | ◯ | ◯ | ◯ | ◯ |

**When you are between episodes of depression, do you experience any of the following symptoms related to anxiety? Please answer YES or NO to each of the symptoms listed.**

Please choose the appropriate response for each item:

| | Yes | No |
|---|---|---|
| **Anxious - worrying about minor things** | ◯ | ◯ |
| **Repetitive fearful thoughts - like being afraid of catching an infection, or that a disaster might happen** | ◯ | ◯ |
| **Feeling suspicious of other people** | ◯ | ◯ |
| **Intrusive, recurrent thoughts about your own health** | ◯ | ◯ |
| **Engagement in very repetitive behaviours – like unnecessary washing, cleaning, or checking things over** | ◯ | ◯ |
| **Panic attacks** | ◯ | ◯ |
| **Avoidance of public areas going out in public or travelling outside your home** | ◯ | ◯ |
| **Preoccupation with past traumatic events** | ◯ | ◯ |

#### Please rate the impact of these symptoms on your day-to-day life when you are between episodes of depression

Please choose the appropriate response for each item:

| | 1 No Impact | 2 Some Impact | 3 Moderate Impact | 4 Major Impact | 5 Severe Impact |
|---|---|---|---|---|---|
| {if(anxietySymptoms1_SQ001=="AO01", anxietySymptoms1_SQ001.question)} | ◯ | ◯ | ◯ | ◯ | ◯ |
| {if(anxietySymptoms1_SQ002=="AO01", anxietySymptoms1_SQ002.question)} | ◯ | ◯ | ◯ | ◯ | ◯ |
| {if(anxietySymptoms1_SQ003=="AO01", anxietySymptoms1_SQ003.question)} | ◯ | ◯ | ◯ | ◯ | ◯ |
| {if(anxietySymptoms1_SQ004=="AO01", anxietySymptoms1_SQ004.question)} | ◯ | ◯ | ◯ | ◯ | ◯ |
| {if(anxietySymptoms1_SQ005=="AO01", anxietySymptoms1_SQ005.question)} | ◯ | ◯ | ◯ | ◯ | ◯ |
| {if(anxietySymptoms1_SQ006=="AO01", anxietySymptoms1_SQ006.question)} | ◯ | ◯ | ◯ | ◯ | ◯ |
| {if(anxietySymptoms1_SQ007=="AO01", anxietySymptoms1_SQ007.question)} | ◯ | ◯ | ◯ | ◯ | ◯ |
| {if(anxietySymptoms1_SQ008=="AO01", anxietySymptoms1_SQ008.question)} | ◯ | ◯ | ◯ | ◯ | ◯ |

**When you are between episodes of depression, do you experience any of the following symptoms related to your memory, ability to focus and think (general cognitive funtion)? Please answer YES or NO to each of the symptoms listed.**

Please choose the appropriate response for each item:

| | Yes | No |
|---|---|---|
| Lack of ability to think and process thoughts | ◯ | ◯ |
| Everything feels like it is a burden: can't be bothered with anything | ◯ | ◯ |
| Lack of ability to concentrate on a book or movie | ◯ | ◯ |
| Lack of ability to concentrate at work/study | ◯ | ◯ |
| Lack of ability to take responsibility for every day tasks | ◯ | ◯ |
| Lack of ability to manage the workload expected of you | ◯ | ◯ |
| Racing thoughts - Your mind isn't able to "shut off" and you can't fully relax | ◯ | ◯ |
| Lack of ability to make decisions | ◯ | ◯ |
| Overly creative | ◯ | ◯ |
| Overly able to focus and manage the workload expected of you | ◯ | ◯ |

#### Please rate the impact of these symptoms on your day-to-day life when you are between episodes of depression

Please choose the appropriate response for each item:

| | 1 No Impact | 2 Some Impact | 3 Moderate Impact | 4 Major Impact | 5 Severe Impact |
|---|---|---|---|---|---|
| **{if(cognitiveSymptoms1_SQ001=="AO01", cognitiveSymptoms1_SQ001.question)}** | ○ | ○ | ○ | ○ | ○ |
| **{if(cognitiveSymptoms1_SQ002=="AO01", cognitiveSymptoms1_SQ002.question)}** | ○ | ○ | ○ | ○ | ○ |
| **{if(cognitiveSymptoms1_SQ003=="AO01", cognitiveSymptoms1_SQ003.question)}** | ○ | ○ | ○ | ○ | ○ |
| **{if(cognitiveSymptoms1_SQ004=="AO01", cognitiveSymptoms1_SQ004.question)}** | ○ | ○ | ○ | ○ | ○ |
| **{if(cognitiveSymptoms1_SQ005=="AO01", cognitiveSymptoms1_SQ005.question)}** | ○ | ○ | ○ | ○ | ○ |
| **{if(cognitiveSymptoms1_SQ006=="AO01", cognitiveSymptoms1_SQ006.question)}** | ○ | ○ | ○ | ○ | ○ |
| **{if(cognitiveSymptoms1_SQ007=="AO01", cognitiveSymptoms1_SQ007.question)}** | ○ | ○ | ○ | ○ | ○ |
| **{if(cognitiveSymptoms1_SQ008=="AO01", cognitiveSymptoms1_SQ008.question)}** | ○ | ○ | ○ | ○ | ○ |
| **{if(cognitiveSymptoms1_SQ009=="AO01", cognitiveSymptoms1_SQ009.question)}** | ○ | ○ | ○ | ○ | ○ |
| **{if(cognitiveSymptoms1_SQ010=="AO01", cognitiveSymptoms1_SQ010.question)}** | ○ | ○ | ○ | ○ | ○ |

**When you are between episodes of depression, do you experience any of the following changes in your sleep pattern or body clock? Please answer YES or NO to each of the symptoms listed.**

Please choose the appropriate response for each item:

| | Yes | No |
|---|---|---|
| **Difficulty getting to sleep** | ○ | ○ |
| **Waking throughout the night** | ○ | ○ |
| **Waking up much later** | ○ | ○ |
| **Oversleeping (sleep more than 8-10 hours a night)** | ○ | ○ |
| **Waking up feeling unrefreshed, always feeling fatigued** | ○ | ○ |
| **Feel worst in the morning** | ○ | ○ |
| **Internal body clock seems out of sync with actual time of day – like jetlag** | ○ | ○ |
| **Early morning (before sunrise) wakening** | ○ | ○ |

#### Please rate the impact of these symptoms on your day-to-day life when you are between episodes of depression

Please choose the appropriate response for each item:

| | 1 No Impact | 2 Some Impact | 3 Moderate Impact | 4 Major Impact | 5 Severe Impact |
|---|---|---|---|---|---|
| {if(sleepSymptoms1_SQ001=="AO01", sleepSymptoms1_SQ001.question)} | ○ | ○ | ○ | ○ | ○ |
| {if(sleepSymptoms1_SQ007=="AO01", sleepSymptoms1_SQ007.question)} | ○ | ○ | ○ | ○ | ○ |
| {if(sleepSymptoms1_SQ008=="AO01", sleepSymptoms1_SQ008.question)} | ○ | ○ | ○ | ○ | ○ |
| {if(sleepSymptoms1_SQ003=="AO01", sleepSymptoms1_SQ003.question)} | ○ | ○ | ○ | ○ | ○ |
| {if(sleepSymptoms1_SQ002=="AO01", sleepSymptoms1_SQ002.question)} | ○ | ○ | ○ | ○ | ○ |
| {if(sleepSymptoms1_SQ004=="AO01", sleepSymptoms1_SQ004.question)} | ○ | ○ | ○ | ○ | ○ |
| {if(sleepSymptoms1_SQ005=="AO01", sleepSymptoms1_SQ005.question)} | ○ | ○ | ○ | ○ | ○ |
| {if(sleepSymptoms1_SQ006=="AO01", sleepSymptoms1_SQ006.question)} | ○ | ○ | ○ | ○ | ○ |

**When you are between episodes of depression, do you experience any of the following changes in your behaviour? Please answer YES or NO to each of the symptoms listed.**

Please choose the appropriate response for each item:

| | Yes | No |
|---|---|---|
| Unable to sit still, feeling the need to keep moving | ○ | ○ |
| Lack of daytime activity | ○ | ○ |
| Overly sensitive to criticism or rejection | ○ | ○ |
| Spend money without thinking it through | ○ | ○ |
| Easily irritated or frustrated - even unprovoked rage | ○ | ○ |
| Alienate friends and/or family | ○ | ○ |
| Lack of interest in participating in social activities | ○ | ○ |
| Lack of interest in sex | ○ | ○ |
| Engage very repetitive behaviours - like unnecessary washing, cleaning or checking over and over | ○ | ○ |
| Cant be bothered to talk: give single word answers when asked a question | ○ | ○ |

#### Please rate the impact of these symptoms on your day-to-day life when you are between episodes of depression

Please choose the appropriate response for each item:

| | 1<br>No Impact | 2<br>Some Impact | 3<br>Moderate Impact | 4<br>Major Impact | 5<br>Severe Impact |
|---|---|---|---|---|---|
| {if(behaviouralSymptoms1_SQ001=="AO01", behaviouralSymptoms1_SQ001.question)} | ○ | ○ | ○ | ○ | ○ |
| {if(behaviouralSymptoms1_SQ009=="AO01", behaviouralSymptoms1_SQ009.question)} | ○ | ○ | ○ | ○ | ○ |
| {if(behaviouralSymptoms1_SQ002=="AO01", behaviouralSymptoms1_SQ002.question)} | ○ | ○ | ○ | ○ | ○ |
| {if(behaviouralSymptoms1_SQ003=="AO01", behaviouralSymptoms1_SQ003.question)} | ○ | ○ | ○ | ○ | ○ |
| {if(behaviouralSymptoms1_SQ004=="AO01", behaviouralSymptoms1_SQ004.question)} | ○ | ○ | ○ | ○ | ○ |
| {if(behaviouralSymptoms1_SQ005=="AO01", behaviouralSymptoms1_SQ005.question)} | ○ | ○ | ○ | ○ | ○ |
| {if(behaviouralSymptoms1_SQ006=="AO01", behaviouralSymptoms1_SQ006.question)} | ○ | ○ | ○ | ○ | ○ |
| {if(behaviouralSymptoms1_SQ007=="AO01", behaviouralSymptoms1_SQ007.question)} | ○ | ○ | ○ | ○ | ○ |
| {if(behaviouralSymptoms1_SQ010=="AO01", behaviouralSymptoms1_SQ010.question)} | ○ | ○ | ○ | ○ | ○ |
| {if(behaviouralSymptoms1_SQ008=="AO01", behaviouralSymptoms1_SQ008.question)} | ○ | ○ | ○ | ○ | ○ |

#### When you are between episodes of depression, do you experience any of the following physical symptoms? Please answer YES or NO to each of the symptoms listed. Please answer YES or NO to each of the symptoms listed.

Please choose the appropriate response for each item:

| | Yes | No |
|---|---|---|
| Lack of energy | ○ | ○ |
| Everything is in slow motion | ○ | ○ |
| Racing heart, sweat or have trouble breathing | ○ | ○ |
| Appetite increases | ○ | ○ |
| Appetite decreases | ○ | ○ |
| Significant weight gain | ○ | ○ |
| Significant weight loss | ○ | ○ |
| Heavy feeling in arms or legs | ○ | ○ |
| Unexplained aches and pains | ○ | ○ |
| Headaches and/or migraines | ○ | ○ |
| Gut problems e.g., constipation, peptic ulcers, reflux, irritable bowel | ○ | ○ |
| Low sex drive | ○ | ○ |
| Sexual dysfunction e.g., inability to get an erection or to reach orgasm or painful | ○ | ○ |
| Changes in menstrual cycle (female only, if applicable) | ○ | ○ |

#### Please rate the impact of these symptoms on your day-to-day life when you are between episodes of depression

Please choose the appropriate response for each item:

| | 1 No Impact | 2 Some Impact | 3 Moderate Impact | 4 Major Impact | 5 Severe Impact |
|---|---|---|---|---|---|
| {if(physicalSymptoms1_SQ014=="AO01", physicalSymptoms1_SQ014.question)} | ○ | ○ | ○ | ○ | ○ |
| {if(physicalSymptoms1_SQ001=="AO01", physicalSymptoms1_SQ001.question)} | ○ | ○ | ○ | ○ | ○ |
| {if(physicalSymptoms1_SQ002=="AO01", physicalSymptoms1_SQ002.question)} | ○ | ○ | ○ | ○ | ○ |
| {if(physicalSymptoms1_SQ003=="AO01", physicalSymptoms1_SQ003.question)} | ○ | ○ | ○ | ○ | ○ |
| {if(physicalSymptoms1_SQ004=="AO01", physicalSymptoms1_SQ004.question)} | ○ | ○ | ○ | ○ | ○ |
| {if(physicalSymptoms1_SQ005=="AO01", physicalSymptoms1_SQ005.question)} | ○ | ○ | ○ | ○ | ○ |
| {if(physicalSymptoms1_SQ006=="AO01", physicalSymptoms1_SQ006.question)} | ○ | ○ | ○ | ○ | ○ |
| {if(physicalSymptoms1_SQ007=="AO01", physicalSymptoms1_SQ007.question)} | ○ | ○ | ○ | ○ | ○ |
| {if(physicalSymptoms1_SQ008=="AO01", physicalSymptoms1_SQ008.question)} | ○ | ○ | ○ | ○ | ○ |
| {if(physicalSymptoms1_SQ009=="AO01", physicalSymptoms1_SQ009.question)} | ○ | ○ | ○ | ○ | ○ |
| {if(physicalSymptoms1_SQ010=="AO01", physicalSymptoms1_SQ010.question)} | ○ | ○ | ○ | ○ | ○ |
| {if(physicalSymptoms1_SQ012=="AO01", physicalSymptoms1_SQ012.question)} | ○ | ○ | ○ | ○ | ○ |
| {if(physicalSymptoms1_SQ013=="AO01", physicalSymptoms1_SQ013.question)} | ○ | ○ | ○ | ○ | ○ |
| {if(physicalSymptoms1_SQ011=="AO01", physicalSymptoms1_SQ011.question)} | ○ | ○ | ○ | ○ | ○ |

**Please describe any other symptoms you may experience when you are between episodes of depression that were not included in the lists above.**

Please write your answer here:

### SECTION 4: Treatments to Manage Your Depression

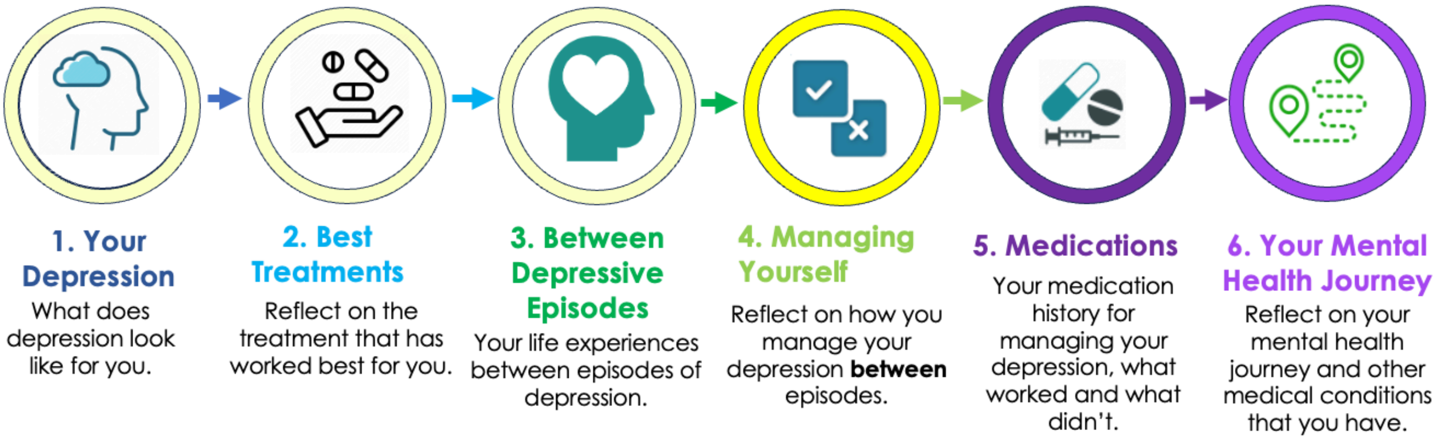

This section of the questionnaire we will ask you what treatments or combination of treatments / strategies you use to keep yourself well to prevent a relapse. The effectiveness of treatments for managing depression is different for everyone. From your experience, what has been the most effective treatment or combination of treatments/ strategies in managing **depressive symptoms between depression episodes that help you to feel at your best**.

*In this section, there are 18 questions, and depending on your responses, this can increase. We anticipate that this section should take no longer than 10-12 minutes to complete.*

**In managing any depressive symptoms between episodes of depression, do you make use of any treatments or combination of treatments?**

Please choose **only one** of the following:

- ◯ Yes
- ◯ No

#### In your experience, what has been the <u>most effective psychological therapy and physical treatment</u> OR <u>combination of these</u> for staying well <u>BETWEEN</u> episdoes of depression - that is, to prevent relapse.

Only answer this question if the following conditions are met:
((treatmentsUsedBtwEpd.NAOK == "Y"))

❗ Check all that apply
Please choose **all** that apply:

- ☐ Cognitive behavioural therapy (CBT)
- ☐ Acceptance and commitment therapy (ACT)
- ☐ Interpersonal therapy (IPT)
- ☐ Dialectical behavioural therapy (DBT)
- ☐ Behavioural therapy
- ☐ Counselling for specific issues/challenges
- ☐ Online or self-management interventions (e.g. mindfulness apps like headspace)
- ☐ Physical exercise (e.g. gym, yoga)
- ☐ Meditation
- ☐ Repetitive Transcranial Magnetic Stimulation (rTMS/TMS)
- ☐ Electroconvulsive therapy (ECT)
- ☐ Light therapy
- ☐ None
- ☐ Other: ____________

#### Please rate the impact of each <u>psychological therapies and physical treatments</u> in managing your depression between episodes.

Only answer this question if the following conditions are met:
((treatmentsUsedBtwEpd.NAOK == "Y")) and ((is_empty(mostEffectiveTherap1_SQ013.NAOK)) and ( ! is_empty(mostEffectiveTherap1)))

Please choose the appropriate response for each item:

| | 1 None | 2 Minor Impact | 3 Some Impact | 4 Moderate Impact | 5 Major Impact | 6 Significant Impact | 7 Extreme Impact |
|---|---|---|---|---|---|---|---|
| **{if(mostEffectiveTherap1_SQ001=="Y", mostEffectiveTherap1_SQ001.question)}** | ○ | ○ | ○ | ○ | ○ | ○ | ○ |
| **{if(mostEffectiveTherap1_SQ006=="Y", mostEffectiveTherap1_SQ006.question)}** | ○ | ○ | ○ | ○ | ○ | ○ | ○ |
| **{if(mostEffectiveTherap1_SQ005=="Y", mostEffectiveTherap1_SQ005.question)}** | ○ | ○ | ○ | ○ | ○ | ○ | ○ |
| **{if(mostEffectiveTherap1_SQ014=="Y", mostEffectiveTherap1_SQ014.question)}** | ○ | ○ | ○ | ○ | ○ | ○ | ○ |
| **{if(mostEffectiveTherap1_SQ003=="Y", mostEffectiveTherap1_SQ003.question)}** | ○ | ○ | ○ | ○ | ○ | ○ | ○ |
| **{if(mostEffectiveTherap1_SQ004=="Y", mostEffectiveTherap1_SQ004.question)}** | ○ | ○ | ○ | ○ | ○ | ○ | ○ |
| **{if(mostEffectiveTherap1_SQ010=="Y", mostEffectiveTherap1_SQ010.question)}** | ○ | ○ | ○ | ○ | ○ | ○ | ○ |
| **{if(mostEffectiveTherap1_SQ011=="Y", mostEffectiveTherap1_SQ011.question)}** | ○ | ○ | ○ | ○ | ○ | ○ | ○ |
| **{if(mostEffectiveTherap1_SQ012=="Y", mostEffectiveTherap1_SQ012.question)}** | ○ | ○ | ○ | ○ | ○ | ○ | ○ |
| **{if(mostEffectiveTherap1_SQ007=="Y", mostEffectiveTherap1_SQ007.question)}** | ○ | ○ | ○ | ○ | ○ | ○ | ○ |
| **{if(mostEffectiveTherap1_SQ008=="Y", mostEffectiveTherap1_SQ008.question)}** | ○ | ○ | ○ | ○ | ○ | ○ | ○ |
| **{if(mostEffectiveTherap1_SQ009=="Y", mostEffectiveTherap1_SQ009.question)}** | ○ | ○ | ○ | ○ | ○ | ○ | ○ |
| **{if(mostEffectiveTherap1_other, mostEffectiveTherap1_other)}** | ○ | ○ | ○ | ○ | ○ | ○ | ○ |

#### This image presents an overview of symptoms people may experience during depression. Think about your set of personal symptoms in each area when answering the questions below.

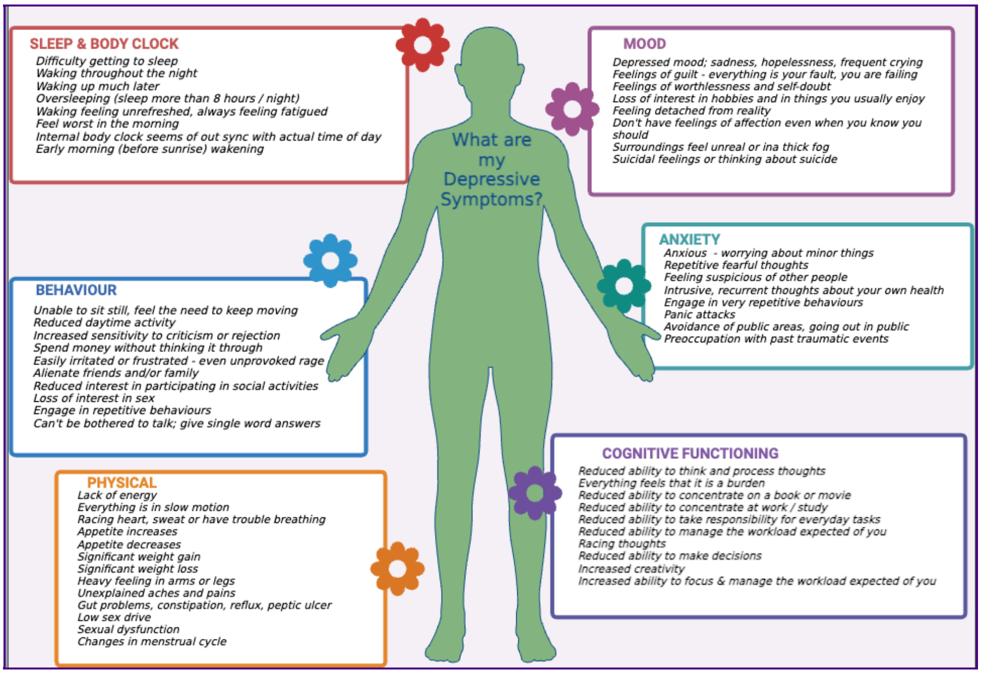

#### Please rate the impact <u>Electroconvulsive Therapy (ECT)</u> had on improving these areas of symptoms that you have reported you experience between episodes of depression.

Only answer this question if the following conditions are met:
((mostEffectiveTherap1_SQ008.NAOK == "Y"))

Please choose the appropriate response for each item:

| | 1 None | 2 Minor Impact | 3 Some Impact | 4 Moderate Impact | 5 Major Impact | 6 Significant Impact | 7 Extreme Impact |
|---|---|---|---|---|---|---|---|
| **Mood** | ○ | ○ | ○ | ○ | ○ | ○ | ○ |
| **Anxiety** | ○ | ○ | ○ | ○ | ○ | ○ | ○ |
| **Memory, ability to focus and think** | ○ | ○ | ○ | ○ | ○ | ○ | ○ |
| **Sleep** | ○ | ○ | ○ | ○ | ○ | ○ | ○ |
| **Behaviour** | ○ | ○ | ○ | ○ | ○ | ○ | ○ |

| | | | | | | | |
|---|---|---|---|---|---|---|---|
| **Physical** | ◯ | ◯ | ◯ | ◯ | ◯ | ◯ | ◯ |

#### Was Electroconvulsive Therapy (ECT) prescribed to you because you experienced any of the following symptoms between your episodes of depression,

#### 1. Psychotic Symptoms
#### 2. Suicidal Thoughts
#### 3. Refusal to eat/drink

Only answer this question if the following conditions are met:
((mostEffectiveTherap1_SQ008.NAOK == "Y"))

Please choose **only one** of the following:

- ◯ Yes
- ◯ No

#### If you benefitted from the psychological therapies and physical treatments for managing depressive symptoms, how quickly did that benefit come on?

Only answer this question if the following conditions are met:
((treatmentsUsedBtwEpd.NAOK == "Y")) and
(is_empty(mostEffectiveTherap1_SQ013.NAOK))

❗ Choose one of the following answers
Please choose **only one** of the following:

- ◯ Can't really specify
- ◯ Different at different times
- ◯ Within two weeks
- ◯ Within four weeks
- ◯ Within four to six weeks
- ◯ Greater than six weeks

**We are now going to ask you about how successful medications have been in managing any symptoms between episodes of depression. Here, we are interested in the most successful medication or combination of medications that have worked for you. In a later section we will ask about experiences with other medications.**

Only answer this question if the following conditions are met:
((treatmentsUsedBtwEpd.NAOK == "Y"))

**In your experience, have you found a single medication OR combination of medications for staying well BETWEEN episodes of depression - that is, to prevent relapse.**

Only answer this question if the following conditions are met:
((treatmentsUsedBtwEpd.NAOK == "Y"))

❶ Choose one of the following answers
Please choose **only one** of the following:

- ◯ Single medication
- ◯ Combination of medications
- ◯ None

#### In your experience, what has been the <u>most effective 'single medication'</u> that you have ever tried for staying well <u>BETWEEN</u> episodes of depression?

***Hover on the ⓘ icon to see some examples for each of the options.***

Only answer this question if the following conditions are met:
((Medication1.NAOK == "AO01"))

❗ Choose one of the following answers
Please choose **only one** of the following:

- ◯ SSRIs (Selective serotonin reuptake inhibitors) ⓘ
- ◯ SNRIs (Serotonin norepinephrine reuptake inhibitors) ⓘ
- ◯ Vortioxetine
- ◯ TCAs (Tricyclic antidepressants) ⓘ
- ◯ TeCAs (Tetracyclic antidepressants) ⓘ
- ◯ MAOIs (Monoamine oxidase inhibitors) ⓘ
- ◯ Melatonin
- ◯ Agomelatine ⓘ
- ◯ Lamotrigine
- ◯ Lithium
- ◯ Antipsychotics ⓘ
- ◯ Stimulants ⓘ
- ◯ Ketamine, Esketamine
- ◯ Psychedelics
- ◯ Medicinal cannabis
- ◯ Brexanolone, Allopregnanolone
- ◯ Other ____________

**In your experience, what has been the <u>most effective "combination of medications"</u> that you have ever tried for staying well <u>BETWEEN</u> episodes of depression?**
***Hover on the ⓘ icon to see some examples for each of the options.***

Only answer this question if the following conditions are met:
((Medication1.NAOK == "AO02"))

❗ Check all that apply
❗ Please select from 2 to 4 answers.
Please choose **all** that apply:

- ☐ SSRIs (Selective serotonin reuptake inhibitors) ⓘ
- ☐ SNRIs (Serotonin norepinephrine reuptake inhibitors) ⓘ
- ☐ Vortioxetine
- ☐ TCAs (Tricyclic antidepressants) ⓘ
- ☐ TeCAs (Tetracyclic antidepressants) ⓘ
- ☐ MAOIs (Monoamine oxidase inhibitors) ⓘ
- ☐ Melatonin
- ☐ Agomelatine ⓘ
- ☐ Lamotrigine
- ☐ Lithium
- ☐ Antipsychotics ⓘ
- ☐ Stimulants ⓘ
- ☐ Ketamine, Esketamine
- ☐ Psychedelics
- ☐ Medicinal cannabis
- ☐ Brexanolone, Allopregnanolone
- ☐ Other: ____________

### SECTION 4 (cont.): Treatments to Manage Your Depression

#### Please rate the impact of the medication for staying well **BETWEEN** episodes of depression

Only answer this question if the following conditions are met:
((treatmentsUsedBtwEpd.NAOK == "Y")) and ((Medication1.NAOK == "AO01"))

Please choose the appropriate response for each item:

| | 1 None | 2 Minor Impact | 3 Some Impact | 4 Moderate Impact | 5 Major Impact | 6 Significant Impact | 7 Extreme Impact |
|---|---|---|---|---|---|---|---|
| {if(singleMedication1=="AO01", CombinationMeds1_SQ001.question)} | ○ | ○ | ○ | ○ | ○ | ○ | ○ |
| {if(singleMedication1=="AO02", CombinationMeds1_SQ002.question)} | ○ | ○ | ○ | ○ | ○ | ○ | ○ |
| {if(singleMedication1=="AO16", CombinationMeds1_SQ016.question)} | ○ | ○ | ○ | ○ | ○ | ○ | ○ |
| {if(singleMedication1=="AO03", CombinationMeds1_SQ003.question)} | ○ | ○ | ○ | ○ | ○ | ○ | ○ |
| {if(singleMedication1=="AO04", CombinationMeds1_SQ004.question)} | ○ | ○ | ○ | ○ | ○ | ○ | ○ |
| {if(singleMedication1=="AO05", CombinationMeds1_SQ005.question)} | ○ | ○ | ○ | ○ | ○ | ○ | ○ |
| {if(singleMedication1=="AO06", CombinationMeds1_SQ006.question)} | ○ | ○ | ○ | ○ | ○ | ○ | ○ |
| {if(singleMedication1=="AO07", CombinationMeds1_SQ007.question)} | ○ | ○ | ○ | ○ | ○ | ○ | ○ |
| {if(singleMedication1=="AO08", CombinationMeds1_SQ008.question)} | ○ | ○ | ○ | ○ | ○ | ○ | ○ |
| {if(singleMedication1=="AO09", CombinationMeds1_SQ009.question)} | ○ | ○ | ○ | ○ | ○ | ○ | ○ |
| {if(singleMedication1=="AO10", CombinationMeds1_SQ010.question)} | ○ | ○ | ○ | ○ | ○ | ○ | ○ |
| {if(singleMedication1=="AO11", CombinationMeds1_SQ011.question)} | ○ | ○ | ○ | ○ | ○ | ○ | ○ |
| {if(singleMedication1=="AO12", CombinationMeds1_SQ012.question)} | ○ | ○ | ○ | ○ | ○ | ○ | ○ |
| {if(singleMedication1=="AO13", CombinationMeds1_SQ013.question)} | ○ | ○ | ○ | ○ | ○ | ○ | ○ |
| {if(singleMedication1=="AO14", CombinationMeds1_SQ014.question)} | ○ | ○ | ○ | ○ | ○ | ○ | ○ |
| {if(singleMedication1=="-oth-", singleMedication1_other)} | ○ | ○ | ○ | ○ | ○ | ○ | ○ |
| {if(singleMedication1=="AO17", CombinationMeds1_SQ017.question)} | ○ | ○ | ○ | ○ | ○ | ○ | ○ |

#### This image presents an overview of symptoms people may experience during depression. Think about your set of personal symptoms in each area when answering the questions below.

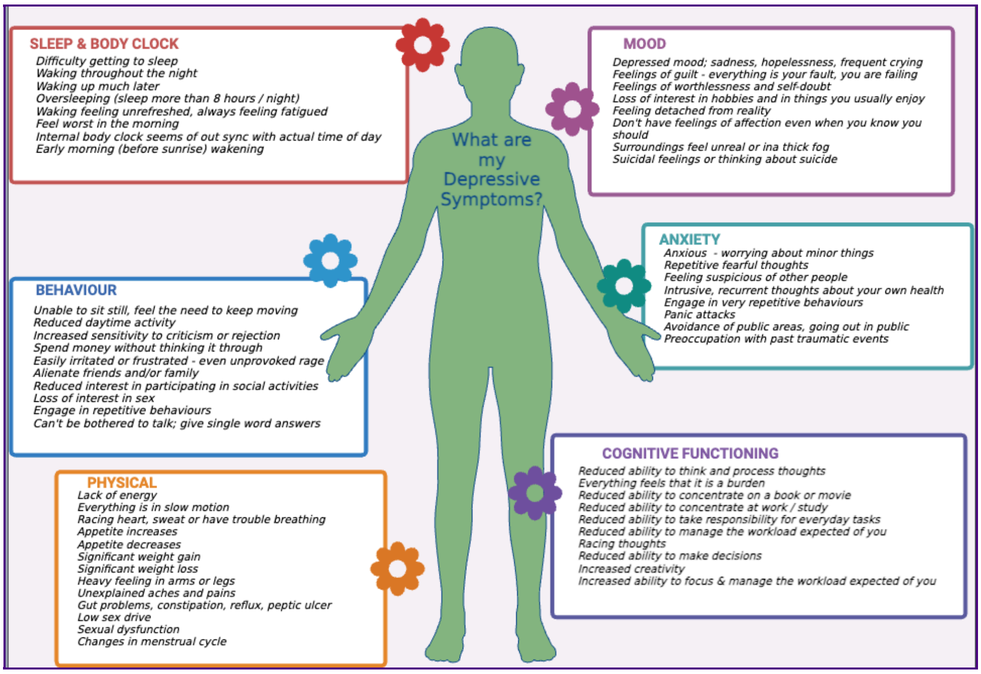

#### Please rate the impact of the medication that helped in improving the following <u>areas of symptoms</u> that you have reported you experience when staying well <u>BETWEEN</u> episodes of depression.

Only answer this question if the following conditions are met:
((treatmentsUsedBtwEpd.NAOK == "Y")) and ( ! is_empty(singleMedication1))

Please choose the appropriate response for each item:

| | 1 None | 2 Minor Impact | 3 Some Impact | 4 Moderate Impact | 5 Major Impact | 6 Significant Impact | 7 Extreme Impact |
|---|---|---|---|---|---|---|---|
| **Mood** | ○ | ○ | ○ | ○ | ○ | ○ | ○ |
| **Anxiety** | ○ | ○ | ○ | ○ | ○ | ○ | ○ |
| **Memory, ability to focus and think** | ○ | ○ | ○ | ○ | ○ | ○ | ○ |

| | | | | | | | |
|---|---|---|---|---|---|---|---|
| **Sleep** | ◯ | ◯ | ◯ | ◯ | ◯ | ◯ | ◯ |
| **Behaviour** | ◯ | ◯ | ◯ | ◯ | ◯ | ◯ | ◯ |
| **Physical** | ◯ | ◯ | ◯ | ◯ | ◯ | ◯ | ◯ |

#### Please rate the impact of the medications for staying well BETWEEN episodes of depression

Only answer this question if the following conditions are met:
((treatmentsUsedBtwEpd.NAOK == "Y")) and ((Medication1.NAOK == "AO02"))

Please choose the appropriate response for each item:

| | 1 None | 2 Minor Impact | 3 Some Impact | 4 Moderate Impact | 5 Major Impact | 6 Significant Impact | 7 Extreme Impact |
|---|---|---|---|---|---|---|---|
| {if(CombinationMeds1_SQ001=="Y", CombinationMeds1_SQ001.question)} | ○ | ○ | ○ | ○ | ○ | ○ | ○ |
| {if(CombinationMeds1_SQ002=="Y", CombinationMeds1_SQ002.question)} | ○ | ○ | ○ | ○ | ○ | ○ | ○ |
| {if(CombinationMeds1_SQ016=="Y", CombinationMeds1_SQ016.question)} | ○ | ○ | ○ | ○ | ○ | ○ | ○ |
| {if(CombinationMeds1_SQ003=="Y", CombinationMeds1_SQ003.question)} | ○ | ○ | ○ | ○ | ○ | ○ | ○ |
| {if(CombinationMeds1_SQ004=="Y", CombinationMeds1_SQ004.question)} | ○ | ○ | ○ | ○ | ○ | ○ | ○ |
| {if(CombinationMeds1_SQ005=="Y", CombinationMeds1_SQ005.question)} | ○ | ○ | ○ | ○ | ○ | ○ | ○ |
| {if(CombinationMeds1_SQ006=="Y", CombinationMeds1_SQ006.question)} | ○ | ○ | ○ | ○ | ○ | ○ | ○ |
| {if(CombinationMeds1_SQ007=="Y", CombinationMeds1_SQ007.question)} | ○ | ○ | ○ | ○ | ○ | ○ | ○ |
| {if(CombinationMeds1_SQ008=="Y", CombinationMeds1_SQ008.question)} | ○ | ○ | ○ | ○ | ○ | ○ | ○ |
| {if(CombinationMeds1_SQ009=="Y", CombinationMeds1_SQ009.question)} | ○ | ○ | ○ | ○ | ○ | ○ | ○ |
| {if(CombinationMeds1_SQ010=="Y", CombinationMeds1_SQ010.question)} | ○ | ○ | ○ | ○ | ○ | ○ | ○ |
| {if(CombinationMeds1_SQ011=="Y", CombinationMeds1_SQ011.question)} | ○ | ○ | ○ | ○ | ○ | ○ | ○ |
| {if(CombinationMeds1_SQ012=="Y", CombinationMeds1_SQ012.question)} | ○ | ○ | ○ | ○ | ○ | ○ | ○ |
| {if(CombinationMeds1_SQ013=="Y", CombinationMeds1_SQ013.question)} | ○ | ○ | ○ | ○ | ○ | ○ | ○ |
| {if(CombinationMeds1_SQ014=="Y", CombinationMeds1_SQ014.question)} | ○ | ○ | ○ | ○ | ○ | ○ | ○ |
| {if(CombinationMeds1_other, CombinationMeds1_other)} | ○ | ○ | ○ | ○ | ○ | ○ | ○ |
| {if(CombinationMeds1_SQ017=="Y", CombinationMeds1_SQ017.question)} | ○ | ○ | ○ | ○ | ○ | ○ | ○ |

#### This image presents an overview of symptoms people may experience during depression. Think about your set of personal symptoms in each area when answering the questions below.

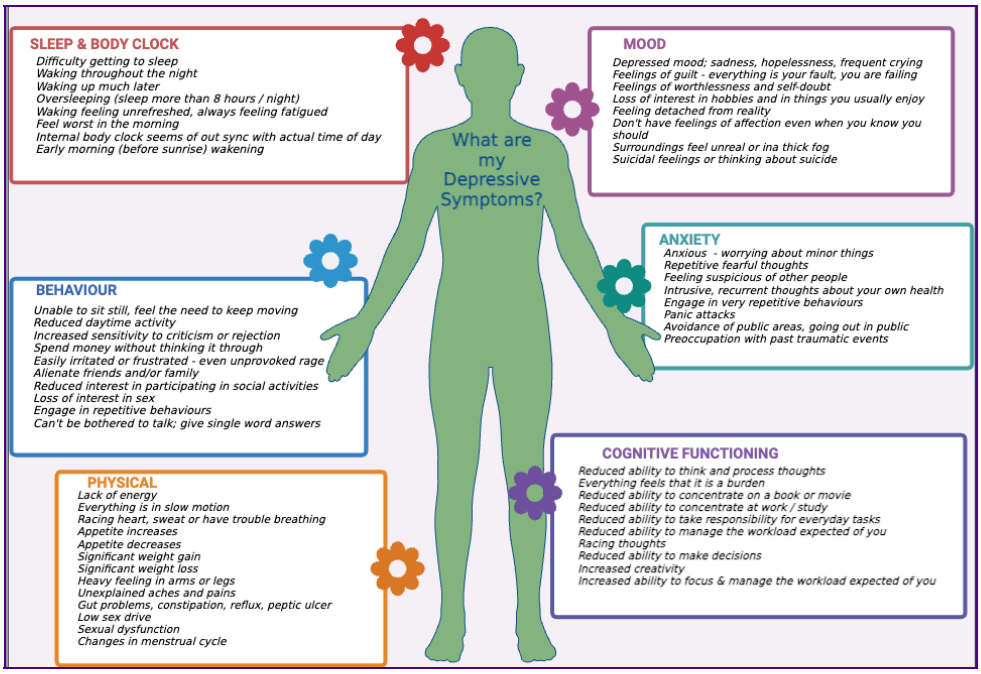

#### Please rate the impact of the following medications that helped in improving the following areas of symptoms that you have reported you experience when staying well BETWEEN episodes of depression.

Only answer this question if the following conditions are met:
((treatmentsUsedBtwEpd.NAOK == "Y")) and ( ! is_empty(CombinationMeds1))

Please choose the appropriate response for each item:

| | 1 None | 2 Minor Impact | 3 Some Impact | 4 Moderate Impact | 5 Major Impact | 6 Significant Impact | 7 Extreme Impact |
|---|---|---|---|---|---|---|---|
| {if(CombinationMeds1_SQ001=="Y", CombinationMeds1_SQ001.question)} Mood | ○ | ○ | ○ | ○ | ○ | ○ | ○ |
| {if(CombinationMeds1_SQ001=="Y", CombinationMeds1_SQ001.question)} Anxiety | ○ | ○ | ○ | ○ | ○ | ○ | ○ |
| {if(CombinationMeds1_SQ001=="Y", CombinationMeds1_SQ001.question)} | ○ | ○ | ○ | ○ | ○ | ○ | ○ |

Memory, ability to focus and think

| | | | | | | |
|---|---|---|---|---|---|---|
| {if(CombinationMeds1_SQ001=="Y", CombinationMeds1_SQ001.question)} Sleep | ◯ | ◯ | ◯ | ◯ | ◯ | ◯ |
| {if(CombinationMeds1_SQ001=="Y", CombinationMeds1_SQ001.question)} Behaviour | ◯ | ◯ | ◯ | ◯ | ◯ | ◯ |
| {if(CombinationMeds1_SQ001=="Y", CombinationMeds1_SQ001.question)} Physical | ◯ | ◯ | ◯ | ◯ | ◯ | ◯ |
| {if(CombinationMeds1_SQ002=="Y", CombinationMeds1_SQ002.question)} Mood | ◯ | ◯ | ◯ | ◯ | ◯ | ◯ |
| {if(CombinationMeds1_SQ002=="Y", CombinationMeds1_SQ002.question)} Anxiety | ◯ | ◯ | ◯ | ◯ | ◯ | ◯ |
| {if(CombinationMeds1_SQ002=="Y", CombinationMeds1_SQ002.question)} Memory, ability to focus and think | ◯ | ◯ | ◯ | ◯ | ◯ | ◯ |
| {if(CombinationMeds1_SQ002=="Y", CombinationMeds1_SQ002.question)} Sleep | ◯ | ◯ | ◯ | ◯ | ◯ | ◯ |
| {if(CombinationMeds1_SQ002=="Y", CombinationMeds1_SQ002.question)} Behaviour | ◯ | ◯ | ◯ | ◯ | ◯ | ◯ |
| {if(CombinationMeds1_SQ002=="Y", CombinationMeds1_SQ002.question)} Physical | ◯ | ◯ | ◯ | ◯ | ◯ | ◯ |
| {if(CombinationMeds1_SQ016=="Y", CombinationMeds1_SQ016.question)} Mood | ◯ | ◯ | ◯ | ◯ | ◯ | ◯ |
| {if(CombinationMeds1_SQ016=="Y", CombinationMeds1_SQ016.question)} Anxiety | ◯ | ◯ | ◯ | ◯ | ◯ | ◯ |
| {if(CombinationMeds1_SQ016=="Y", CombinationMeds1_SQ016.question)} Memory, ability to focus and think | ◯ | ◯ | ◯ | ◯ | ◯ | ◯ |
| {if(CombinationMeds1_SQ016=="Y", CombinationMeds1_SQ016.question)} Sleep | ◯ | ◯ | ◯ | ◯ | ◯ | ◯ |
| {if(CombinationMeds1_SQ016=="Y", CombinationMeds1_SQ016.question)} Behaviour | ◯ | ◯ | ◯ | ◯ | ◯ | ◯ |
| {if(CombinationMeds1_SQ016=="Y", CombinationMeds1_SQ016.question)} Physical | ◯ | ◯ | ◯ | ◯ | ◯ | ◯ |

{if(CombinationMeds1_SQ003=="Y",

| | | | | | | | |
|---|---|---|---|---|---|---|---|
| CombinationMeds1_SQ003.question)} Mood | ○ | ○ | ○ | ○ | ○ | ○ | ○ |
| {if(CombinationMeds1_SQ003=="Y", CombinationMeds1_SQ003.question)} Anxiety | ○ | ○ | ○ | ○ | ○ | ○ | ○ |
| {if(CombinationMeds1_SQ003=="Y", CombinationMeds1_SQ003.question)} Memory, ability to focus and think | ○ | ○ | ○ | ○ | ○ | ○ | ○ |
| {if(CombinationMeds1_SQ003=="Y", CombinationMeds1_SQ003.question)} Sleep | ○ | ○ | ○ | ○ | ○ | ○ | ○ |
| {if(CombinationMeds1_SQ003=="Y", CombinationMeds1_SQ003.question)} Behaviour | ○ | ○ | ○ | ○ | ○ | ○ | ○ |
| {if(CombinationMeds1_SQ003=="Y", CombinationMeds1_SQ003.question)} Physical | ○ | ○ | ○ | ○ | ○ | ○ | ○ |
| {if(CombinationMeds1_SQ004=="Y", CombinationMeds1_SQ004.question)} Mood | ○ | ○ | ○ | ○ | ○ | ○ | ○ |
| {if(CombinationMeds1_SQ004=="Y", CombinationMeds1_SQ004.question)} Anxiety | ○ | ○ | ○ | ○ | ○ | ○ | ○ |
| {if(CombinationMeds1_SQ004=="Y", CombinationMeds1_SQ004.question)} Memory, ability to focus and think | ○ | ○ | ○ | ○ | ○ | ○ | ○ |
| {if(CombinationMeds1_SQ004=="Y", CombinationMeds1_SQ004.question)} Sleep | ○ | ○ | ○ | ○ | ○ | ○ | ○ |
| {if(CombinationMeds1_SQ004=="Y", CombinationMeds1_SQ004.question)} Behaviour | ○ | ○ | ○ | ○ | ○ | ○ | ○ |
| {if(CombinationMeds1_SQ004=="Y", CombinationMeds1_SQ004.question)} Physical | ○ | ○ | ○ | ○ | ○ | ○ | ○ |
| {if(CombinationMeds1_SQ005=="Y", CombinationMeds1_SQ005.question)} Mood | ○ | ○ | ○ | ○ | ○ | ○ | ○ |
| {if(CombinationMeds1_SQ005=="Y", CombinationMeds1_SQ005.question)} Anxiety | ○ | ○ | ○ | ○ | ○ | ○ | ○ |
| {if(CombinationMeds1_SQ005=="Y", CombinationMeds1_SQ005.question)} Memory, ability to focus and think | ○ | ○ | ○ | ○ | ○ | ○ | ○ |
| {if(CombinationMeds1_SQ005=="Y", CombinationMeds1_SQ005.question)} Sleep | ○ | ○ | ○ | ○ | ○ | ○ | ○ |

{if(CombinationMeds1_SQ005=="Y", CombinationMeds1_SQ005.question)} Behaviour

{if(CombinationMeds1_SQ005=="Y", CombinationMeds1_SQ005.question)} Physical

{if(CombinationMeds1_SQ006=="Y", CombinationMeds1_SQ006.question)} Mood

{if(CombinationMeds1_SQ006=="Y", CombinationMeds1_SQ006.question)} Anxiety

{if(CombinationMeds1_SQ006=="Y", CombinationMeds1_SQ006.question)} Memory, ability to focus and think

{if(CombinationMeds1_SQ006=="Y", CombinationMeds1_SQ006.question)} Sleep

{if(CombinationMeds1_SQ006=="Y", CombinationMeds1_SQ006.question)} Behaviour

{if(CombinationMeds1_SQ006=="Y", CombinationMeds1_SQ006.question)} Physical

{if(CombinationMeds1_SQ007=="Y", CombinationMeds1_SQ007.question)} Mood

{if(CombinationMeds1_SQ007=="Y", CombinationMeds1_SQ007.question)} Anxiety

{if(CombinationMeds1_SQ007=="Y", CombinationMeds1_SQ007.question)} Memory, ability to focus and think

{if(CombinationMeds1_SQ007=="Y", CombinationMeds1_SQ007.question)} Sleep

{if(CombinationMeds1_SQ007=="Y", CombinationMeds1_SQ007.question)} Behaviour

{if(CombinationMeds1_SQ007=="Y", CombinationMeds1_SQ007.question)} Physical

{if(CombinationMeds1_SQ008=="Y", CombinationMeds1_SQ008.question)} Mood

{if(CombinationMeds1_SQ008=="Y", CombinationMeds1_SQ008.question)} Anxiety

| | | | | | | | |
|---|---|---|---|---|---|---|---|
| {if(CombinationMeds1_SQ008=="Y", CombinationMeds1_SQ008.question)} Memory, ability to focus and think | ◯ | ◯ | ◯ | ◯ | ◯ | ◯ | ◯ |
| {if(CombinationMeds1_SQ008=="Y", CombinationMeds1_SQ008.question)} Sleep | ◯ | ◯ | ◯ | ◯ | ◯ | ◯ | ◯ |
| {if(CombinationMeds1_SQ008=="Y", CombinationMeds1_SQ008.question)} Behaviour | ◯ | ◯ | ◯ | ◯ | ◯ | ◯ | ◯ |
| {if(CombinationMeds1_SQ008=="Y", CombinationMeds1_SQ008.question)} Physical | ◯ | ◯ | ◯ | ◯ | ◯ | ◯ | ◯ |
| {if(CombinationMeds1_SQ009=="Y", CombinationMeds1_SQ009.question)} Mood | ◯ | ◯ | ◯ | ◯ | ◯ | ◯ | ◯ |
| {if(CombinationMeds1_SQ009=="Y", CombinationMeds1_SQ009.question)} Anxiety | ◯ | ◯ | ◯ | ◯ | ◯ | ◯ | ◯ |
| {if(CombinationMeds1_SQ009=="Y", CombinationMeds1_SQ009.question)} Memory, ability to focus and think | ◯ | ◯ | ◯ | ◯ | ◯ | ◯ | ◯ |
| {if(CombinationMeds1_SQ009=="Y", CombinationMeds1_SQ009.question)} Sleep | ◯ | ◯ | ◯ | ◯ | ◯ | ◯ | ◯ |
| {if(CombinationMeds1_SQ009=="Y", CombinationMeds1_SQ009.question)} Behaviour | ◯ | ◯ | ◯ | ◯ | ◯ | ◯ | ◯ |
| {if(CombinationMeds1_SQ009=="Y", CombinationMeds1_SQ009.question)} Physical | ◯ | ◯ | ◯ | ◯ | ◯ | ◯ | ◯ |
| {if(CombinationMeds1_SQ010=="Y", CombinationMeds1_SQ010.question)} Mood | ◯ | ◯ | ◯ | ◯ | ◯ | ◯ | ◯ |
| {if(CombinationMeds1_SQ010=="Y", CombinationMeds1_SQ010.question)} Anxiety | ◯ | ◯ | ◯ | ◯ | ◯ | ◯ | ◯ |
| {if(CombinationMeds1_SQ010=="Y", CombinationMeds1_SQ010.question)} Memory, ability to focus and think | ◯ | ◯ | ◯ | ◯ | ◯ | ◯ | ◯ |
| {if(CombinationMeds1_SQ010=="Y", CombinationMeds1_SQ010.question)} Sleep | ◯ | ◯ | ◯ | ◯ | ◯ | ◯ | ◯ |
| {if(CombinationMeds1_SQ010=="Y", CombinationMeds1_SQ010.question)} Behaviour | ◯ | ◯ | ◯ | ◯ | ◯ | ◯ | ◯ |
| {if(CombinationMeds1_SQ010=="Y", CombinationMeds1_SQ010.question)} | | | | | | | |

| | | | | | | | |
|---|---|---|---|---|---|---|---|
| Physical | ◯ | ◯ | ◯ | ◯ | ◯ | ◯ | ◯ |
| {if(CombinationMeds1_SQ011=="Y", CombinationMeds1_SQ011.question)} Mood | ◯ | ◯ | ◯ | ◯ | ◯ | ◯ | ◯ |
| {if(CombinationMeds1_SQ011=="Y", CombinationMeds1_SQ011.question)} Anxiety | ◯ | ◯ | ◯ | ◯ | ◯ | ◯ | ◯ |
| {if(CombinationMeds1_SQ011=="Y", CombinationMeds1_SQ011.question)} Memory, ability to focus and think | ◯ | ◯ | ◯ | ◯ | ◯ | ◯ | ◯ |
| {if(CombinationMeds1_SQ011=="Y", CombinationMeds1_SQ011.question)} Sleep | ◯ | ◯ | ◯ | ◯ | ◯ | ◯ | ◯ |
| {if(CombinationMeds1_SQ011=="Y", CombinationMeds1_SQ011.question)} Behaviour | ◯ | ◯ | ◯ | ◯ | ◯ | ◯ | ◯ |
| {if(CombinationMeds1_SQ011=="Y", CombinationMeds1_SQ011.question)} Physical | ◯ | ◯ | ◯ | ◯ | ◯ | ◯ | ◯ |
| {if(CombinationMeds1_SQ012=="Y", CombinationMeds1_SQ012.question)} Mood | ◯ | ◯ | ◯ | ◯ | ◯ | ◯ | ◯ |
| {if(CombinationMeds1_SQ012=="Y", CombinationMeds1_SQ012.question)} Anxiety | ◯ | ◯ | ◯ | ◯ | ◯ | ◯ | ◯ |
| {if(CombinationMeds1_SQ012=="Y", CombinationMeds1_SQ012.question)} Memory, ability to focus and think | ◯ | ◯ | ◯ | ◯ | ◯ | ◯ | ◯ |
| {if(CombinationMeds1_SQ012=="Y", CombinationMeds1_SQ012.question)} Sleep | ◯ | ◯ | ◯ | ◯ | ◯ | ◯ | ◯ |
| {if(CombinationMeds1_SQ012=="Y", CombinationMeds1_SQ012.question)} Behaviour | ◯ | ◯ | ◯ | ◯ | ◯ | ◯ | ◯ |
| {if(CombinationMeds1_SQ012=="Y", CombinationMeds1_SQ012.question)} Physical | ◯ | ◯ | ◯ | ◯ | ◯ | ◯ | ◯ |
| {if(CombinationMeds1_SQ013=="Y", CombinationMeds1_SQ013.question)} Mood | ◯ | ◯ | ◯ | ◯ | ◯ | ◯ | ◯ |
| {if(CombinationMeds1_SQ013=="Y", CombinationMeds1_SQ013.question)} Anxiety | ◯ | ◯ | ◯ | ◯ | ◯ | ◯ | ◯ |
| {if(CombinationMeds1_SQ013=="Y", CombinationMeds1_SQ013.question)} Memory, ability to focus and think | ◯ | ◯ | ◯ | ◯ | ◯ | ◯ | ◯ |

{if(CombinationMeds1_SQ013=="Y", CombinationMeds1_SQ013.question)}
Sleep

{if(CombinationMeds1_SQ013=="Y", CombinationMeds1_SQ013.question)}
Behaviour

{if(CombinationMeds1_SQ013=="Y", CombinationMeds1_SQ013.question)}
Physical

{if(CombinationMeds1_SQ014=="Y", CombinationMeds1_SQ014.question)}
Mood

{if(CombinationMeds1_SQ014=="Y", CombinationMeds1_SQ014.question)}
Anxiety

{if(CombinationMeds1_SQ014=="Y", CombinationMeds1_SQ014.question)}
Memory, ability to focus and think

{if(CombinationMeds1_SQ014=="Y", CombinationMeds1_SQ014.question)}
Sleep

{if(CombinationMeds1_SQ014=="Y", CombinationMeds1_SQ014.question)}
Behaviour

{if(CombinationMeds1_SQ014=="Y", CombinationMeds1_SQ014.question)}
Physical

{if(CombinationMeds1_other, CombinationMeds1_other)}
Mood

{if(CombinationMeds1_other, CombinationMeds1_other)}
Anxiety

{if(CombinationMeds1_other, CombinationMeds1_other)}
Memory, ability to focus and think

{if(CombinationMeds1_other, CombinationMeds1_other)}
Sleep

{if(CombinationMeds1_other, CombinationMeds1_other)}
Behaviour

{if(CombinationMeds1_other, CombinationMeds1_other)}
Physical

{if(CombinationMeds1_SQ017=="Y", CombinationMeds1_SQ017.question)}
Mood

| | | | | | | | |
|---|---|---|---|---|---|---|---|
| {if(CombinationMeds1_SQ017=="Y", CombinationMeds1_SQ017.question)} **Anxiety** | ○ | ○ | ○ | ○ | ○ | ○ | ○ |
| {if(CombinationMeds1_SQ017=="Y", CombinationMeds1_SQ017.question)} **Memory, ability to focus and think** | ○ | ○ | ○ | ○ | ○ | ○ | ○ |
| {if(CombinationMeds1_SQ017=="Y", CombinationMeds1_SQ017.question)} **Sleep** | ○ | ○ | ○ | ○ | ○ | ○ | ○ |
| {if(CombinationMeds1_SQ017=="Y", CombinationMeds1_SQ017.question)} **Behaviour** | ○ | ○ | ○ | ○ | ○ | ○ | ○ |
| {if(CombinationMeds1_SQ017=="Y", CombinationMeds1_SQ017.question)} **Physical** | ○ | ○ | ○ | ○ | ○ | ○ | ○ |

#### If you benefitted from the most effective medication or medication combination for staying well BETWEEN episodes of depression, how quickly did that benefit come on?

Only answer this question if the following conditions are met:
((treatmentsUsedBtwEpd.NAOK == "Y")) and (is_empty(Medication1.NAOK == "AO03"))

❗ Choose one of the following answers
Please choose **only one** of the following:

- ○ Can't really specify
- ○ Different at different times
- ○ Within two weeks
- ○ Within four weeks
- ○ Within four to six weeks
- ○ Greater than six weeks

#### Do you experience any side effects from the most effective medications for staying well BETWEEN the episodes of depression?

Only answer this question if the following conditions are met:
((treatmentsUsedBtwEpd.NAOK == "Y")) and (is_empty(Medication1.NAOK == "AO03"))

Please choose **only one** of the following:

- ◯ Yes
- ◯ No

#### Can you please describe the side-effects you experienced?

Only answer this question if the following conditions are met:
((medSideEffects1.NAOK == "Y"))

❗ Comment only when you choose an answer.
Please choose all that apply and provide a comment:

- ☐ Weight gain
- ☐ Nausea
- ☐ Dizziness
- ☐ Headaches
- ☐ Sleepy, zombie-like
- ☐ Loss of interest in sex
- ☐ Diarrhoea
- ☐ Other gut problems
- ☐ Other side effects

Please use the free text box to give more details about any side effects

#### When staying well <u>BETWEEN</u> the episodes of depression, have you used any of the following substances to try and manage your anxiety, depression or related mental health problems?

Only answer this question if the following conditions are met:
((treatmentsUsedBtwEpd.NAOK == "Y"))

Please choose the appropriate response for each item:

| | Yes | No |
|---|---|---|
| **Alcohol** | ○ | ○ |
| **Tobacco** | ○ | ○ |
| **Cannabis** | ○ | ○ |
| **Stimulants** | ○ | ○ |
| **Other Substances** | ○ | ○ |

❗ Check all that apply
Please choose **all** that apply:

- ☐ Psychological therapy
- ☐ Physical Treatment
- ☐ Medications

#### If you are able to rank the order of the successful therapy, or physical treatment, or medication for staying well BETWEEN your episodes of depression, then add the ranking.

Only answer this question if the following conditions are met:
((treatmentsUsedBtwEpd.NAOK == "Y")) and
((is_empty(mostEffectiveTherap1_SQ013.NAOK)) or (Medication1.NAOK == "AO01" or Medication1.NAOK == "AO02"))

❗ Please select at most 3 answers
Please number each box in order of preference from 1 to 3

- [ ] Psychological therapy
- [ ] Physical treatment
- [ ] Medications

Double-click or drag-and-drop items in the left list to move them to the right - your highest ranking item should be on the top right, moving through to your lowest ranking item.

#### Please describe what other substances you have tried when staying well BETWEEN your episodes of depression. Do not include substances that have been prescribed by a medical professional.

Only answer this question if the following conditions are met:
((substancesUsage1_SQ002.NAOK == "AO01"))

Please write your answer here:

**If there any other treatments you used for staying well BETWEEN episodes of depression that were not included in the lists above please describe them below.**

Only answer this question if the following conditions are met:
((treatmentsUsedBtwEpd.NAOK == "Y"))

Please write your answer here:

### SECTION 5: All Medications For Depression

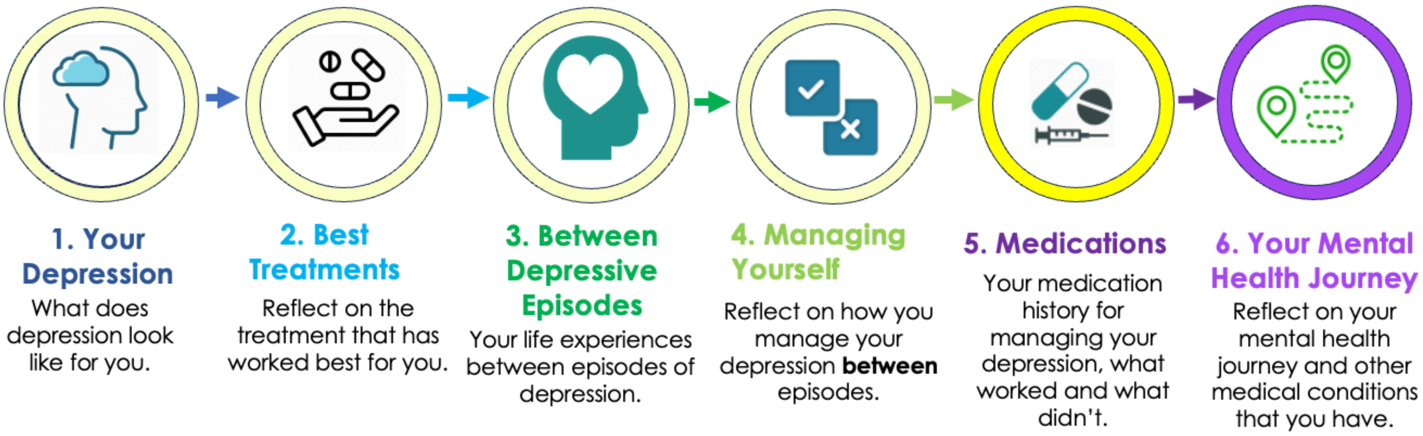

In this section of the questionnaire, we will be asking questions about **any** medications you have taken to manage your depression. In previous sections we focussed on the treatments that have worked best for you. In this section we want to focus on medications that you have **ever** tried and used.

*In this section, there are 25 questions, and depending on your responses, this can increase. We anticipate that this section should take no longer than 10 minutes to complete.*

#### Are you currently taking any medication to treat your depression?

Please choose **only one** of the following:

- ◯ Yes
- ◯ No

#### In previous sections you told us that the medication that worked best for you is '{singleMedication.shown}'.
#### Is that the medication you are currently taking?

Only answer this question if the following conditions are met:
((takingMedsCurrently.NAOK == "Y") and (Medication.NAOK == "AO01"))

Please choose **only one** of the following:

- ◯ Yes
- ◯ No

**In previous sections you told us that the medications that worked best for you are '{if(CombinationMeds_SQ001=="Y", join(CombinationMeds_SQ001.question, ","))} {if(CombinationMeds_SQ002=="Y", join(CombinationMeds_SQ002.question, ","))} {if(CombinationMeds_SQ003=="Y", join(CombinationMeds_SQ003.question, ","))} {if(CombinationMeds_SQ004=="Y", join(CombinationMeds_SQ004.question, ","))} {if(CombinationMeds_SQ005=="Y", join(CombinationMeds_SQ005.question, ","))} {if(CombinationMeds_SQ006=="Y", join(CombinationMeds_SQ006.question, ","))} {if(CombinationMeds_SQ007=="Y", join(CombinationMeds_SQ007.question, ","))} {if(CombinationMeds_SQ008=="Y", join(CombinationMeds_SQ008.question, ","))} {if(CombinationMeds_SQ009=="Y", join(CombinationMeds_SQ009.question, ","))} {if(CombinationMeds_SQ010=="Y", join(CombinationMeds_SQ010.question, ","))} {if(CombinationMeds_SQ011=="Y", join(CombinationMeds_SQ011.question, ","))} {if(CombinationMeds_SQ012=="Y", join(CombinationMeds_SQ012.question, ","))} {if(CombinationMeds_SQ013=="Y", join(CombinationMeds_SQ013.question, ","))} {if(CombinationMeds_SQ014=="Y", join(CombinationMeds_SQ014.question, ","))} {if(CombinationMeds_other, join(CombinationMeds_other, "."))}'**
**Are these the medications you are currently taking?**

Only answer this question if the following conditions are met:
((takingMedsCurrently.NAOK == "Y") and (Medication.NAOK == "AO02"))

Please choose **only one** of the following:

- ◯ Yes
- ◯ No

**In previous sections you told us that the medication that worked best for you is '{singleMedication.shown}'.**
**Have you taken this medication in the last year?**

Only answer this question if the following conditions are met:
((takingMedsCurrently.NAOK == "N") and (Medication.NAOK == "AO01"))

Please choose **only one** of the following:

- Yes
- No

**In previous sections you told us that the medications that worked best for you are '{if(CombinationMeds_SQ001=="Y", join(CombinationMeds_SQ001.question, ","))}**
**{if(CombinationMeds_SQ002=="Y", join(CombinationMeds_SQ002.question, ","))}**
**{if(CombinationMeds_SQ003=="Y", join(CombinationMeds_SQ003.question, ","))}**
**{if(CombinationMeds_SQ004=="Y", join(CombinationMeds_SQ004.question, ","))}**
**{if(CombinationMeds_SQ005=="Y", join(CombinationMeds_SQ005.question, ","))}**
**{if(CombinationMeds_SQ006=="Y", join(CombinationMeds_SQ006.question, ","))}**
**{if(CombinationMeds_SQ007=="Y", join(CombinationMeds_SQ007.question, ","))}**
**{if(CombinationMeds_SQ008=="Y", join(CombinationMeds_SQ008.question, ","))}**
**{if(CombinationMeds_SQ009=="Y", join(CombinationMeds_SQ009.question, ","))}**
**{if(CombinationMeds_SQ010=="Y", join(CombinationMeds_SQ010.question, ","))}**
**{if(CombinationMeds_SQ011=="Y", join(CombinationMeds_SQ011.question, ","))}**
**{if(CombinationMeds_SQ012=="Y", join(CombinationMeds_SQ012.question, ","))}**
**{if(CombinationMeds_SQ013=="Y",**

**join(CombinationMeds_SQ013.question, ","))}**
**{if(CombinationMeds_SQ014=="Y",**
**join(CombinationMeds_SQ014.question, ","))}**
**{if(CombinationMeds_other, join(CombinationMeds_other,**
**"."))}'**

#### Have you taken these medications in the last year?

Only answer this question if the following conditions are met:
((takingMedsCurrently.NAOK == "N") and (Medication.NAOK == "AO02"))

Please choose **only one** of the following:

- ◯ Yes
- ◯ No

#### Have you ever been prescribed medications for your depression?

Only answer this question if the following conditions are met:
((Medication.NAOK == "AO03") or (positiveChange.NAOK == "N"))

Please choose **only one** of the following:

- ◯ Yes
- ◯ No

#### What medications from the list below have you <u>ever</u> been prescribed?

❗ Check all that apply
Please choose **all** that apply:

- ☐ SSRIs (Selective serotonin reuptake inhibitors)
- ☐ SNRIs (Serotonin norepinephrine reuptake inhibitors)
- ☐ TCAs (Tricyclic antidepressants)
- ☐ TeCAs (Tetracyclic antidepressants)
- ☐ MAOIs (Monoamine oxidase inhibitors)
- ☐ Melatonin
- ☐ Agomelatine
- ☐ Lamotrigine
- ☐ Lithium
- ☐ Antipsychotics
- ☐ Stimulants
- ☐ Ketamine, Esketamine
- ☐ Psychedelics
- ☐ Medicinal cannabis
- ☐ Other: ____________________

**What prescribed medications have you taken in the last year to manage your depression?**

❗ Check all that apply
Please choose **all** that apply:

- ☐ SSRIs (Selective serotonin reuptake inhibitors)
- ☐ SNRIs (Serotonin norepinephrine reuptake inhibitors)
- ☐ TCAs (Tricyclic antidepressants)
- ☐ TeCAs (Tetracyclic antidepressants)
- ☐ MAOIs (Monoamine oxidase inhibitors)
- ☐ Melatonin
- ☐ Agomelatine
- ☐ Lamotrigine
- ☐ Lithium
- ☐ Antipsychotics
- ☐ Stimulants
- ☐ Ketamine, Esketamine
- ☐ Psychedelics
- ☐ Medicinal cannabis
- ☐ None
- ☐ Other: ____________

##### Which of these medications are you <u>currently</u> taking?

Only answer this question if the following conditions are met:
((currentAndLastYrMeds_SQ001.NAOK == "Y")) or
((currentAndLastYrMeds_SQ002.NAOK == "Y")) or
((currentAndLastYrMeds_SQ003.NAOK == "Y")) or
((currentAndLastYrMeds_SQ004.NAOK == "Y")) or
((currentAndLastYrMeds_SQ005.NAOK == "Y")) or
((currentAndLastYrMeds_SQ006.NAOK == "Y")) or
((currentAndLastYrMeds_SQ007.NAOK == "Y")) or
((currentAndLastYrMeds_SQ008.NAOK == "Y")) or
((currentAndLastYrMeds_SQ009.NAOK == "Y")) or
((currentAndLastYrMeds_SQ010.NAOK == "Y")) or ((currentAndLastYrMeds_SQ011.NAOK == "Y")) or ((currentAndLastYrMeds_SQ012.NAOK == "Y")) or
((currentAndLastYrMeds_SQ013.NAOK == "Y")) or
((currentAndLastYrMeds_SQ014.NAOK == "Y"))

❗ Check all that apply
Please choose **all** that apply:

- ☐ {currentAndLastYrMeds_SQ001.question}
- ☐ {currentAndLastYrMeds_SQ002.question}
- ☐ {currentAndLastYrMeds_SQ003.question}
- ☐ {currentAndLastYrMeds_SQ004.question}
- ☐ {currentAndLastYrMeds_SQ005.question}
- ☐ {currentAndLastYrMeds_SQ006.question}
- ☐ {currentAndLastYrMeds_SQ007.question}
- ☐ {currentAndLastYrMeds_SQ008.question}
- ☐ {currentAndLastYrMeds_SQ009.question}
- ☐ {currentAndLastYrMeds_SQ010.question}
- ☐ {currentAndLastYrMeds_SQ011.question}
- ☐ {currentAndLastYrMeds_SQ012.question}
- ☐ {currentAndLastYrMeds_SQ013.question}
- ☐ {currentAndLastYrMeds_SQ014.question}
- ☐ None
- ☐ Other: ____________________

#### SECTION 5 (cont.): All Medications For Depression

**We would now like to ask about side effects associated with any of the medications you have ever taken. We will ask about one medication at a time. If medications have been taken together it may be hard to allocate the side effects to a single medication. Please answer to the best of your ability.**

| | Side Effect 1 | Side Effect 2 | Side Effect 3 | Side Effect 4 |
|---|---|---|---|---|
| {medPrescribedEver_SQ001.question} | | | | |
| {medPrescribedEver_SQ002.question} | | | | |
| {medPrescribedEver_SQ003.question} | | | | |
| {medPrescribedEver_SQ004.question} | | | | |
| {medPrescribedEver_SQ005.question} | | | | |
| {medPrescribedEver_SQ006.question} | | | | |
| {medPrescribedEver_SQ007.question} | | | | |
| {medPrescribedEver_SQ008.question} | | | | |
| {medPrescribedEver_SQ009.question} | | | | |
| {medPrescribedEver_SQ010.question} | | | | |
| {medPrescribedEver_SQ011.question} | | | | |
| {medPrescribedEver_SQ012.question} | | | | |
| {medPrescribedEver_SQ013.question} | | | | |
| {medPrescribedEver_SQ014.question} | | | | |

| |
|---|
| {medPrescribedEver_other} |

Select **up to** four side effects per treatment

##### If you have <u>any other side effects</u> that were not included in the lists above, please specify them here <u>along with the medication</u>.

Please write your answer here:

##### How long did you take (or have been taking) this medication (approximately)?

| | Years | Months | Weeks | Days |
|---|---|---|---|---|
| {medPrescribedEver_SQ001.question} | | | | |
| {medPrescribedEver_SQ002.question} | | | | |
| {medPrescribedEver_SQ003.question} | | | | |
| {medPrescribedEver_SQ004.question} | | | | |
| {medPrescribedEver_SQ005.question} | | | | |
| {medPrescribedEver_SQ006.question} | | | | |

| |
|---|
| **{medPrescribedEver_SQ007.question}** |
| **{medPrescribedEver_SQ008.question}** |
| **{medPrescribedEver_SQ009.question}** |
| **{medPrescribedEver_SQ010.question}** |
| **{medPrescribedEver_SQ011.question}** |
| **{medPrescribedEver_SQ012.question}** |
| **{medPrescribedEver_SQ013.question}** |
| **{medPrescribedEver_SQ014.question}** |
| **{medPrescribedEver_other}** |

Choose the most suitable time units and fill in the corresponding column - you do not need to fill in every column.

#### If you stopped taking this medication, why did you stop taking this medication? We are displaying the list of medication you have tried but are not currently taking.

Please choose the appropriate response for each item:

| | I started feeling better | I experienced side effects | My symptoms were unchanged - it was not working for me | It cost too much | My prescription was changed | Something else / Other | Don't Know |
|---|---|---|---|---|---|---|---|
| {medPrescribedEver_SQ001.question} | ◯ | ◯ | ◯ | ◯ | ◯ | ◯ | ◯ |
| {medPrescribedEver_SQ002.question} | ◯ | ◯ | ◯ | ◯ | ◯ | ◯ | ◯ |
| {medPrescribedEver_SQ003.question} | ◯ | ◯ | ◯ | ◯ | ◯ | ◯ | ◯ |
| {medPrescribedEver_SQ004.question} | ◯ | ◯ | ◯ | ◯ | ◯ | ◯ | ◯ |
| {medPrescribedEver_SQ005.question} | ◯ | ◯ | ◯ | ◯ | ◯ | ◯ | ◯ |
| {medPrescribedEver_SQ006.question} | ◯ | ◯ | ◯ | ◯ | ◯ | ◯ | ◯ |
| {medPrescribedEver_SQ007.question} | ◯ | ◯ | ◯ | ◯ | ◯ | ◯ | ◯ |
| {medPrescribedEver_SQ008.question} | ◯ | ◯ | ◯ | ◯ | ◯ | ◯ | ◯ |
| {medPrescribedEver_SQ009.question} | ◯ | ◯ | ◯ | ◯ | ◯ | ◯ | ◯ |
| {medPrescribedEver_SQ010.question} | ◯ | ◯ | ◯ | ◯ | ◯ | ◯ | ◯ |
| {medPrescribedEver_SQ011.question} | ◯ | ◯ | ◯ | ◯ | ◯ | ◯ | ◯ |
| {medPrescribedEver_SQ012.question} | ◯ | ◯ | ◯ | ◯ | ◯ | ◯ | ◯ |
| {medPrescribedEver_SQ013.question} | ◯ | ◯ | ◯ | ◯ | ◯ | ◯ | ◯ |
| {medPrescribedEver_SQ014.question} | ◯ | ◯ | ◯ | ◯ | ◯ | ◯ | ◯ |
| {medPrescribedEver_other} | ◯ | ◯ | ◯ | ◯ | ◯ | ◯ | ◯ |

#### Did any of the below medications you have ever tried gave fast relief from depression (within the first 2 weeks)?

❶ Check all that apply
Please choose **all** that apply:

- ☐ {medPrescribedEver_SQ001.question}
- ☐ {medPrescribedEver_SQ002.question}
- ☐ {medPrescribedEver_SQ003.question}
- ☐ {medPrescribedEver_SQ004.question}
- ☐ {medPrescribedEver_SQ005.question}
- ☐ {medPrescribedEver_SQ006.question}
- ☐ {medPrescribedEver_SQ007.question}
- ☐ {medPrescribedEver_SQ008.question}
- ☐ {medPrescribedEver_SQ009.question}
- ☐ {medPrescribedEver_SQ010.question}
- ☐ {medPrescribedEver_SQ011.question}
- ☐ {medPrescribedEver_SQ012.question}
- ☐ {medPrescribedEver_SQ013.question}
- ☐ {medPrescribedEver_SQ014.question}
- ☐ {medPrescribedEver_other}
- ☐ None

#### What was the first symptom you noticed where the medication provided fast relief? Here is the list of depressive symptoms you reported you experience when you are depressed.

Only answer this question if the following conditions are met:
0

❶ Please select one answer
Please choose **all** that apply:

- ☐ {if(moodSymptoms_SQ001=="AO01", moodSymptoms_SQ001.question)}
- ☐ {if(moodSymptoms_SQ002=="AO01", moodSymptoms_SQ002.question)}

- ☐ {if(moodSymptoms_SQ003=="AO01", moodSymptoms_SQ003.question)}
- ☐ {if(moodSymptoms_SQ004=="AO01", moodSymptoms_SQ004.question)}
- ☐ {if(moodSymptoms_SQ006=="AO01", moodSymptoms_SQ006.question)}
- ☐ {if(moodSymptoms_SQ007=="AO01", moodSymptoms_SQ007.question)}
- ☐ {if(moodSymptoms_SQ008=="AO01", moodSymptoms_SQ008.question)}
- ☐ {if(moodSymptoms_SQ005=="AO01", moodSymptoms_SQ005.question)}
- ☐ {if(anxietySymptoms_SQ001=="AO01", anxietySymptoms_SQ001.question)}
- ☐ {if(anxietySymptoms_SQ006=="AO01", anxietySymptoms_SQ006.question)}
- ☐ {if(anxietySymptoms_SQ007=="AO01", anxietySymptoms_SQ007.question)}
- ☐ {if(anxietySymptoms_SQ008=="AO01", anxietySymptoms_SQ008.question)}
- ☐ {if(cognitiveSymptoms_SQ001=="AO01", cognitiveSymptoms_SQ001.question)}
- ☐ {if(cognitiveSymptoms_SQ002=="AO01", cognitiveSymptoms_SQ002.question)}
- ☐ {if(anxietySymptoms_SQ002=="AO01", anxietySymptoms_SQ002.question)}
- ☐ {if(anxietySymptoms_SQ003=="AO01", anxietySymptoms_SQ003.question)}
- ☐ {if(anxietySymptoms_SQ004=="AO01", anxietySymptoms_SQ004.question)}
- ☐ {if(anxietySymptoms_SQ005=="AO01", anxietySymptoms_SQ005.question)}
- ☐ {if(cognitiveSymptoms_SQ003=="AO01", cognitiveSymptoms_SQ003.question)}
- ☐ {if(cognitiveSymptoms_SQ004=="AO01", cognitiveSymptoms_SQ004.question)}
- ☐ {if(cognitiveSymptoms_SQ005=="AO01", cognitiveSymptoms_SQ005.question)}
- ☐ {if(cognitiveSymptoms_SQ006=="AO01", cognitiveSymptoms_SQ006.question)}
- ☐ {if(cognitiveSymptoms_SQ007=="AO01", cognitiveSymptoms_SQ007.question)}
- ☐ {if(cognitiveSymptoms_SQ008=="AO01", cognitiveSymptoms_SQ008.question)}
- ☐ {if(cognitiveSymptoms_SQ009=="AO01", cognitiveSymptoms_SQ009.question)}
- ☐ {if(cognitiveSymptoms_SQ010=="AO01", cognitiveSymptoms_SQ010.question)}
- ☐ {if(sleepSymptoms_SQ001=="AO01", sleepSymptoms_SQ001.question)}
- ☐ {if(sleepSymptoms_SQ007=="AO01", sleepSymptoms_SQ007.question)}
- ☐ {if(sleepSymptoms_SQ008=="AO01", sleepSymptoms_SQ008.question)}
- ☐ {if(sleepSymptoms_SQ003=="AO01", sleepSymptoms_SQ003.question)}
- ☐ {if(sleepSymptoms_SQ002=="AO01", sleepSymptoms_SQ002.question)}
- ☐ {if(sleepSymptoms_SQ004=="AO01", sleepSymptoms_SQ004.question)}
- ☐ {if(sleepSymptoms_SQ005=="AO01", sleepSymptoms_SQ005.question)}
- ☐ {if(sleepSymptoms_SQ006=="AO01", sleepSymptoms_SQ006.question)}
- ☐ {if(behaviouralSymptoms_SQ001=="AO01", behaviouralSymptoms_SQ001.question)}
- ☐ {if(behaviouralSymptoms_SQ009=="AO01",

behaviouralSymptoms_SQ009.question)}

- ☐ {if(behaviouralSymptoms_SQ002=="AO01", behaviouralSymptoms_SQ002.question)}
- ☐ {if(behaviouralSymptoms_SQ003=="AO01", behaviouralSymptoms_SQ003.question)}
- ☐ {if(behaviouralSymptoms_SQ004=="AO01", behaviouralSymptoms_SQ004.question)}
- ☐ {if(behaviouralSymptoms_SQ005=="AO01", behaviouralSymptoms_SQ005.question)}
- ☐ {if(behaviouralSymptoms_SQ006=="AO01", behaviouralSymptoms_SQ006.question)}
- ☐ {if(behaviouralSymptoms_SQ007=="AO01", behaviouralSymptoms_SQ007.question)}
- ☐ {if(behaviouralSymptoms_SQ010=="AO01", behaviouralSymptoms_SQ010.question)}
- ☐ {if(behaviouralSymptoms_SQ008=="AO01", behaviouralSymptoms_SQ008.question)}
- ☐ {if(physicalSymptoms_SQ014=="AO01", physicalSymptoms_SQ014.question)}
- ☐ {if(physicalSymptoms_SQ001=="AO01", physicalSymptoms_SQ001.question)}
- ☐ {if(physicalSymptoms_SQ002=="AO01", physicalSymptoms_SQ002.question)}
- ☐ {if(physicalSymptoms_SQ003=="AO01", physicalSymptoms_SQ003.question)}
- ☐ {if(physicalSymptoms_SQ004=="AO01", physicalSymptoms_SQ004.question)}
- ☐ {if(physicalSymptoms_SQ005=="AO01", physicalSymptoms_SQ005.question)}
- ☐ {if(physicalSymptoms_SQ006=="AO01", physicalSymptoms_SQ006.question)}
- ☐ {if(physicalSymptoms_SQ007=="AO01", physicalSymptoms_SQ007.question)}
- ☐ {if(physicalSymptoms_SQ008=="AO01", physicalSymptoms_SQ008.question)}
- ☐ {if(physicalSymptoms_SQ009=="AO01", physicalSymptoms_SQ009.question)}
- ☐ {if(physicalSymptoms_SQ010=="AO01", physicalSymptoms_SQ010.question)}
- ☐ {if(physicalSymptoms_SQ012=="AO01", physicalSymptoms_SQ012.question)}
- ☐ {if(physicalSymptoms_SQ013=="AO01", physicalSymptoms_SQ013.question)}
- ☐ {if(physicalSymptoms_SQ011=="AO01", physicalSymptoms_SQ011.question)}
- ☐ Other: ________

#### This image presents an overview of symptoms people may experience during depression. Think about your set of personal symptoms in each area when answering the questions below.

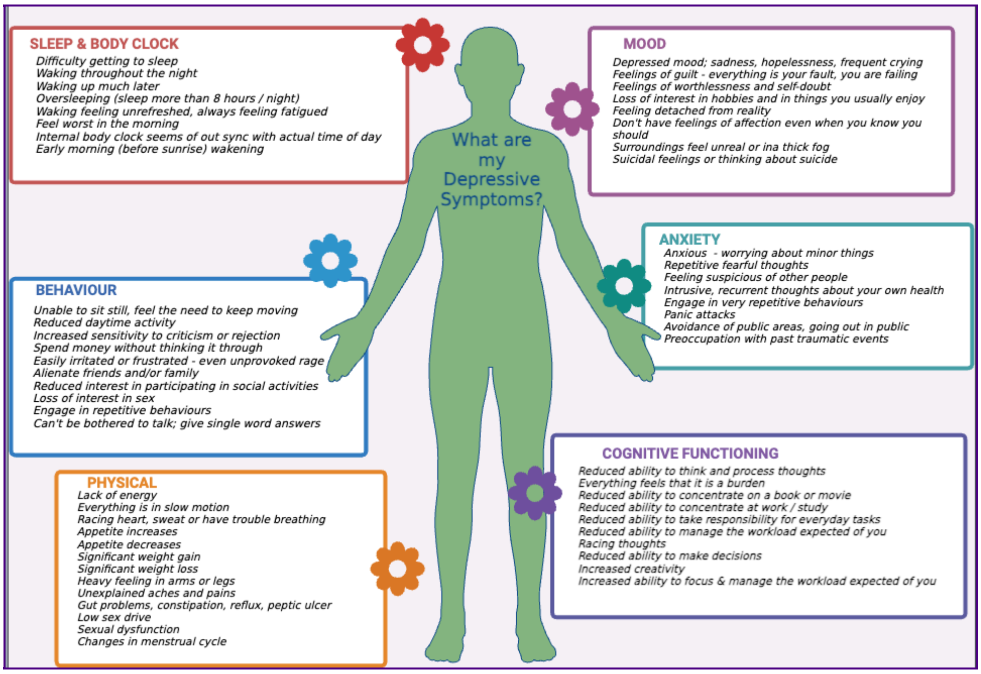

#### What was the <u>first area of symptoms you noticed</u> where the medication provided fast relief?

#### Here is the list of area of symptoms you reported you experience when you are depressed.

Only answer this question if the following conditions are met:
((is_empty(fastRelief_SQ031.NAOK)))

❗ Please select one answer
Please choose **all** that apply:

- ☐ Mood
- ☐ Anxiety
- ☐ Cognitive functioning
- ☐ Sleep pattern and body clock
- ☐ Behaviour
- ☐ Physical

#### What was this first symptom where the medication provided fast relief?

Only answer this question if the following conditions are met:
((is_empty(fastRelief_SQ031.NAOK)))

Please write your answer here:

#### On starting any of the medications you have ever taken, did you experience any side effects within the first two weeks?

Please choose **only one** of the following:

- Yes
- No

#### Please select all the side effects, you have experienced within the first two weeks when you started taking the below medication/s. We will ask about one medication at a time. If medications have been taken together it may be hard to allocate the side effects to a single medication. Please answer to the best of your ability.

Only answer this question if the following conditions are met:
((sideEffctsFrst2Weeks.NAOK == "Y"))

| | Side Effect 1 | Side Efect 2 | Side Effect 3 | Side Effect 4 |
|---|---|---|---|---|
| {medPrescribedEver_SQ001.question} | | | | |
| {medPrescribedEver_SQ002.question} | | | | |
| {medPrescribedEver_SQ003.question} | | | | |

| |
|---|
| {medPrescribedEver_SQ004.question} |
| {medPrescribedEver_SQ005.question} |
| {medPrescribedEver_SQ006.question} |
| {medPrescribedEver_SQ007.question} |
| {medPrescribedEver_SQ008.question} |
| {medPrescribedEver_SQ009.question} |
| {medPrescribedEver_SQ010.question} |
| {medPrescribedEver_SQ011.question} |
| {medPrescribedEver_SQ012.question} |
| {medPrescribedEver_SQ013.question} |
| {medPrescribedEver_SQ014.question} |
| {medPrescribedEver_other} |
| {currentAndLastYrMeds_SQ016.question} |
| {currentAndLastYrMeds_SQ017.question} |
| {currentAndLastYrMeds_SQ018.question} |
| {currentAndLastYrMeds_SQ019.question} |
| {currentAndLastYrMeds_SQ020.question} |

| |
|---|
| {currentAndLastYrMeds_SQ021.question} |
| {currentAndLastYrMeds_SQ022.question} |
| {currentAndLastYrMeds_SQ023.question} |
| {currentAndLastYrMeds_SQ024.question} |
| {currentAndLastYrMeds_SQ025.question} |
| {currentAndLastYrMeds_SQ026.question} |
| {currentAndLastYrMeds_SQ027.question} |
| {currentAndLastYrMeds_SQ028.question} |
| {currentAndLastYrMeds_SQ029.question} |
| {currentAndLastYrMeds_other} |

**If you have <u>any other side effects</u> that were not included in the lists above, please specify them here <u>along with the medication</u>.**

Only answer this question if the following conditions are met:
((sideEffctsFrst2Weeks.NAOK == "Y"))

Please write your answer here:

**Did you ever experience such a <u>severe reaction</u> to one of these antidepression medications that you had to stop the drug <u>within the first two weeks</u> of taking it?**

Please choose **only one** of the following:

- ◯ Yes
- ◯ No

#### Which of the below medications did you have to stop because you experienced a severe reaction within the first two weeks of taking it?

Only answer this question if the following conditions are met:
((severeReaction.NAOK == "Y"))

❗ Check all that apply
Please choose **all** that apply:

- ☐ {medPrescribedEver_SQ001.question}
- ☐ {medPrescribedEver_SQ002.question}
- ☐ {medPrescribedEver_SQ003.question}
- ☐ {medPrescribedEver_SQ004.question}
- ☐ {medPrescribedEver_SQ005.question}
- ☐ {medPrescribedEver_SQ006.question}
- ☐ {medPrescribedEver_SQ007.question}
- ☐ {medPrescribedEver_SQ008.question}
- ☐ {medPrescribedEver_SQ009.question}
- ☐ {medPrescribedEver_SQ010.question}
- ☐ {medPrescribedEver_SQ011.question}
- ☐ {medPrescribedEver_SQ012.question}
- ☐ {medPrescribedEver_SQ013.question}
- ☐ {medPrescribedEver_SQ014.question}
- ☐ {medPrescribedEver_other}
- ☐ Other: ________

#### For any of the medication you have ever tried did you experience any side effects after the first month of taking it?

Please choose **only one** of the following:

- ◯ Yes
- ◯ No

#### Please select all the side effects, you have experienced after the first month of taking the below medication/s. We will ask about one medication at a time. If medications have been taken together it may be hard to allocate the side effects to a single medication. Please answer to the best of your ability.

Only answer this question if the following conditions are met:
((sideEffctsFirstMonth.NAOK == "Y"))

| | Side Effect 1 | Side Effect 2 | Side Effect 3 | Side Effect 4 |
|---|---|---|---|---|
| {medPrescribedEver_SQ001.question} | | | | |
| {medPrescribedEver_SQ002.question} | | | | |
| {medPrescribedEver_SQ003.question} | | | | |
| {medPrescribedEver_SQ004.question} | | | | |
| {medPrescribedEver_SQ005.question} | | | | |
| {medPrescribedEver_SQ006.question} | | | | |
| {medPrescribedEver_SQ007.question} | | | | |
| {medPrescribedEver_SQ008.question} | | | | |

| |
|---|
| **{medPrescribedEver_SQ009.question}** |
| **{medPrescribedEver_SQ010.question}** |
| **{medPrescribedEver_SQ011.question}** |
| **{medPrescribedEver_SQ012.question}** |
| **{medPrescribedEver_SQ013.question}** |
| **{medPrescribedEver_SQ014.question}** |
| **{medPrescribedEver_other}** |

#### If you have <u>any other side effects</u> that were not included in the lists above, please specify them here <u>along with the medication</u>.

Only answer this question if the following conditions are met:
((sideEffctsFirstMonth.NAOK == "Y"))

Please write your answer here:

**Do you feel strongly that side effects were associated with a specific brand of antidepressant? If so, please describe.**

Please write your answer here:

**Lastly, current medications can impact measures of biomarkers measured in your blood sample. Please list all the medications you are currently taking for any reason. Please add 'None' if you are not taking any medications.**

Please write your answer here:

**If you prefer, you can photograph the medication label of the medications you are currently taking and upload it here.**

❗ Please upload at most 15 files
Kindly attach the aforementioned documents along with the survey

You can upload any number of files

### Australian Genetics of Depression Study: Anti-depressant Cell-omics

There are 103 questions in this survey.

#### SECTION 6A: Menstruation and Pregnancy

Symptoms of depression can be impacted by changes in a female’s menstrual cycle and the use of hormonal medications. This next set of questions asks about contraceptive use, menstruation, pregnancy and birth and their impact on your depression. Answering these questions may involve recalling information about miscarriage and loss.

##### Please confirm if you are comfortable answering the following set of questions pertaining to menstruation and pregnancy.

Please choose **only one** of the following:

- ◯ Happy to complete
- ◯ Skip
- ◯ Not applicable to me

#### SECTION 6A: Menstruation and Pregnancy

*In this section, there are 35 questions, and depending on your responses, this can increase. We anticipate that this section should take no longer than 10 minutes to complete.*

##### Have you ever taken hormonal contraceptives?

Please choose **only one** of the following:

- ◯ ✔Yes
- ◯ ⊘No
- ◯ ━Prefer not to say

#### Which of the following hormonal contraceptives have you ever taken?

Please choose **all** that apply:

- ☐ Oral birth control pill (unsure of the type)
- ☐ Combined (oestrogen and progestin) oral birth control pill
- ☐ Progestin only oral birth control pill ("mini-pill")
- ☐ Transdermal (patches)
- ☐ Vaginal ring
- ☐ Progestin containing Coil/IUD
- ☐ Hormonal Implant
- ☐ Gonadotropin (GnRH) analogue

#### At what age did you start taking hormonal contraceptives?

Please write your answer here:

#### Are you currently taking hormonal contraceptives?

Please choose **only one** of the following:

- ○ Yes
- ○ No

#### Which of the following hormonal contraceptives are you currently taking?

Please choose **all** that apply:

- ☐ Oral birth control pill (unsure of the type)
- ☐ Combined (oestrogen and progestin) oral birth control pill
- ☐ Progestin only oral birth control pill ("mini-pill")
- ☐ Transdermal (patches)
- ☐ Vaginal ring
- ☐ Progestin containing Coil/IUD
- ☐ Hormonal Implant
- ☐ Gonadotropin (GnRH) analogue

#### What is the name of the oral birth control pill you are currently taking?

Please write your answer here:

#### From your experience, have you tried any hormonal contraceptives that have made your symptoms of depression worse?

Please choose **only one** of the following:

- ○ Yes
- ○ No

#### Which contraceptive(s) make your depressive symptoms worse?

Please choose **all** that apply:

- ☐ Oral birth control pill (unsure of the type)
- ☐ Combined (oestrogen and progestin) oral birth control pill
- ☐ Progestin only oral birth control pill ("mini-pill")
- ☐ Transdermal (patches) Vaginal ring
- ☐ Progestin containing Coil/IUD
- ☐ Hormonal Implant
- ☐ Gonadotropin (GnRH) analogue

#### If you know the drug name please detail below:

Please write your answer here:

#### This image presents an overview of symptoms people may experience during depression. Think about your set of personal symptoms in each area when answering the questions below.

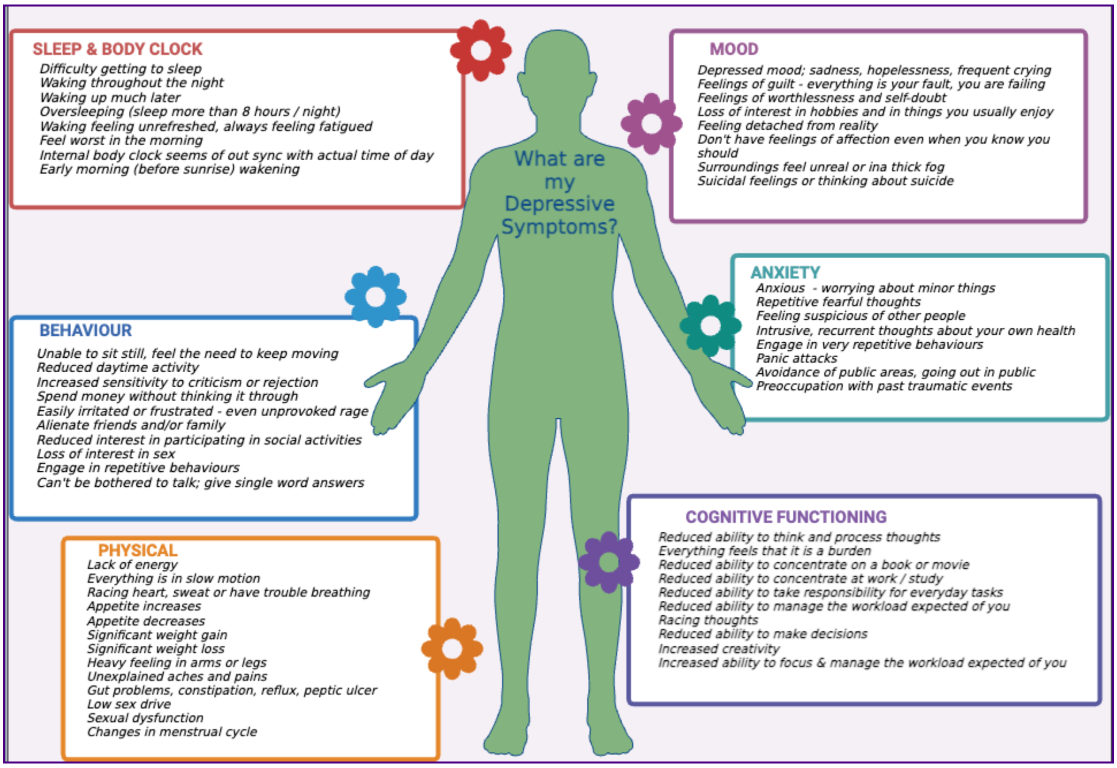

#### Thinking about your personal set of depressive symptoms which symptom areas of depression are worsened by the hormonal contraceptives?

Please choose **all** that apply:

- ☐ Mood
- ☐ Anxiety
- ☐ Cognitive functioning
- ☐ Sleep
- ☐ Behavioural
- ☐ Physical

**These next questions will ask you about information related to your menstruation history and cycle in relation to your depression.**

#### Have you started menopause?

Please choose **only one** of the following:

- ◯ ✔Yes
- ◯ ⊘No
- ◯ ?Unsure
- ◯ ━Prefer not to say

#### At what age did you have your last period?

Please write your answer here:

#### Following menopause, did your symptoms of depression:

Please choose **only one** of the following:

- ◯ Get better
- ◯ Get worse
- ◯ No change

#### At what age did you have your first period?

Please write your answer here:

#### From your experience, is there a particular stage of your menstrual cycle that makes your symptoms of depression worse?

Please choose **only one** of the following:

- ◯ Yes
- ◯ No

#### Please select the stage of your cycle that makes your symptoms of depression worse?

Please choose **all** that apply:

- ☐ More than 2 days before a period
- ☐ On the days immediately before a period
- ☐ On the first day of a period
- ☐ Mid cycle (during ovulation)
- ☐ Within a few days after a period
- ☐ Throughout the entire period
- ☐ Uncertain

**This image presents an overview of symptoms people may experience during depression. Think about your set of personal symptoms in each area when answering the questions below.**

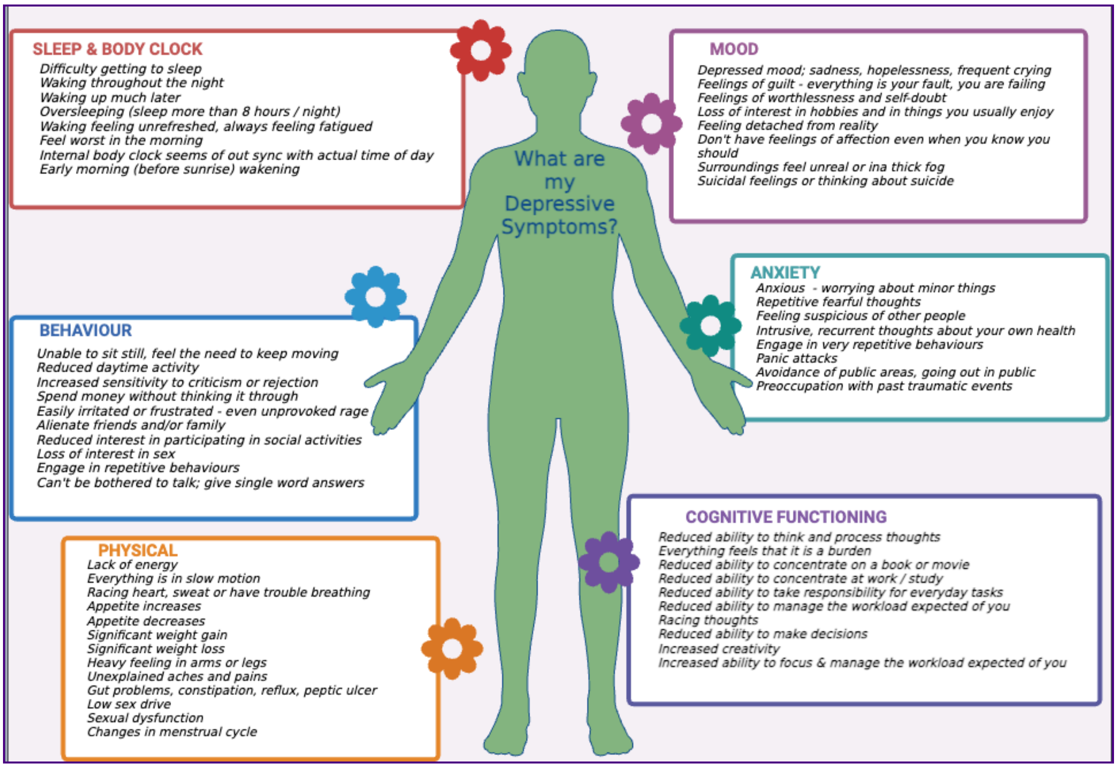

**Thinking about your personal set of depressive symptoms and any impact on your menstrual cycle; - where there are changes in your depressive symptoms identify which areas of symptoms are affected and rate their impact.**

#### More than 2 days before a period:

Please choose the appropriate response for each item:

| | 1<br>No Impact | 2<br>Some Impact | 3<br>Moderate Impact | 4<br>Major Impact | 5<br>Severe Impact |
|---|---|---|---|---|---|
| **Mood** | ○ | ○ | ○ | ○ | ○ |
| **Anxiety** | ○ | ○ | ○ | ○ | ○ |
| **Sleep** | ○ | ○ | ○ | ○ | ○ |
| **Cognitive functioning** | ○ | ○ | ○ | ○ | ○ |
| **Behavioural** | ○ | ○ | ○ | ○ | ○ |
| **Physical** | ○ | ○ | ○ | ○ | ○ |

#### On the days immediately before a period:

Please choose the appropriate response for each item:

| | 1<br>No Impact | 2<br>Some Impact | 3<br>Moderate Impact | 4<br>Major Impact | 5<br>Severe Impact |
|---|---|---|---|---|---|
| **Mood** | ○ | ○ | ○ | ○ | ○ |
| **Anxiety** | ○ | ○ | ○ | ○ | ○ |
| **Sleep** | ○ | ○ | ○ | ○ | ○ |
| **Cognitive functioning** | ○ | ○ | ○ | ○ | ○ |
| **Behavioural** | ○ | ○ | ○ | ○ | ○ |
| **Physical** | ○ | ○ | ○ | ○ | ○ |

#### On the first day of a period:

Please choose the appropriate response for each item:

| | 1 No Impact | 2 Some Impact | 3 Moderate Impact | 4 Major Impact | 5 Severe Impact |
|---|---|---|---|---|---|
| **Mood** | ◯ | ◯ | ◯ | ◯ | ◯ |
| **Anxiety** | ◯ | ◯ | ◯ | ◯ | ◯ |
| **Sleep** | ◯ | ◯ | ◯ | ◯ | ◯ |
| **Cognitive functioning** | ◯ | ◯ | ◯ | ◯ | ◯ |
| **Behavioural** | ◯ | ◯ | ◯ | ◯ | ◯ |
| **Physical** | ◯ | ◯ | ◯ | ◯ | ◯ |

#### Mid cycle (during ovulation)

Please choose the appropriate response for each item:

| | 1 No Impact | 2 Some Impact | 3 Moderate Impact | 4 Major Impact | 5 Severe Impact |
|---|---|---|---|---|---|
| **Mood** | ◯ | ◯ | ◯ | ◯ | ◯ |
| **Anxiety** | ◯ | ◯ | ◯ | ◯ | ◯ |
| **Sleep** | ◯ | ◯ | ◯ | ◯ | ◯ |
| **Cognitive functioning** | ◯ | ◯ | ◯ | ◯ | ◯ |
| **Behavioural** | ◯ | ◯ | ◯ | ◯ | ◯ |
| **Physical** | ◯ | ◯ | ◯ | ◯ | ◯ |

#### Within a few days after a period:

Please choose the appropriate response for each item:

| | 1 No Impact | 2 Some Impact | 3 Moderate Impact | 4 Major Impact | 5 Severe Impact |
|---|---|---|---|---|---|
| **Mood** | ○ | ○ | ○ | ○ | ○ |
| **Anxiety** | ○ | ○ | ○ | ○ | ○ |
| **Sleep** | ○ | ○ | ○ | ○ | ○ |
| **Cognitive functioning** | ○ | ○ | ○ | ○ | ○ |
| **Behavioural** | ○ | ○ | ○ | ○ | ○ |
| **Physical** | ○ | ○ | ○ | ○ | ○ |

#### Throughout the entire period:

Please choose the appropriate response for each item:

| | 1 No Impact | 2 Some Impact | 3 Moderate Impact | 4 Major Impact | 5 Severe Impact |
|---|---|---|---|---|---|
| **Mood** | ○ | ○ | ○ | ○ | ○ |
| **Anxiety** | ○ | ○ | ○ | ○ | ○ |
| **Sleep** | ○ | ○ | ○ | ○ | ○ |
| **Cognitive functioning** | ○ | ○ | ○ | ○ | ○ |
| **Behavioural** | ○ | ○ | ○ | ○ | ○ |
| **Physical** | ○ | ○ | ○ | ○ | ○ |

**These next questions will ask you about your pregnancy history and reproductive health in relation to your depression.**

#### Have you ever fallen pregnant?

Please choose **only one** of the following:

- ◯ ✔Yes
- ◯ ⊘No
- ◯ ━Prefer not to say

#### How many times have you fallen pregnant?

Please write your answer here:

#### During your pregnancy, did your symptoms of depression get worse?

Please choose **only one** of the following:

- ◯ ✔Yes
- ◯ ⊘No
- ◯ ━Prefer not to say

#### Thinking about your personal set of depressive symptoms which symptom areas were worsened during your pregnancy?

Please choose **all** that apply:

- ☐ Mood
- ☐ Anxiety
- ☐ Cognitive functioning
- ☐ Sleep
- ☐ Behavioural
- ☐ Physical

#### Please rate the impact of pregnancy had of each of the symptom areas.

Please choose the appropriate response for each item:

| | 1 No Impact | 2 Some Impact | 3 Moderate Impact | 4 Major Impact | 5 Severe Impact |
|---|---|---|---|---|---|
| {if(pregSympt_SQ001=="Y", pregSympt_SQ001.question)} | ○ | ○ | ○ | ○ | ○ |
| {if(pregSympt_SQ002=="Y", pregSympt_SQ002.question)} | ○ | ○ | ○ | ○ | ○ |
| {if(pregSympt_SQ003=="Y", pregSympt_SQ003.question)} | ○ | ○ | ○ | ○ | ○ |
| {if(pregSympt_SQ004=="Y", pregSympt_SQ004.question)} | ○ | ○ | ○ | ○ | ○ |
| {if(pregSympt_SQ005=="Y", pregSympt_SQ005.question)} | ○ | ○ | ○ | ○ | ○ |
| {if(pregSympt_SQ006=="Y", pregSympt_SQ006.question)} | ○ | ○ | ○ | ○ | ○ |

#### If you have given birth, did your symptoms of depression get worse after giving birth?

Please choose **only one** of the following:

- ◯ ✔Yes
- ◯ ⊘No
- ◯ ━Prefer not to say

#### What symptom areas were impacted during the post-partum period (3 months after birth)?

Please choose **all** that apply:

- ☐ Mood
- ☐ Anxiety
- ☐ Sleep
- ☐ Cognitive functioning
- ☐ Behavioural
- ☐ Physical

#### Please rate the impact on each of the symptom areas.

Please choose the appropriate response for each item:

| | 1 No Impact | 2 Some Impact | 3 Moderate Impact | 4 Major Impact | 5 Severe Impact |
|---|---|---|---|---|---|
| **{if(birthSympt_SQ001=="Y", birthSympt_SQ001.question)}** | ◯ | ◯ | ◯ | ◯ | ◯ |
| **{if(birthSympt_SQ002=="Y", birthSympt_SQ002.question)}** | ◯ | ◯ | ◯ | ◯ | ◯ |
| **{if(birthSympt_SQ003=="Y", birthSympt_SQ003.question)}** | ◯ | ◯ | ◯ | ◯ | ◯ |
| **{if(birthSympt_SQ004=="Y", birthSympt_SQ004.question)}** | ◯ | ◯ | ◯ | ◯ | ◯ |
| **{if(birthSympt_SQ005=="Y", birthSympt_SQ005.question)}** | ◯ | ◯ | ◯ | ◯ | ◯ |
| **{if(birthSympt_SQ006=="Y", birthSympt_SQ006.question)}** | ◯ | ◯ | ◯ | ◯ | ◯ |

#### Have you experienced infertility?

Please choose **only one** of the following:

- ◯ ✔Yes
- ◯ ⊘No
- ◯ ?Unsure
- ◯ ━Prefer not to say

#### Have you ever received treatment for infertility?

Please choose **only one** of the following:

- ◯ ✔Yes
- ◯ ⊘No
- ◯ ━Prefer not to say

#### Have you had a hysterectomy (removal of uterus)?

Please choose **only one** of the following:

- ◯ ✔Yes
- ◯ ⊘No
- ◯ ━Prefer not to say

#### Were your ovaries removed?

Please choose **only one** of the following:

- ◯ Yes, both
- ◯ Yes, one side
- ◯ No

#### What age were you when your ovary(s) were removed?

Please write your answer here:

#### Following your hysterectomy, did your symptoms of depression:

Please choose **only one** of the following:

- ◯ Get better
- ◯ Get worse
- ◯ No change

#### In which domain did you see this improvement in depression symptoms?

Please choose **all** that apply:

- ☐ Mood
- ☐ Anxiety
- ☐ Cognitive functioning
- ☐ Sleep
- ☐ Behavioural
- ☐ Physical

#### Have you ever taken Hormone Replacement Therapy (HRT)?

Please choose **only one** of the following:

- ◯ ✔Yes
- ◯ ⊘No
- ◯ ━Prefer not to say

##### If yes, are you currently taking HRT?

Please choose **only one** of the following:

- ◯ Yes
- ◯ No

##### In your own words, is there anything you would like to say about your experience with menstruation, contraceptives, or pregnancy in relation to your depression?

Please write your answer here:

### SECTION 6: My Mental Health Journey

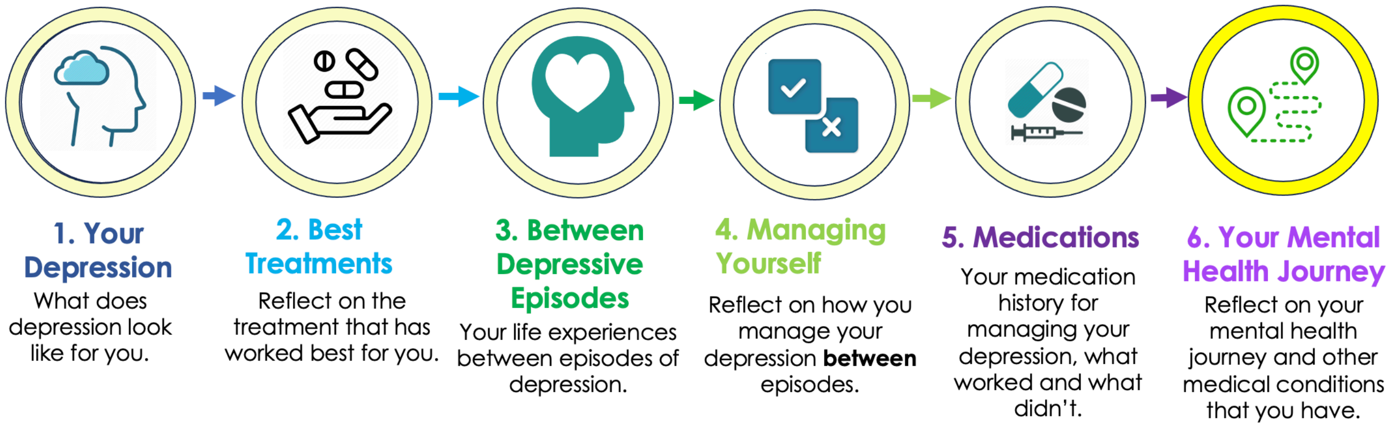

This is the final section of this questionnaire and thank you for progressing through the previous sections. In this section we would like to ask about any other mental health conditions you have been diagnosed with in addition to Major Depressive Disorder. This information helps to inform us about your personal mental health journey and health experiences over your lifetime.

*In this section, there are 30 questions, and depending on your responses, this can increase. We anticipate that this section should take no longer than 10-12 minutes to complete.*

#### Have you been diagnosed with any of the following over the course of your lifetime?

| | Diagnosis | Age in years |
|---|---|---|
| **Depression** | | |
| **Anxiety** | | |
| **Obsessive compulsive disorder** | | |
| **Bipolar** | | |
| **Schizophrenia** | | |
| **Trauma and stress-related disorders** | | |
| **Autism spectrum disorder** | | |
| **Attention deficit/hyperactivity disorder** | | |
| **Substance use disorders** | | |
| **Major eating difficulties** | | |
| **Major sleep difficulties** | | |
| **Premenstrual Dysphoric Disorder** | | |

**The next set of questions will probe a bit further about how you experience depression and what other factors if any, you may experience. This additional information is important in understanding how your mental health journey may be different to another person who has depression. We know that a depression diagnosis is different for different people and by collecting this data we hope to gain a better understanding of the diversity of this disorder in the community.**

#### Did you experience any symptoms of depression prior to your diagnosis?

Please choose **only one** of the following:

- ◯ Yes
- ◯ No

#### How old were you when you first started experiencing symptoms of depression?

Please write your answer here:

#### How old were you when you were first given a diagnosis of depression?

Please write your answer here:

**Did you experience anxiety <u>before</u> your diagnosis of depression?**

Please choose **only one** of the following:

- ◯ Yes
- ◯ No

**Does your depression tend to switch ‘on’ and then ‘off’ in ways that appear to be unrelated to life stressors?**

Please choose **only one** of the following:

- ◯ Yes
- ◯ No

**Do you have periods where your mood or energy levels fluctuates significantly? *i.e., periods of depressed mood, low energy, and oversleeping are replaced by periods of high mood, lots of energy and a decreased need for sleep.***

Please choose **only one** of the following:

- ◯ Yes
- ◯ No

#### Does this mood or energy level fluctuation occur over a:

Please choose **only one** of the following:

- ◯ Course of a day
- ◯ Course of a week
- ◯ Course of a month
- ◯ Unsure

#### Has this occurred more than once over the course of your depression?

Please choose **only one** of the following:

- ◯ Yes
- ◯ No

#### Does your mood or energy state fluctuate significantly in relation to changes in seasons?

Please choose **only one** of the following:

- ◯ Yes
- ◯ No

#### If yes, at what time of year does the depression or low energy state come on?

Please choose **all** that apply:

- ☐ Summer
- ☐ Autumn
- ☐ Winter
- ☐ Spring

#### Do you ever experience psychotic symptoms when you are depressed?

Please choose **only one** of the following:

- ◯ ✔Yes
- ◯ ⊘No
- ◯ ?Unsure
- ◯ ━Prefer not to say

For example: hearing or seeing things that other people can't, believing people are trying to harm you etc.

#### Do these psychotic symptoms include:

Please choose **all** that apply:

- ☐ Hearing or seeing things that other people can’t
- ☐ Believing people are trying to harm you
- ☐ Other: ____________________

#### You selected that you have been diagnosed with an anxiety disorder. Please select all anxiety disorders that you have been diagnosed with over the course of your lifetime:

Please choose **all** that apply:

- ☐ Generalised Anxiety Disorder
- ☐ Social Anxiety Disorder
- ☐ Separation Anxiety Disorder
- ☐ Panic Disorder
- ☐ Specific phobia
- ☐ Agoraphobia
- ☐ Selective mutism

#### What age did you first experience symptoms of an anxiety disorder?

Please write your answer here:

#### What age did you first experience symptoms of OCD?

Please write your answer here:

#### Please select the bipolar disorder that you have been diagnosed with:

Please choose **only one** of the following:

- ◯ Bipolar type 1
- ◯ Bipolar type 2
- ◯ Cyclothymic disorder
- ◯ Bipolar disorder not otherwise specified
- ◯ Unsure of the type

#### What age did you first experience symptoms of bipolar disorder?

Please write your answer here:

#### What age did you first experience symptoms of schizophrenia?

Please write your answer here:

#### Please select the trauma and stressor related disorders that you have been diagnosed with:

Please choose **all** that apply:

- ☐ Post-traumatic stress disorder (PTSD)
- ☐ Acute stress disorder
- ☐ Prolonged grief disorder
- ☐ Adjustment disorder

#### What age did you first experience symptoms of Autism Spectrum Disorder?

Please write your answer here:

#### What age did you first experience symptoms of this trauma and stressor related disorder?

Please write your answer here:

#### Please select the type of attention deficit/hyperactivity disorder that you have been diagnosed with:

Please choose **only one** of the following:

- ◯ Unsure / Don’t Know
- ◯ Combined presentation (ADHD-C) Includes both inattention and hyperactive-impulsive presentation
- ◯ Predominately inattentive presentation (ADHD-PI)
- ◯ Predominately hyperactive-impulsive presentation (ADHD-HI)

#### What age did you first experience symptoms of attention deficit/hyperactivity disorder?

Please write your answer here:

#### Please select the type of substance use disorder that you have been diagnosed with:

Please choose **all** that apply:

- ☐ Alcohol-related
- ☐ Cannabis-related
- ☐ Hallucinogen-related
- ☐ Inhalant-related
- ☐ Opioid-related
- ☐ Sedative-, Hypnotic-, or Anxiolytic-related
- ☐ Stimulant-related
- ☐ Tobacco-related

#### What age did you first experience symptoms of substance-use disorder?

Please write your answer here:

#### Please select the major eating difficulties that you have been diagnosed with over the course of your lifetime:

Please choose **all** that apply:

- ☐ Anorexia
- ☐ Bulimia
- ☐ Binge-eating disorder

#### What age did you first experience symptoms of major eating difficulty?

Please write your answer here:

#### Please select all the major sleep difficulties that you have been diagnosed with over the course of your lifetime:

Please choose **all** that apply:

- ☐ Insomnia
- ☐ Narcolepsy
- ☐ Hypersomnolence disorder
- ☐ Sleep apnoea
- ☐ Non-Rapid Eye Movement Sleep Arousal Disorders (Sleepwalking, sleep terrors)
- ☐ Nightmare disorder

#### What age did you first experience symptoms of any major sleep difficulty?

Please write your answer here:

#### What age did you first experience Premenstrual dysphoric disorder?

Please write your answer here:

#### The following questions will ask you about any other medical conditions or disorders you may have had in your lifetime. This information may be able to assist us in looking at other medical conditions that could influence the onset or severity of mental illness.

#### Have you been diagnosed with any other medical conditions apart from depression?

Please choose **only one** of the following:

- ◯ Yes
- ◯ No

#### Please select the medical conditions you have from below:

Please choose **all** that apply:

- ☐ **Cancers/Neoplasms**
- ☐ **Diseases of the circulatory system**
  *e.g.: rheumatic, hypertensive, ischaemic heart diseases, diseases of arteries/veins/lymphatic vessels/nodes, high cholesterol*
- ☐ **Endocrine, nutritional and metabolic diseases**
  *e.g.: thyroid gland, diabetes, endocrine glands, malnutrition, obesity*
- ☐ **Diseases of the nervous system**
  *e.g.: inflammatory diseases, Parkinson's, Alzheimer's, neurodegenerative diseases*
- ☐ **Other**

#### Do you have any of these medical conditions?

Please choose **all** that apply:

- ☐ **Infectious and parasitic diseases**
  e.g.: hepatitis, STDs, tuberculosis, herpes, mosquito-related diseases, tick-related diseases , influenza, viral or bacterial infections
- ☐ **Diseases of the blood and blood-forming organs and disorders involving the immune mechanism**
  e.g.: anaemias
- ☐ **Diseases of the eye and adnexa**
- ☐ **Diseases of the ear and mastoid process**
- ☐ **Diseases of the respiratory system**
  **e.g.: lung diseases, respiratory tract diseases**
- ☐ **Diseases of the digestive system**
  **e.g.: intestines, liver, pancreas, stomach, hernia**
- ☐ **Diseases of the skin and subcutaneous tissue**
- ☐ **Diseases of musculoskeletal system and connective tissue**
  **e.g.: diseases of muscles/joints**
- ☐ **Diseases of the genitourinary system**
  **e.g.: kidney, urinary system, breasts, genital organs**
- ☐ **Congenital malformations, deformations and chromosomal abnormalities**
  **e.g.: spina bifida, encephalocele, down's syndrome**
- ☐ **Symptoms, signs and abnormal clinical laboratory findings, not elsewhere classified**
  **e.g.: abnormalities of heart beat, pain in throat and chest, heartburn, dysphagia, faecal incontinence, polyuria, urethral discharge, coma, senility**
- ☐ **Injury, poisoning and certain other consequences of external causes**
  **e.g.: fracture, dislocation, sprain, foreign material entering body, burns, frostbite, drug poisoning, alcohol poisoning, asphyxiation, transplantation rejection**

#### Do you have a diagnosis of diabetes?

**Please choose only one of the following:**

- ◯ **Yes**
- ◯ **No**

#### Which type of diabetes have you been diagnosed with?

**Please choose only one of the following:**

- ◯ Type I
- ◯ Type II

#### What age were you diagnosed with diabetes?

**Please write your answer here:**

#### Have you been treated for high cholesterol levels within the last 2 years?

**Please choose only one of the following:**

- ◯ Yes
- ◯ No

#### Are you currently taking medication for high cholesterol?

**Please choose only one of the following:**

- ◯ Yes
- ◯ No

#### What is the name of the cholesterol medication?

**Please write your answer here:**

#### At what age did you start taking medication for high cholesterol?

**Please write your answer here:**

#### Were you treated for high blood pressure within the last 2 years?

**Please choose only one of the following:**

- ◯ **Yes**
- ◯ **No**

#### Are you currently taking medication for high blood pressure?

**Please choose only one of the following:**

- ◯ **Yes**
- ◯ **No**

#### What is the name of the medication you are taking for high blood pressure?

**Please write your answer here:**

#### At what age did you start taking medication for your high blood pressure?

**Please write your answer here:**

**Do you have a “mind’s eye”? For example, when you think of an apple how clear is the image you see?**

**While most people are able to conjure an image of a scene or face in their minds, some people cannot and are unable to visualize any type of image in their head. These individuals have no "mind's eye," or their imagination is essentially blind. This ability to visualize events and images plays an important part in people's lives.**

**For example, when you think of an apple how clear is the image you see? Using the example shown below compare the image of an apple you can visualise to the picture; is it clear and colourful , dull or blank?**

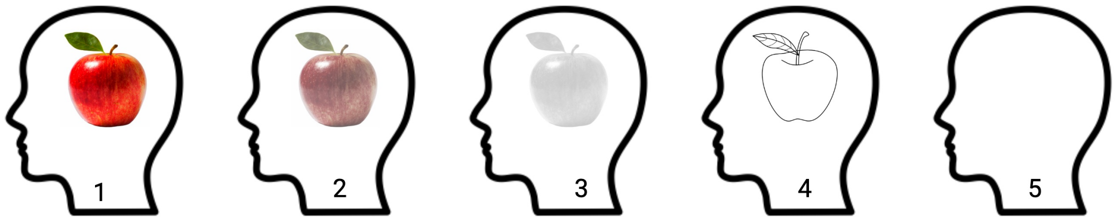

**Now think of a shining star, how clear is this image and using the example above select the option which the image most closely resembles.**

Please choose only one of the following:

- ◯ 1
- ◯ 2
- ◯ 3
- ◯ 4
- ◯ 5

**Musical “earworms” refer to the phenomenon of having songs “stuck” in one's head. It is an involuntary musical imagery that is usually repetitive and can be considered as a form of musical memory.**

**How often in a given day do you experience the musical earworms?**

Please choose only one of the following:

- ◯ Not at all
- ◯ Occasionally
- ◯ 1-2 times per day
- ◯ 3-5 times per day
- ◯ a lot, more than 5 times per day

**Is there any other information you would like to share with us regarding your health that this survey has not already covered? It may also include other information about your personal mental health journey that you feel is important for us to know.**

Please write your answer here:

**This is the end of the questionnaire in relation to asking you about your depression, your medications and your mental health journey. Thank you very much for your commitment to completing this questionnaire, your information is invaluable to this project.**

**If completing this questionnaire has raised any issues for you, please reach out to the following groups, who can provide advice, support, and help:**
**Beyond Blue / Lifeline / Pregnancy Loss Australia**

**You have agreed to provide a blood sample and so we would now like to confirm some details around this part of your participation in this project.**

### Providing a Biological Sample

**Thank you for completing the questionnaire. Are you still willing to provide a blood sample?**

**Please choose only one of the following:**

- ◯ Yes
- ◯ No

#### Please enter the most convenient address for us to send your the blood sample kit. This could be different to your residential/work address.

**Please write your answer(s) here:**

- **Street Address**
- **Suburb**
- **State**
- **Postcode**

#### We will now prepare your sample kit. Please confirm if you are happy to receive this in the next two weeks.

**Please choose only one of the following:**

- ◯ **I confirm**
- ◯ **Delay sample kit**
  ***(e.g. if you are unable to collect kit in the next two weeks due to being away)***

#### From what date would you like to receive your sample kit?

**Please enter a date:**

**Once we have received your sample and questionnaires, we would like to provide you with an $50 reimbursement. Would you like to receive this reimbursement as a gift card or donate the reimbursement back to our research?**

Please choose only one of the following:

- ◯ Gift card
- ◯ I would like to donate my reimbursement back to this project

**Thank you very much for your contribution.**

Thank you very much for completing the Cell-O questionnaire. Your responses have been securely recorded in our research database. We will be in touch with you shortly if you have agreed to provide a blood sample to us.

Thanks again on behalf of Professor Naomi Wray and the Cell-O research team!

Submit your survey.
Thank you for completing this survey.
